# A user-driven framework for dose selection in pregnancy: proof-of-concept for sertraline

**DOI:** 10.1101/2024.03.19.24304542

**Authors:** CJM Koldeweij, AC Dibbets, BD Franklin, HCJ Scheepers, SN de Wildt

## Abstract

Despite growing knowledge of pregnancy-induced changes in physiology that may alter maternal and fetal pharmacokinetics, and therefore drug efficacy and safety, evidence-based antenatal doses are lacking for most drugs. Pharmacokinetic models and expanding clinical data in pregnancy may support antenatal doses. In this article, we introduce a comprehensive and user-driven Framework for Dose Selection in Pregnancy (FDSP), developed and validated to support the clinical implementation of best-evidence and in some cases, model-informed doses for pregnant women and/or fetuses. After initial development and validation by experts, the framework prototype was piloted to formulate an antenatal dosing strategy for sertraline in depression and anxiety disorders. Next, the framework was validated and assessed for usability by a multidisciplinary working committee of end-users comprising healthcare practitioners, experts from other disciplines including pharmacometrics, reproductive toxicology and medical ethics, alongside pregnant women and a partner. The resulting framework encompasses the following: rationale for drug selection, a comprehensive analysis of pharmacokinetic and dose-related efficacy and safety data, and implementation aspects including feasibility and desirability of the recommended antenatal dose based on a structured maternal and fetal benefit-risk assessment. An antenatal dose recommendation for sertraline, as a proof-of-concept, was formulated using this approach and endorsed for clinical use by the working committee. The FDSP, as demonstrated by the example of sertraline, is fit for supporting the development of best-evidence acceptable and clinically feasible antenatal doses.

## Introduction

Pregnancy-induced changes in maternal physiology and placental transfer may warrant dose alterations for certain drugs. Examples of maternal changes affecting drug pharmacokinetics in pregnancy include an increased plasma volume, augmented renal filtration and changes in the expression and activity of drug metabolizing enzymes(1–3). These pharmacokinetic changes, along with potential alterations in pharmacodynamics, may affect drug safety and efficacy during pregnancy, in some cases warranting adjusted antenatal doses(4). However, while over 80% of pregnant women use medications(5), specifically researched antenatal doses are lacking for most drugs(4, 6). Despite efforts to increase enrollment of pregnant women in clinical research(7, 8), ethical concerns about potential fetal harm and the lack of obligation for pharmaceutical companies to investigate antenatal dosing before market access contribute to a paucity of data to support evidence-based antenatal doses(9, 10). Information on the pharmacokinetics, safety and efficacy of drugs in pregnancy remains scarce (6, 11–14). In the absence of antenatal dosing guidance, clinicians caring for pregnant women often prescribe doses intended for non-pregnant adults, or, sometimes, reduce these doses due to concerns over fetal harm(4). Given altered pharmacokinetics and pharmacodynamics during pregnancy, such practices may result in suboptimal safety and efficacy for certain drugs.

In this context, the emergence of pharmacokinetic modelling as an additional tool to support antenatal dosing appears promising. Alongside traditional pharmacokinetic studies, population-based (pop-PK) and physiologically-based (PBPK) pharmacokinetic models can be used to simulate maternal and fetal drug exposures throughout pregnancy based on limited clinical data(15, 16).

The feasibility of issuing antenatal doses drawing on pharmacokinetic models alongside clinical and animal data is currently being investigated as part of project MADAM (Model-Adjusted Doses for All Mothers). The resulting doses will be accessible to HCPs and patients as part of an online resource on antenatal doses. In line with other clinical guidance but also, given the specific challenge of selecting doses that optimize maternal and fetal risks and benefits despite limited evidence(17), making antenatal dose recommendations requires a systematic methodology. Therefore, we leveraged the insights of experts and patients convened as part of project MADAM to develop a user-driven Framework for Dose Selection in Pregnancy (FDSP), taking into account both pharmacological evidence and implementation considerations. In this article, we report on the development, validation and pilot use of the FDSP for the antidepressant sertraline as a proof-of-concept.

## Methods

### Aims of the framework

The FDSP was conceived as a comprehensive framework for compiling, analyzing and reporting data, along with other considerations, to support the issuing and clinical implementation of antenatal doses. It can be applied to examine drug doses for addressing maternal indications (either de novo or for continued use in pregnancy) or fetal conditions, encompassing both prescription and over-the-counter drugs. Tailored to facilitate dosing decisions by the working committee of project MADAM, the design of the FDSP accommodates usage by other formulary and guideline committees. Key considerations in FDSP development were ensuring completeness by incorporating all relevant aspects for determining evidence-based, acceptable and feasible antenatal doses, and usability for both those completing the framework, and reviewers assessing the proposed antenatal doses. Another priority was the leanness of the framework, to facilitate ease and speed of completion and review. Alongside guiding antenatal dose review by formulary committees, the framework was designed for reporting the underlying evidence and considerations to healthcare practitioners and pregnant women for the recommended doses.

### Sources

Alongside clinical expertise, the FDSP draws on two additional sources. First it expands on a method for benefit-risk assessment of off-label pediatric drug use and dosing (BRAVO)(18). The BRAVO framework is based on the “Problem, Objectives, Alternatives, Consequences, Trade-offs, Uncertainty, Risk attitudes, and Linked decisions” (PrOACT-URL) decision-making guide(19). The PrOACT-URL methodology is used as a benefit-risk methodology by the European Medicines Agency (20, 21). It comprises nine sections: 1) problem definition (scope for the dose recommendation including medical indication and alternatives), 2) efficacy and 3) safety in relationship to the 4) dose, followed by five sections to assess dose feasibility based on the 5) consequences, (6) trade-offs, 7) uncertainty, 8) risk tolerance and 9) linked decisions (e.g. a patient’s informed consent). Second, the FDSP was developed to address the distinct challenges of antenatal dose selection, including limited *in vivo* pharmacokinetic evidence (12), implying greater reliance on modelling and simulations(22), and the need to balance potentially misaligned maternal and fetal risks and benefits(17). Therefore FDSP design drew on insights from an international stakeholder analysis (submitted for publication).

This analysis, conducted among European pregnant women and healthcare practitioners, explored the acceptability and perceived barriers and facilitators for model-informed antenatal doses. It revealed a high willingness-to-use such doses among both groups subject to certain conditions, including transparent communication on rationales for altered antenatal doses and details on the quality of appraised evidence, including from pharmacokinetic models.

### Framework development and validation

FDSP development occurred in three stages and followed a user-driven approach(23).

**- Prototype development***–* The initial FDSP comprised four sections, each with several questions and suggested information sources. The prototype was written in English to enable international use. Section 1 on ‘Drug selection’ examined the rationale for investigating a drug dose in pregnancy. Section 2 on ‘Dose selection’ aimed to compile and analyze all pharmacokinetic, efficacy and safety data related to antenatal dosing, alongside information on current dosing and administration practices. Section 3 on ‘Dose implementation’ was designed to evaluate the acceptability of a proposed dose considering the maternal and fetal risks and benefits, along with an appraisal of the dose certainty and feasibility. Section 4 outlined the final ‘Dose recommendation’ for clinical use.

**- Validation by experts** - The prototype underwent validation by the following experts: one obstetrician-gynecologist, one pediatrician-pharmacologist, one toxicologist, two pharmacometricians, and one formulary expert. Key aspects for review included the relevance, comprehensiveness and clarity of the framework. Alongside face validation, the prototype underwent iterative adaptations through rapid cycle design (Appendix 1)(24).

**- Piloting for sertraline, validation and usability assessment by end-users** - Next, the FDSP was piloted for sertraline, chosen as a case study as the most widely used antidepressant in pregnancy(25). Two researchers (CK, medical doctor trained in social sciences and CD, medical student, supervised by LS, obstetrician-gynecologist, and SNW, obstetrician and pediatric-clinical pharmacologist) completed the framework for sertraline. Next, a national committee of 22 experts and representatives with diverse experience in the Netherlands (Figure 1), recruited under project MADAM, appraised the proposed dose for clinical use and assessed the validity and usability of the FDSP in this process. The committee offered written and plenary feedback on the relevance, comprehensiveness and user-friendliness of the FDSP as applied for sertraline (Appendix 1). Framework usability was rated using the System Usability Scale(26).

**Figure 1–.**
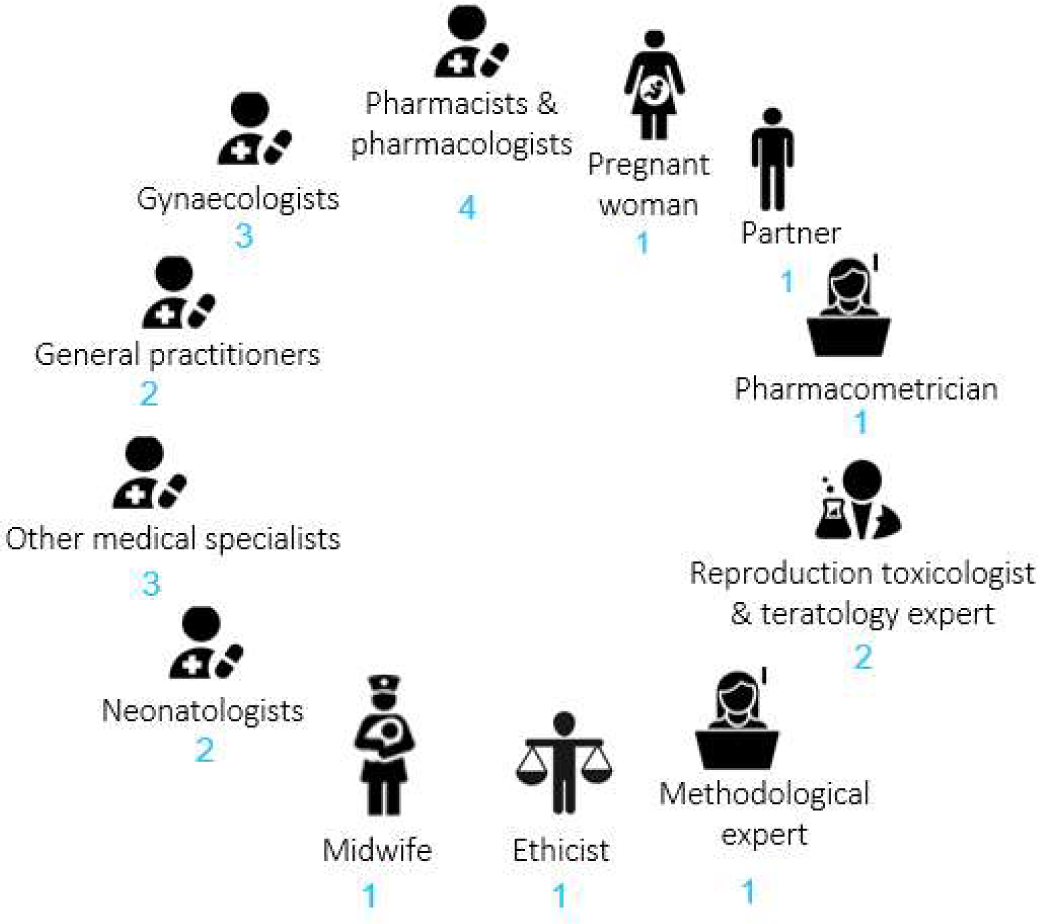
Composition of the FDSP reviewing committee. FDSP: framework for dose selection in pregnancy

## Results

### Generic framework

The resulting framework is presented in Table 1.

**Table 1–.**
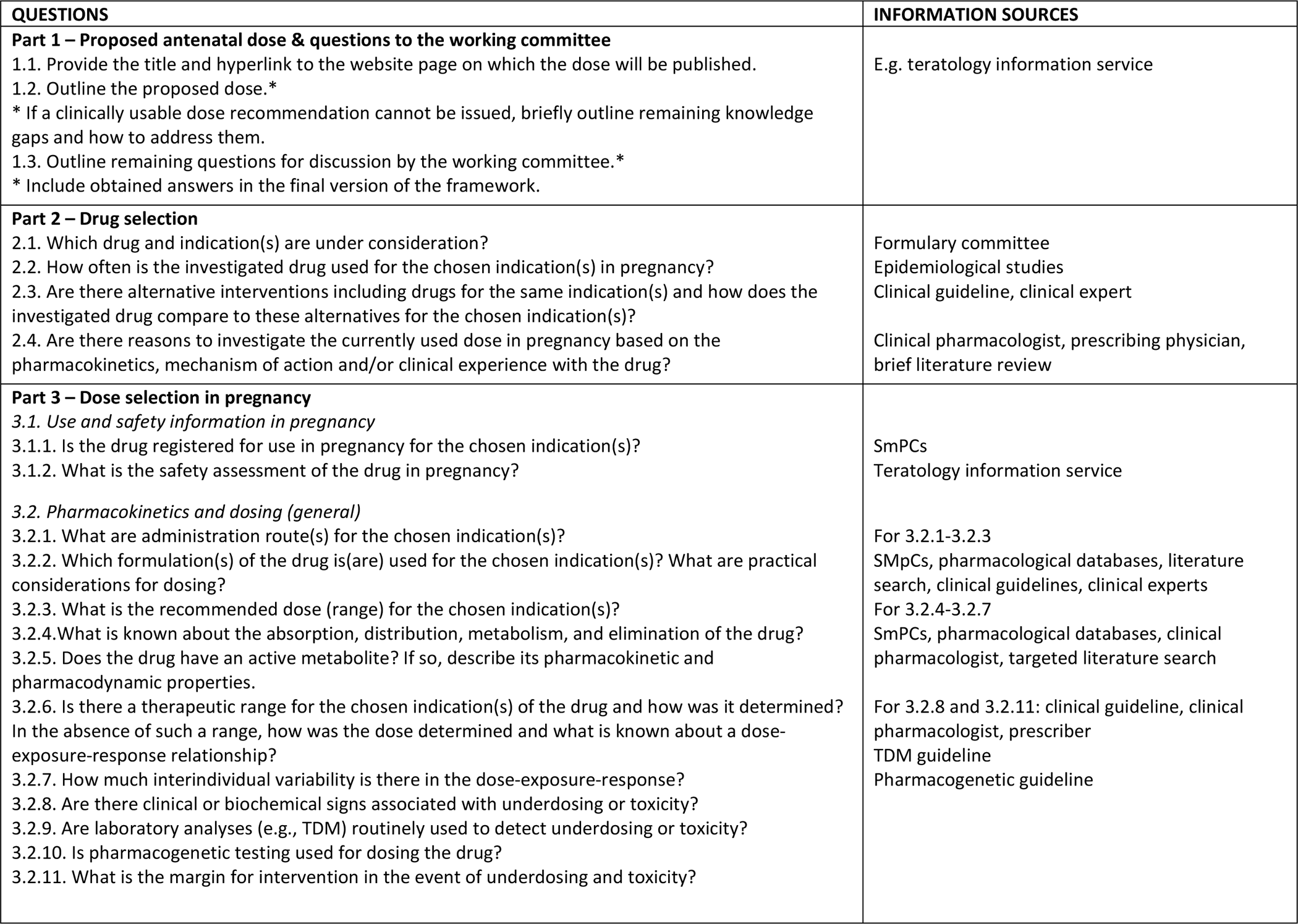
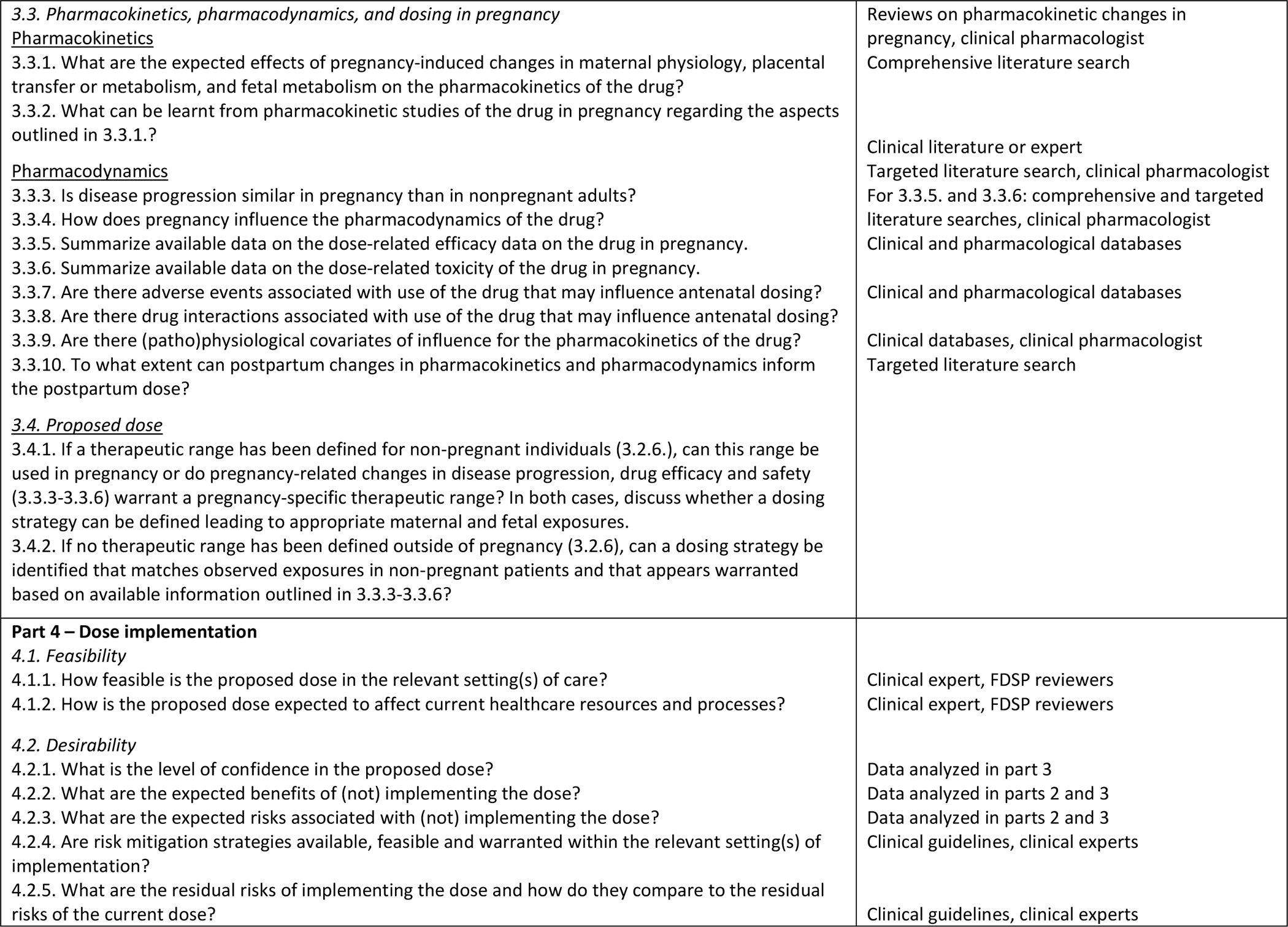
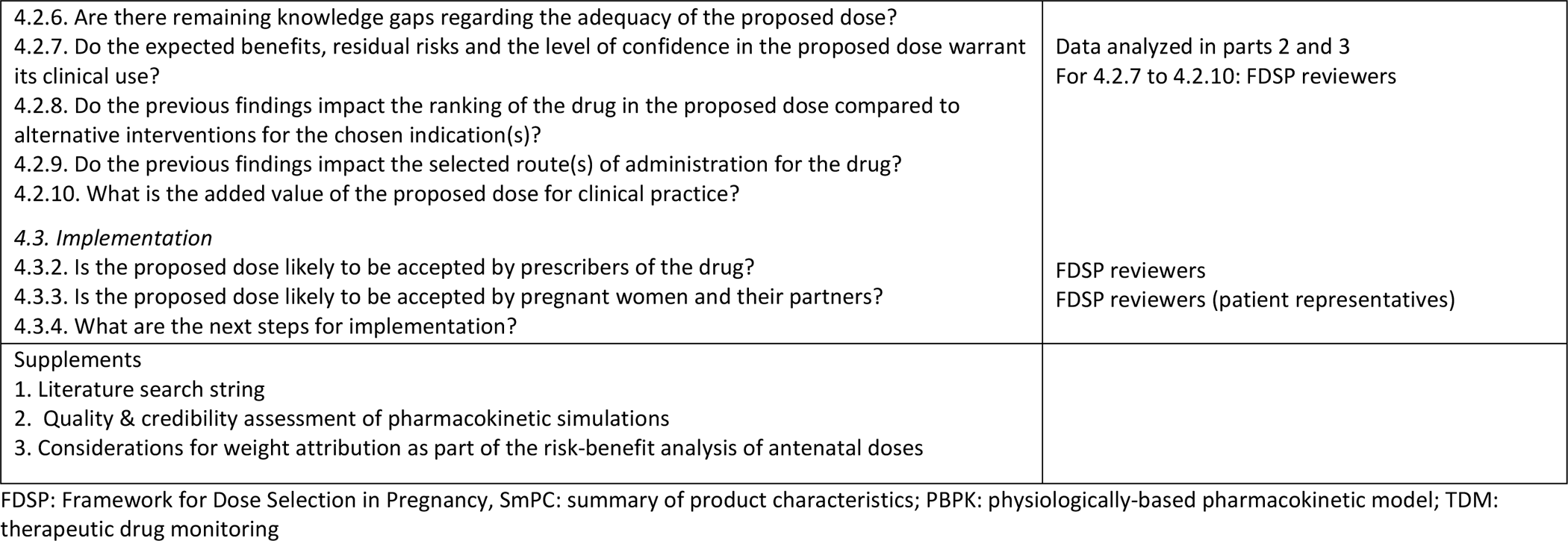
Framework for Dose Selection in pregnancy.

The antenatal dose review process using the FDSP is delineated in Figure 2. The first stage is selection of a drug for antenatal dose review. Drug selection criteria may include clinical factors such as frequency of use in pregnancy, evidence of suboptimal treatment with current doses, and availability of pregnancy-related pharmacokinetic data. Data for framework completion can be obtained from a general literature search, supplemented by targeted searches and expert insights. Framework completion follows a modular approach, allowing for questions to be selectively addressed depending on the drug indication (maternal or fetal), data availability and formulary needs. Following appraisal by the reviewing committee, the completed FDSP should be amended to reflect input from reviewers before publication of the dose, if deemed ready for use.

**Figure 2–.**
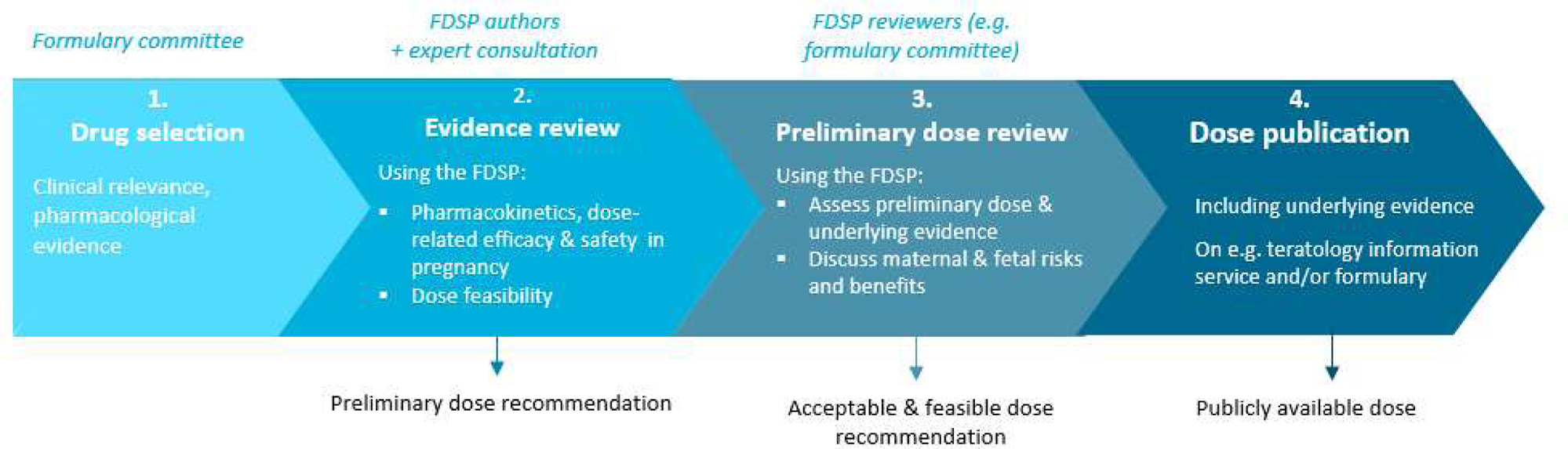
Steps for issuing an antenatal dose using the Framework for Dose selection in Pregnancy. FDSP: Framework for Dose selection in Pregnancy, MELINDA: model-informed dosing for all

### Framework piloting

The piloted FDSP for sertraline in the Netherlands is outlined in Appendix 2, with each section described and exemplified for sertraline below.

**Part 1: Dose recommendation** *–* The first FDSP section outlines the proposed antenatal dose recommendation derived from analyses in subsequent sections. The wording of this recommendation should convey the degree of certainty in the available evidence following the Grading of Recommendations Assessment, Development, and Evaluation (GRADE) guidelines and is subject to endorsement by FDSP reviewers(27).

**Antenatal sertraline dose** a) For women who initiated sertraline use before pregnancy: maintain the dose used before pregnancy. Increased sertraline clearance may lead to a reduced effect in the second and third trimesters. If symptoms worsen during pregnancy, a dose increase should be considered, following titration steps for non-pregnant adults. Doses exceeding 150 mg daily require careful consideration and discussion with the patient.
b) For women starting sertraline during pregnancy: follow standard guidance for dose selection and titration. The antenatal dose recommendations for women who started sertraline before pregnancy otherwise apply.
Cytochrome P450 (CYP)2C19 variants can impact the required sertraline dose. Pharmacogenetic testing should be considered if unexplained side effects or an inadequate response to sertraline are reported during pregnancy, especially in women with a history of such issues with medications metabolized by CYP2C19. For pregnant women with a CYP2C19 variant:

- Normal or ultrarapid metabolizers: maintain the pre-pregnancy dose in prior sertraline users, and adjust as needed based on response and side effects. For new sertraline users, follow guidance for non-pregnant adults for dose initiation and titration.
- Poor metabolizers: the same recommendations apply with a maximum dose of 150 mg daily.
Therapeutic drug monitoring is not routinely advised for sertraline in the Netherlands.

**Part 2: Drug selection –** This section specifies the indication(s) for which the dose is reviewed, alternative drugs for treating these condition(s) and rationale for selecting the drug for antenatal dose review.

**Drug and indications** Sertraline is a commonly prescribed antidepressant in pregnancy in the Netherlands and internationally(25, 28). Indications include depression and anxiety disorders(29, 30).
**Alternatives** Serotonin reuptake inhibitors (SSRIs) are a second-line treatment for depression and/or anxiety disorders(29, 30). Initial or combined treatment involves non-pharmacological interventions including psychotherapy. During pregnancy, the relative preference for non-pharmacological interventions is greater to minimize potential fetal risks(31).
**Rationales for antenatal dose review** Pregnancy-induced changes in sertraline distribution and metabolism may lower maternal exposure to the drug, potentially impacting treatment efficacy during pregnancy(32, 33).

**Part 3: Dose selection**– The third FDSP section comprises information regarding current use, safety and dosing of the drug along with pharmacokinetic and pharmacodynamic data relevant to antenatal dosing. Drug use aspects include route(s) of administration, standard doses and formulations for the chosen indication(s), and additional dosing considerations such as strategies to identify underdosing or toxicity. Pregnancy-related information, including licensing information and available antenatal dosing guidance, if any, should be described, alongside maternal and fetal risks from antenatal drug use. Relevant pharmacokinetic data include the ADME (absorption, distribution, metabolism, and elimination) properties of the drug, maternal and fetal drug exposures throughout gestation, along with placental transfer. Data can be extracted from traditional, physiologically-based and population-based pharmacokinetic studies. The quality of pharmacokinetic studies can be graded using the adapted Jadad classification(34), with specific questions in the FDSP for appraising the credibility of available pharmacokinetic models. Furthermore, information should be gained on a therapeutic range, or, in its absence, a routinely observed concentration range, alongside available data on dose-related efficacy and safety. These concentration ranges may be defined outside of pregnancy (in non-pregnant adults or in neonates) and during gestation (in pregnant women and fetuses). For drugs with limited human data on dose-related efficacy and safety during pregnancy, the applicability of animal data should be assessed. In the absence of a therapeutic range or reference concentration range in pregnancy, one should infer the influence of pregnancy on the dose-exposure-response drawing on available pharmacokinetic and pharmacodynamic data in pregnancy. Other relevant pharmacodynamic data encompass the influence of pregnancy on disease progression and the pharmacodynamics of the considered drug, along with drug interactions, adverse events and covariates that may influence antenatal dosing. Lastly, Part 3 includes details on pharmacokinetic and pharmacodynamic changes that may influence postpartum dosing. Taken together, these data should inform a dosing strategy that ensures adequate maternal and/or fetal exposure(s) throughout relevant gestational ages for the chosen indication(s), aligning with the identified target concentration range(s) and considering pharmacodynamic aspects in pregnancy. For drugs lacking a therapeutic range, appropriate antenatal doses may be extrapolated from non-pregnant adult doses by incorporating pharmacokinetic changes in pregnancy (e.g. a 30% reduction in maternal exposure may warrant a 30% dose increase, assuming a similar concentration-response relationship; the impact of such dose alterations on maternal and fetal drug safety is addressed in Part 4).

**Use and safety in pregnancy** *Sertraline is not licensed for use in pregnancy in Europe or the United States*(*32, 35*)*. However, it is a preferred SSRI for pregnancy given a favorable safety profile*(*36*)*. Neonatal adverse events associated with antenatal use include persistent pulmonary hypertension of the newborn (PPHN) (baseline incidence 2 cases per 1000 births; odds ratio 2.01 (95% CI 1.32-3.05) in sertraline-exposed children after 20 weeks of gestational age) and moderate neonatal adaptation syndrome (NAS), affecting 25 to 30% of exposed neonates during the third trimester*(*37*)*. While PPHN is a life-threatening condition requiring multiple medical interventions*(*38*)*, NAS is transient and self-limiting. Because of these two risks, it is recommended that fetuses exposed to sertraline in the third trimester be delivered at hospital for observation and potential treatment*(*36*)*. Lastly, SSRI use may be associated with a slightly elevated risk of septal defects*(*39, 40*)*. Study findings are inconsistent concerning a potential link with small-for-gestational-age*(*41, 42*).
**Pharmacokinetics** Sertraline is administered orally in a 25-200 mg daily dosing range in non-pregnant adults(29, 35). After initiation of a low dose, sertraline dosing is individually titrated based on response and side effects. Sertraline exhibits a complex hepatic metabolism involving five CYP enzymes(43–45). In non-pregnant adults, interindividual variation in sertraline pharmacokinetics is most likely related to CYP2C19 polymorphisms, warranting CYP2C19 genotyping in certain patients for specific indications(46, 47). A comprehensive literature search identified 14 pharmacokinetic studies in pregnancy, and one maternal-fetal physiologically-based pharmacokinetic model for sertraline(48–63). Of heterogeneous quality, these studies suggested that pregnancy-induced alterations in maternal physiology may result in a moderate decrease (15-20%) in the maternal plasma concentration of sertraline, especially in the second and third trimesters, compared to baseline. This decrease can most likely be attributed to the combined effect of CYP enzyme induction (excluding CYP2C19) and a decrease in albumin concentration during pregnancy(33, 64). Similar to non-pregnant adults, maternal exposure to sertraline is marked by interindividual variability likely derived from genetic variation in CYP2C19(48, 50–58, 60–62, 64, 65). Placental transfer of sertraline ranges between 30-40%(51–53, 55–57, 61, 65, 66). These pharmacokinetic findings imply that, to maintain sufficient maternal exposures, sertraline doses should not be reduced during pregnancy and may even have to be increased for certain women in the second or third trimester depending on their clinical response.
**Pharmacodynamics** Our understanding of the influence of pregnancy on sertraline efficacy is insufficient to inform antenatal dosing. Given large interindividual variation in dose-exposure and dose-response in non-pregnant adults, a therapeutic range for sertraline is lacking(46, 67, 68). However, a dose-response relationship appears likely at an individual level. While data on dose-related fetal toxicity are limited (none for small-for-gestational-age or PPHN)(38, 40), evidence suggests a dose-relationship between moderate NAS and sertraline doses exceeding 100 mg in the third trimester (relative risk 3.31 (95% CI 1.05-10.39))(69). This dose-effect relationship may extend to other fetal or neonatal outcomes determined in the second or third trimester of fetal development (thus excluding septal defects formed earlier in pregnancy).

**Part 4: Dose implementation** – Lastly, this section examines the feasibility, desirability, and steps for implementation of the proposed dose based on data analyzed in part 3. Part 4 invites users to appraise technical, logistical and ethical considerations to determine whether clinical implementation is feasible and warranted. This assessment may incorporate expert insights and FDSP reviewers’ perspectives. Dose feasibility takes into consideration aspects including available drug formulations, drug preparation and administration, along with the resource implications of implementing the proposed dose in relevant settings of care. Desirability of the proposed dose can be determined by first describing the level of confidence in the dose, given the certainty of the reference concentration range and overall assessment of the relevance, completeness, and quality of underlying pharmacological data(70, 71). Second, maternal and fetal risks and benefits associated with the dose should be summarized drawing on information collected in part 3. Risk mitigation strategies (e.g. follow-up of clinical symptoms or therapeutic drug monitoring to detect underdosing or toxicity) can be outlined to determine residual risks (i.e., the remaining risks when risk mitigation strategies are in place)(18). The residual risks and benefits should be weighed against each other and against the risks and benefits of not implementing the dose. Remaining knowledge gaps regarding dose adequacy should be described. Drawing on this analysis, FDSP reviewers can assess whether the proposed dose can reasonably be implemented in clinical care. In instances that it cannot, practical recommendations on how to address the remaining knowledge gaps should be issued. Lastly, part 4 involves considering the acceptability of the proposed dose for healthcare practitioners and pregnant women and defining remaining implementation steps.

**Dose feasibility** The suggested antenatal sertraline dose aligns with the standard dosing range for non-pregnant adults(32, 35), and thus appears feasible across relevant care settings.
**Dose desirability** Despite the moderate to low quality of available pharmacokinetic data, confidence in the proposed dose appears sufficient. Notwithstanding interindividual variation in sertraline pharmacokinetics and the lack of a well-defined therapeutic range, available data suggest a likely reduction in maternal exposure during pregnancy for most women. Dose titration based on individual symptoms thus appears warranted, especially in the second and third trimesters. Considering individual dose titration and the recommended antenatal dose falling within the standard dosing range for non-pregnant adults (with 150 mg as a suggested maximum dose out of caution given potential CYP2C19 poor metabolizers), the likelihood of maternal overdosing appears low. Looking at fetal toxicity, an elevated risk of NAS, with a proven dose-response relationship in the third trimester(69), should be weighed against the moderately severe and self-limiting nature of this condition(72). While not demonstrated, a dose-response relationship may exist for other neonatal outcomes, PPHN being most critical given its life-threatening prognosis(73). Overall, given the self-limiting nature of NAS and the low absolute incidence and likely limited risk elevation of PPHN with increased sertraline doses in the second and third trimesters(37) these risks were considered outweighed by the expected benefits of adequate treatment through dose titration. Likely maternal benefits comprise improved well-being and a reduced risk of postpartum depression, along with indirect fetal and neonatal benefits including a reduced likelihood of perinatal complications linked to maternal stress, and enhanced postpartum bonding(40).
**Implementation steps** The sertraline dose will be published on the website of the Dutch Teratology Information Service. Implementation can be facilitated by educating clinicians and pregnant women about the importance of maintaining adequate sertraline levels during pregnancy, which may in some cases require a dose increase to keep depression and/or anxiety symptoms under control.

### Framework usability

Ten committee members, including healthcare practitioners, non-clinical experts and patients, rated the usability of the FDSP as reviewers. The willingness-to-use the framework was high (median score 4.5 out of 5), as was the score for integrity (median score 4.0) with the framework scoring slightly lower on ease of use, unnecessary complexity and need for additional technical support (median scores 3.5, 2 and 2.5 out of 5 respectively), while the received a median score of 2.5 out of 5). The framework was perceived as convenient to use. Confidence in using the framework received a median score of 3.5, but committee members expected to learn how to use the framework quickly (median score 4.0).

## Discussion

We introduced a Framework for Dose Selection in Pregnancy that was applied to sertraline and validated by a multidisciplinary committee of experts and patients.

### Added value

To our knowledge, this is the first available framework to guide antenatal dose selection based on existing data. As such, it may provide the impetus and necessary guidance for guideline committees to establish adequate doses in pregnancy, thereby addressing an unmet health need(4, 11, 12, 14). While other dose selection frameworks exist focusing on pediatric dosing(74), or the general population(75), these are not fit-for-purpose for issuing best-evidence, and potentially, model-informed, antenatal doses. One reason is the need to balance maternal and fetal risks and benefits of proposed antenatal doses based on available pharmacological data(9, 17). Secondly, given scarce pharmacokinetic data in pregnancy(11–13), antenatal doses are likely to often draw on pharmacokinetic modelling and simulations(14, 15). However, despite recent attempts to introduce a blueprint for issuing model-informed doses(76), comprehensive tools for appraising the credibility of pharmacokinetic simulations alongside other evidence to inform clinical practice are lacking(77, 78). To address these challenges, framework design drew on a stakeholder analysis on perceived barriers and facilitators for implementing model-informed antenatal doses. In addition, intended FDSP users including healthcare practitioners, patients, and other experts were involved in FDSP development and validation, following a multidisciplinary, user-driven design approach. The obtained framework supports the assessment of a broad range of evidence, including from pharmacokinetic models. Drawing on clinical pharmacological principles(18), it can guide antenatal dose selection in the absence of a therapeutic range, as shown for sertraline. For fetal drugs, extrapolations could be made drawing on neonatal pharmacokinetic and/or pharmacodynamic data. Furthermore, the FDSP invites its users to assess the quality of both traditional and simulation-based pharmacokinetic studies used, enhancing the credibility of issued (model-informed) doses. Beyond supporting a systematic evidence review, the FDSP can be used to appraise the maternal and fetal risks and benefits of considered antenatal doses, helping to determine acceptable doses amid uncertainty. It also incorporates key considerations for dose feasibility. Lastly, the FDSP allows transparent reporting of these dosing considerations to healthcare pactitioners and patients, thus fostering adoption of recommended doses.

### Limitations and implications for research and practice

Despite the added value of the FDSP, several limitations apply. First, framework completion appears time-consuming given the extensive data analyses involved. While this was not specifically undertaken for sertraline, the timeframe required for completion will be assessed in future FDSP uses, and a simplified framework may be developed to enhance efficiency of use. Second, while we aimed to make the FDSP comprehensible for reviewers with diverse backgrounds, its parts may differ in accessibility for readers with varying clinical and pharmacological knowledge. However, despite some FDSP reviewers having selectively read information outlined for sertraline, the multidisciplinary nature of the reviewing committee allowed members to leverage each other’s expertise to form an informed opinion collectively. In our view, this underscores the advantages of a multidisciplinary dose review also involving patients. This can help ensure the incorporation of diverse perspectives as part of complex decision-making(79), a practice whose benefits have previously been acknowledged(80). FDSP use by other formulary committees for various drugs can offer more insights into the applicability of this methods for antenatal dosing. Third, while the FDSP was assessed for face and content validity, a next validation step may involve establishing whether the resulting doses are associated with improved maternal and fetal outcomes compared to standard dosing(81). Fourth, to avoid duplication of efforts and accelerate the international adoption of best-evidence antenatal doses, we encourage FDSP use by other guideline committees and open sharing of the endorsed doses and underlying data. While specific implementation considerations may differ among countries, pharmacological analyses are likely to apply internationally. Key dissemination channels for the obtained doses include academic publications along with national teratology information services, clinical guidelines and formularies. In pediatrics, country or population-specific formularies are provide examples of implementation of off-label drug use in clinical practice(74, 82, 83).

## Conclusions

We presented a comprehensive and user-driven framework incorporating various types of evidence and considerations to inform the selection of best-evidence, acceptable, and clinically feasible doses in pregnancy. In the absence of other tools for issuing systematically researched antenatal doses, this framework can help bridge an important public health gap by enhancing maternofetal therapies. Although the FDSP was designed for antenatal dose selection, lessons from its use may also inform dosing for other patient groups lacking best-evidence doses.

## Data Availability

All data produced in the present work are contained in the manuscript

## Study highlights

**What is the current knowledge on the topic**? Despite growing understanding of pregnancy-induced changes in physiology that may alter maternal and fetal drug efficacy and safety, evidence-based antenatal doses for clinical practice are missing for most drugs.

**What question did this study address?** Can a rigorous and user-driven methods be developed to issue evidence-based, acceptable and clinically feasible antenatal doses?

**Does the study add to our current knowledge?** In this article, we introduce a user-driven Framework for Dose Selection in Pregnancy. This framework has been developed and validated to support the clinical implementation of best-evidence and in some cases, model-informed antenatal doses, that are also acceptable and clinically feasible. Applicability of this framework is illustrated through its pilot use for issuing an antenatal dose recommendation for sertraline.

**Might this change clinical pharmacology or translational sciences?** In the absence of tools for issuing systematically researched and clinically feasible antenatal doses, this framework for antenatal dose selection can help improve maternal and fetal therapies, addressing a crucial public health gap. The proposed antenatal sertraline dose has been endorsed for use in Dutch clinical practice.

## Supporting information

Supplementary information accompanies this paper on the Clinical Pharmacology & Therapeutics website (www.cpt-journal.com).

## Acknowledgments

Thanks to the MADAM Working Committee members and research team for their feedback on the framework design and pharmacological analyses for sertraline.

## Author contributions

CK designed and performed the research, analyzing the data together with CD while receiving guidance from SDW and LS on the pharmacological and clinical content and overall framework design. CK additionally wrote the manuscript with editorial assistance from CD. BDF offered advice on the study design.

