## Supplemental files for "A user-driven framework for dose selection in pregnancy: proof-of-concept for sertraline"

### Appendices

**1. Framework adaptations** **p. 2**

1.a. Summary of changes along the FDSP development cycle p. 2

1.b. Proposed and adopted changes based on feedback from the working committee of project MADAM p. 3-8

**2. FDSP piloted for sertraline** **p. 10-67**

2.a. Main document

2.b. Supplements p. 68-78

#### 1. Framework adaptations

##### *1. a. Summary of adaptations as part of the development cycle*

| **Framework development and validation steps** | **Adaptations of the framework** |
|---|---|
| 1 Review and validation by experts | * Order & wording of questions altered, e.g. addition of a question on alternative interventions<br>* More detailed instructions for use: e.g. distinguish between fetal and maternal aspects when analyzing data or considerations to support a dose; distinguish between trimesters of pregnancy |
| 2 Pilot testing for sertraline:<br>- Framework completion<br><br><br><br><br><br><br>- Framework review, validation and usability assessment by ends-users (MADAM working committee)* | <br>* Development of generic literature search string<br>* Addition of an executive summary<br>* Addition of sub-conclusions for FDSP parts<br>* Addition of glossary with pharmacological concepts<br>* Part 4 (‘Dose recommendation’) switched to part 1<br>* Order & wording of questions altered for greater leanness and intelligibility<br><br>* Addition of a patient summary<br>* More detailed information on dose-related toxicity (especially fetal), requiring FDSP authors to compile available data and make hypotheses on dose-related toxicity for various identified fetal adverse events when data is missing (Parts 2 and 3)<br>* Addition of a visual template to appraise the maternal and fetal risks and benefits of proposed antenatal doses |

* A more detailed discussion of framework adjustments discussed by the Working Committee of project MADAM, and a summary of the adopted changes is outlined in Appendix 1.b.

*1.b – Considered and adopted adaptations based on feedback from the working committee of project MADAM*

Abbreviations
DRD: dose rationale document; FDSP: Framework for Dose Selection in pregnancy; HCPs: healthcare practitioners; MELINDA: Model-Informed Dosing website with access to summarized version of DRD for HCPs and patients; MIPF: model-informed pregnancy formulary; WC: Working Committee

M1: doctor-researcher MADAM;  M2: anesthestist; M3: general practitioner; M4: obstetrician-gynecologist; M5: clinical pharmacologist-neonatologist; M6: patient representative; M7: obstetrician-gynecologist, researcher MADAM; M8: partner of pregnant woman; M9: midwife; M10: reproduction toxicologist; M11: pharmacologist; modeler; M12: clinical pharmacologist-paediatrician; researcher MADAM; M13: clinical pharmacologist in training; M14: obstetrician-pharmacologist in training; M15: padiatrician in training; M16: ethicist; M17: obstetrician-gynecologist; M18: general practitioner; former midwife; M19: psychiatrist-clinical pharmacologist; M20: hospital pharmacist; M21: teratology specialist; M22: methodological expert

| Proposed changes in FDSP | Arguments for whether or not to implement the proposed changes | Framework adjustment | Work process adjustment |
|---|---|---|---|
| **Relevance/completeness** | | | |
| For maternal medications:<br>* Outline dose recommendation for **postpartum period** (M3, M13)*<br>* Add a section on **lactation** (M8)* | * Limited data on maternal PK changes in first 6 weeks postpartum, outside of modelling scope<br>* Limited PK data from lactating women | * **Postpartum**: outline dosing guidance & considerations whenever possible<br>*** Lactation:** outside scope | |
| **Readability** | | | |
| <u>Length</u><br><br>* Remove **pharmacological definitions** (M15)<br><br>* Remove questions that **overlap** with framework from **decision tree** (M12)* | * Framework is long (M4, M9, M14, M15, M16, M17, M19)<br>* Framework remains readable when focusing on key sections as outlined in instructions for reading (M6)<br><br>* Definitions are helpful for reading, especially for patients (M6, M9) | * Include definitions in separate **glossary** and remove definitions from HCP version of DRD; keep definitions in **patient version of the DRD** (see description below)<br><br>* Include a short **summary of the decision tree in the DRD** (full decision tree in Appendix); remove questions overlapping with FDSP from decision tree | * Create **Dropbox** with all relevant documents for WC members to access at all times (including glossary) |
| <u>Complexity</u><br>* Organize a limited review of the **decision tree** by experts for each included medication (M7) | * Remains complex for non-modellers (M18)<br>* Need to create transparency & accountability on model-informed dosing (M1, M14) | * Decision tree to be reviewed by experts in MADAM team as part of DRD writing & by WC members that feel competent (decision tree will be included as an | * Online seminar on model-informed dosing in pregnancy and underlying techniques for WC members on 14/11/23 |

| | | | |
|---|---|---|---|
| * Add a **summary for non-healthcare practitioners** (M1)<br><br>* Why are DRDs not written in **Dutch** if meant to inform dosing decisions by Dutch HCPs and pregnant women? (M18) | * DRDs are meant for use and consultation by an international audience in the future (MELINDA website only available in English) | Appendix in the DRD, WC members can send written feedback prior to the relevant WC meeting)<br><br>* **Patient version of the DRD** will be trialled during 2nd meeting of WC, including Dutch, patient version of executive summary, and additional definitions | |
| **Efficiency/sustainability** | | | |
| Framework & work process<br>* Reduce the number of **non-essential questions** as part of a pragmatic approach (M7, M4)<br>→ **Skip part 2** (selection of medicines) and reduce **part** 3 for certain medications (M12)<br>→ **Skip specific questions of part 3** if not necessary for dosing (e.g. pregnancy-induced changes in CYP enzymes) depending on medication (M4)<br>* Assess **time/cost requirement**s associated with using various versions of the framework (M7)<br><br>* Discussions of **risk-benefit analysis** should focus on dosing and **not medication use** (M7) | * Framework is long (M4, M9, M14, M15, M16, M17, M19)<br>* Comprehensive framework is necessary to make informed decisions about doses that 5ptimize both maternal and fetal risks & benefits (M3, M6, M9, M17)<br>* Background information in framework helps HCPs address all questions patients may have (M14, M17, M6)<br>* Comprehensive framework useful at this stage in order to build trust among users, especially given conservative stance with regards to prescribing medication in pregnancy (M14 M5, M20)<br>* Part 2 including information on medication use and indication(s) is very useful as background for non-HCPs (M14, M16) | **Lighter and modular version of the framework,** will be introduced during 2nd meeting of WC, and exemplified for a different medication<br><br>* Dose-specific **risk-benefit assessment** section developed for inclusion in the FDSP, will be introduced during 2nd meeting of WC | * Use **visual aid** created for risk-benefit assessment section to support WC discussions |
| Selection of medications<br>* Establish clearer **criteria for selecting** medications (M13, M3)<br><br>* Prioritize medications that are likely to require a **dose adjustment** during | * Medications that do not require a dose adjustment are also relevant for prescribers and | * Explore whether medications can be **prioritized** and/or **categorized** based on **pre-** | *Medications to be included regardless of whether a dose adjustment is required |

| | | | |
|---|---|---|---|
| pregnancy based on a preliminary search and/or PK-PD properties e.g. narrow therapeutic range (M7, M12, M20)<br><br>* **Categorize medications** based on availability of evidence (M8)<br><br>* Include medications for which **clear evidence** and/or an **evidence-based dose** is available without a full analysis using the framework (M12) | can help build trust in the approach of the MIPF → include in MIPF and state that no adjustment is needed (M15, M1, M7)<br><br>* Responds to an urgent clinical need (M7)<br>* However, it is important to familiarize WC with use of framework & to demonstrate the merits of the framework/methods to MIPF end-users as part of proof-of-concept approach (M1) | **established criteria** (at a later stage)<br><br>* Explore whether **a lighter version of the framework** can be used for certain medications based on these criteria | * Medications selected based on a) clinical need (WC can help inform the choice of medications) and b) the availability of PK evidence |
| **Credibility/usability** | | | |
| Definitions<br>* Outline definitions of **trimesters** of pregnancy and amenorrhea duration or outline time in **gestational weeks** (M4)<br>* Outline definitions for **subjective terms** e.g. likely, probable etc (M7) | *HCPs across different specialties use different/inaccurate definitions of trimesters (M4)<br><br>* May not be feasible, especially regarding fetal safety (M1, M10) | * Outline **definitions of trimesters and/or gestational weeks** whenever relevant as part of DRD; include this definition in the dose recommendation | * Discuss with Lareb whether a **banner with definitions** can be added to all relevant kennispagina's |
| Fetal safety<br>* Provide more **layered information** on fetal safety (include animal/preclinical studies, explore AUC versus Cmax approaches) (M10)<br><br>* Distinguish between **permanent & transient fetal effects** (M4)* | * Information on fetal safety including on teratogenicity is often lacking (M5, M20)<br>* Most WC members have limited expertise on how to interpret animal/reprotox studies (M11)<br>* Sustainability: do not repeat analyses already performed by Lareb on teratogenicity (M1, M21) | * Clearly distinguish between these two categories in each relevant question | * **Consult teratology expert** during DRD writing process to determine the extent to which preclinical/animal data need to be included to support the proposed dose |
| Risk-benefit analysis<br>* Risk-benefit analysis should focus on **dose adjustments** (as opposed to medication use) during pregnancy (M7) | * Identifying appropriate indications for a given medication is not the mandate of the WC; will lighten the process (sustainability) (M22, M7) | * Dose-specific **risk-benefit assessment** section, including a visual aid and | * Use **visual aid** created for risk-benefit |

| | | | |
|---|---|---|---|
| * Make a clearer distinction between **maternal & fetal considerations** for each question in the framework (M22, M5)<br>* Distinguish between **permanent & transient fetal effects** (M4)*<br>* Explore **patient preferences** and **experience** (e.g. efficacy versus side effects & therapy adherence) in more depth (M6)<br>* Make **explicit statements** on available options for dosing based on the associated maternal and fetal risks and benefits & respective **weights** assigned to these risks and benefits (M16)<br>* Create a **visual template** to inform the risk-benefit balance of the standard dose versus potentially adjusted dose in pregnancy to facilitate discussions (M12)<br><br>* Explicitly address whether the risk-benefit analysis alters the **balance between the medication** at hand and **alternative interventions** for the indication(s) under consideration (M16, M8) | * Risks-benefit analysis of use already conducted by Lareb (M21)<br><br>* Can help navigate decision-making in the presence of high uncertainty, among others regarding fetal safety (M12) | explicit statements on dosing choices, developed for inclusion in the FDSP, will be introduced during 2nd meeting of WC<br><br>Should be explicitly addressed in section 4 | assessment section to support WC discussions |
| Quality of evidence<br>* Attach a **quality grade** e.g. GRADE to dose recommendations based on the quantity/quality of available and/or researched evidence (M7, M8)*<br>* Add a **disclaimer** to dose recommendation for medications that are less well investigated (M7) | * Validity of dose not just dependent on level of evidence but on subjectively assigned weights to fetal & maternal risks & benefits (M1)* | * **Quality grading system** can be reexamined at a later stage, when the MADAM research team & WC have gained more experience with DRDs<br>* Include a short summary of the quality of available data on Lareb (?) and link to summary of Dose Rationale Document on **MELINDA website** | |
| Background information for users | * Keep the dose recommendation concise (M13) | | |

| | | | |
|---|---|---|---|
| Add a section on '**other considerations'** in dose recommendation (following example of Federatie Medisch Specialisten guidelines) including on:<br>***Rationales and assigned weights** as part of risk-benefit analysis of adjusted dose (VJ)<br>* **Information to communicate to patients** e.g. inform patients that the dose may have to be increased as a result of physiological changes as a result of pregnancy (LS, LK) | * Be mindful that any information that is included in the dose recommendation creates a precedent (M22)<br><br>On dosing considerations and assigned weights<br>* Understanding of dosing considerations is likely to increase HCPs' willingness to prescribe the medication (M15)<br>* Information on weights assigned to various arguments for dosing will help for shared-decision-making (M1)<br><br>On communication to patients<br>* Information that is communicated to parents can significantly influence effect of therapy: e.g. follow-up; therapy adherence (M7)<br>* Overbearing information, undermines the notion of HCP autonomy (HCPs are supposed to communicate with patients) (M2)<br>* Makes HCPs liable for informing their patients | * Include a short section on **arguments underpinning risk-benefit analysis** in **dose recommendation?**<br>* **Do not include** a section on **patient communication**? | |
| Quality control<br>* Include a mechanism to gain **user/prescriber feedback** on the validity of the proposed doses e.g. signaling of adverse effects (M12, M8, M22) | * Will create bias in incidence of side effects (LK, SW)<br>* Difficult to attribute side effects in a dynamic period like pregnancy (M8) | | * Use existing **feedback button** on Lareb website |
| **Access** | | | |
| HCPs<br>* Need to increase the awareness of Lareb website among general practitioners (M7) | * GPs are an important group of prescribers for pregnant women | | * Leverage advisory board members' connections to the Dutch society of GPs (**NHG)** |
| Patients<br>* Develop & publish patient information on dedicated websites e.g. thuisarts.nl | * Patients want to be informed & take decisions on medication dosing (M7, M1)<br>* Need for intelligible information for patients (M6) | * **Patient version of the DRD** will be trialled during 2nd meeting of WC, including Dutch, patient version of | * **Patient information** on model-informed dosing approach will be made available on **MELINDA website** |

| | | | |
|---|---|---|---|
| | * Main portal with information on medications for patients is apotheek.nl (Patientenfederatie, during 1st meeting of Advisory Board) | executive summary, and additional definitions | * Contact established with **apotheek.nl** via Patientenfederatie to share link to MELINDA website and/or information on MADAM and the underlying approach |

#### 2. FDSP piloted for sertraline

##### 2.a. Main document

*This framework comprises instructions for authors for each relevant question. A slightly amended version of the framework, including definitions and background information on pharmacological and medical concepts, was developed for FDSP reviewers.*

#### EXECUTIVE SUMMARY

Sertraline is a widely used antidepressant during pregnancy. The current dosing strategy for sertraline in nonpregnant adults follows a 'start low, go slow' approach, whereby a dose is titrated based on the effect on an individual patient's symptoms within a 25-200 mg range.
So far, no specific dose recommendation in pregnancy for sertraline has been issued. Changes in the distribution and metabolism of sertraline during pregnancy may justify an alteration of the standard dosing strategy followed in nonpregnant adults (see **Part 2** on the clinical and pharmacological arguments for investigating the dose of sertraline in pregnancy).

**Part 3.1.** of this document examines information on the use and safety of sertraline in pregnancy. **Part 3.2** summarizes the available knowledge on the pharmacokinetics of sertraline in nonpregnant adults. **Part 3.3.** then looks at the pharmacokinetics, dose-related efficacy and safety data of sertraline during pregnancy. The findings of **Part 3** can be summarized as follows:

- A large variation in the pharmacokinetics of sertraline (i.e. how sertraline is processed by the body, resulting in a given plasma concentration) is observed among nonpregnant individuals. This variation is most likely related to genetic variation in the multiple hepatic (CYP) enzymes responsible for metabolizing sertraline. This variation means that it is hard to define a therapeutic range (i.e. a concentration range for which the medication works with optimal safety and efficacy). There is no clear toxic threshold for sertraline (i.e. a dose or blood concentration at which it is known to cause harm) but toxicity occurs infrequently outside of pregnancy.
- It is likely that pregnancy-induced changes in maternal physiology result in an overall decrease in the maternal plasma concentration of sertraline, especially in the second and third trimesters, in comparison to pre-pregnancy levels. While the extent of this decrease is hard to quantify, it is likely to be moderate on average, with some variation across pregnant individuals.
- The latter decrease is likely to be for a large part caused by the combined effect of CYP enzyme induction (apart from CYP2C19) and a decrease in albumin concentration during pregnancy. The interindividual variability in maternal exposure to (i.e. the plasma concentration of) sertraline during pregnancy can most likely be attributed to genetic variation in CYP2C19.
- The latter information was obtained from a combination of small, moderate-quality pharmacokinetic studies (described in **Paragraph 3.3.1**) and a physiologically-based pharmacokinetic model in pregnancy (described in **Paragraph 3.3.2).**

- All in all, these pharmacokinetic data suggest that the dose of sertraline should not be reduced and may even have to be increased for certain pregnant women in the second or third trimesters of pregnancy depending on the clinical response to treatment over time.

- Our knowledge of the effect of pregnancy on the pharmacodynamics of sertraline (i.e. the mechanisms through which sertraline exerts its effects) is insufficient to inform a pregnancy-adjusted dose.
- While there is a paucity data on the fetal toxicity of sertraline in relationship to dose (none available for most fetal and neonatal outcomes), evidence of a dose relationship between the occurrence of poor neonatal adaptation syndrome (i.e. a combination of transient neurological symptoms resulting from sertraline withdrawal shortly after birth) and the use of doses of sertraline ≥ 100 mg in the third trimester of pregnancy was found. The latter dose-effect relationship may exist for other fetal or neonatal outcomes and may constitute a trade-off for dosage increase in the third trimester of pregnancy.

The latter information was used to issue the dose recommendation of sertraline in pregnancy outlined in **Part 1** of this document, which also includes questions to the Working Committee.

**Part 4** examines the feasibility and acceptability of the proposed dose. In short,

- The level of confidence in the proposed dosing strategy appears sufficient. Despite variations in the pharmacokinetics of sertraline across individuals and the lack of a well-established maternal therapeutic range as a reference for dosing, a cautious approach was adopted for issuing a pregnancy-adjusted dose (i.e. use of a conservative reference range for identifying an optimal dose through modelling). Based on the chosen approach, some pregnant women may be underdosed but maternal overdosing appears unlikely. The risk of maternal overdosing also appears minimal given our recommendation to titrate the dose based on a pregnant individual's clinical response to sertraline. One area of uncertainty concerns fetal toxicity in relationship to dose, which is most likely to be a consideration for dosing in the third trimester of pregnancy given the association between third trimester exposure and at least one (transient) neonatal outcome.
- The recommended dose strategy appears feasible as it does not significantly depart from the dose recommendation in nonpregnant adults.
- The Working Committee deems the proposed dose acceptable taking into account the direct **maternal benefits** of an appropriately treated depression and/or anxiety disorder and the indirect **fetal benefits**, including a lower risk of adverse perinatal outcomes including prematurity, intrauterine growth restriction, and a reduced likelihood of maternal stress and potentially harmful maternal behaviors. While in theory, a higher dose of sertraline could be linked to an **increased risk of fetal harm**, the absolute incidence of permanent fetal defects associated with sertraline use is very low and the recommended dose remains with the standard range for which the latter data were obtained. Overall, in light of this positive risk-benefit ratio, the Working Committee finds the proposed dose to be advisable.
- Added value: while the proposed dose does not fundamentally deviate from the dose in nonpregnant adults, it is based on a more extensive review of the available evidence as well as on an explicit discussion of the fetal and maternal benefits and risks involved which may help create more certainty and uniformity in dosing practices in pregnancy.

#### PATIENT SUMMARY

Sertraline is a medication prescribed for the treatment of depression or anxiety disorders. Sertraline users often start with a low dose that is gradually increased until an effective dosage is reached to manage symptoms. The corresponding dose is then maintained. The dose (amount) of sertraline received by pregnant women to treat their depressive and/or anxiety symptoms may sometimes be higher than outside of pregnancy.

**What are the benefits and risks of using sertraline during pregnancy?**

Sertraline can be used during pregnancy. An important benefit is that it reduces a mother's depression and anxiety symptoms. This is also likely to benefit the child because a mother's poor condition may increase the risk of preterm birth and low birth weight of the baby. Other studies have found that sertraline use may increase the risk of preterm birth and low birth weight of the baby, however it is unclear if these effects are due to sertraline, the underlying illness of the mother or other factors. Sertraline may slightly increase the risk of treatable heart defects in the child but this risk is small. However more recent studies have not found this link. Sertraline use, especially in the third trimester, increases the risk of high blood pressure in the newborn's lungs, a severe illness. However, the likelihood of this risk occurring remains very low even when sertraline is used. Sometimes, sertraline may cause short-term withdrawal symptoms in a newborn baby, for example slight shaking and crying. These symptoms usually disappear on their own within a few days after birth. It is recommended that babies whose mothers used sertraline during the third trimester of pregnancy be born in the hospital so they can be monitored for withdrawal symptoms and respiratory issues.

**Should pregnant women use a different dose than non-pregnant individuals?**

To date, there has been little research on the most suitable sertraline dose for pregnant women. Because a woman's body changes during pregnancy, she might require a different dose compared to adults who are not pregnant. We reviewed the literature to determine an optimal dosing strategy for sertraline during pregnancy. This included looking at studies using computer models that mimic changes in a pregnant woman's body and assess the suitability of different doses at various stages of pregnancy.

- The effects of sertraline for a given dose can vary between individuals. This likely stems from differences in how sertraline is processed and removed from the body, possibly due to genetic differences between people. That's why patients may use different doses of sertraline.
- There isn't a defined amount, such as a given dose or blood concentration, where sertraline becomes harmful to the user. Harmful effects are very rare outside of pregnancy, which is likely the case for pregnant women as well.
- During pregnancy, changes in a woman's body are likely to lower sertraline levels in the blood, especially in the second and third trimesters, compared to before pregnancy. Hence, the sertraline dose should generally not be decreased during pregnancy and may even need to be higher for certain pregnant women in the second or third trimester. This depends on how a pregnant woman responds to treatment over time: the dose may have to be increased because the effect on her symptoms is less than before pregnancy or insufficient if she just started treatment. Therefore, it is important to monitor a woman's symptoms of depression during pregnancy, so that the dose can be adjusted accordingly.
- Even when the dose is increased during pregnancy, it should always remain within the dosing range for non-pregnant individuals.

**Could a higher dose be harmful to the baby?**
There is little information on whether a higher sertraline dose leads to harm for the baby. Higher doses may increase the chance of side effects such as reduced birth weight and high blood pressure in the newborn's lungs. However, even when using sertraline, the likelihood of these conditions occurring is very low. Evidence has been found on a link between higher sertraline doses and withdrawal symptoms in the newborn baby. This link was only seen when sertraline is used in the third trimester. The risk of heart defects is unlikely to be linked to the dose because such defects only occur when the babies' organs are being formed in the first trimester when the dose is unlikely to be increased. Overall, the risks of increasing the dose of sertraline during pregnancy for the baby are deemed low because increased doses will remain within the normal dosing range of sertraline for which fetal safety data have been obtained.

**Could a higher dose be harmful to the pregnant woman?**
The risk for a pregnant woman to receive too much sertraline is low. There are almost never signs of harm from higher sertraline doses outside of pregnancy. This is likely to also be the case during pregnancy given that sertraline levels for a given dose tend to be lower. In addition, an 'overdose' appears unlikely because the dose is gradually increased based on the pregnant woman's symptoms.

**Risks and benefits for the proposed sertraline dosing strategy during pregnancy compared to the standard dose**

**Proposed dosing strategy:** increase the dose within the normal range if symptoms worsen during pregnancy; adjust the dose progressively based on the pregnant woman's symptoms.

| Risks for the mother | Benefits for the mother |
|---|---|
| - None expected: no harmful effects within normal dose range in users who are not pregnant ↔ | **- Better control of depression or anxiety symptoms linked to reduced likelihood of harmful behaviors and other pregnancy complications such as preterm birth** ⇑<br>**- Risk of postpartum depression*** ⇓ |
| **Risks for the baby** | **Benefits for the baby** |
| - Possible risk of reduced birth weight ⇑<br>- Low risk of high blood pressure in the lungs when using sertraline in the third trimester ⇑<br>**- Risk of short-term withdrawal symptoms at birth when sertraline is used in the third trimester** ↑ | **- Indirect benefits due to better controlled depression in the mother** ⇑<br>**- Potential improved bonding due to reduced risk of postpartum depression in the mother** ⇑ |

*Postpartum depression is a type of depression that some people experience after having a baby

↑ = likelihood demonstrably increases with dose adjustment (evidence on dose-effect relationship)
⇑ = likelihood theoretically increases with dose adjustment (no evidence on dose-effect relationship)
↓ = likelihood demonstrably decreases with dose adjustment (evidence on dose-effect relationship)
⇓ = likelihood theoretically decreases with dose adjustment (no evidence on dose-effect relationship)
↔ = likelihood unchanged with dose adjustment

**Bold: risks that carry relatively more weight**
*Green arrow: beneficial effects of dose adjustment*
*Blue arrow: neutral effects of dose adjustment*
*Red arrow: detrimental effects*

**Final decision:**
The proposed dosing strategy appears reasonable based on the following: because the possible risks of increased sertraline doses for the baby occur rarely or resolve within days, these risks are considered to weigh less than the benefits of effectively treating a pregnant woman's depression and anxiety during pregnancy, benefitting her and her child.

### TABLE OF CONTENTS

**Part 1 – Dose recommendation in pregnancy and questions to the Working Committee** **p. 18-20**
1.1. Publication page on Lareb p. 18
1.2. Proposed dose recommendation p. 18-19
1.3. Questions for discussion by the MADAM Working Committee p. 19-20

**Part 2 – Selection of medication** **p. 21-23**
2.1. Medication and selected indications p. 21
2.2. Extent of use in pregnancy p. 21
2.3. Alternative treatment interventions p. 22
2.4. Reasons for investigating dosing in pregnancy p. 22-23
***** **Conclusion part 2** **p. 23**

**Part 3 – Analysis of pharmacokinetic, efficacy and safety data** **p. 24-49**
3.1. Use and safety information in pregnancy p. 24
3.1.1. Registration for use in pregnancy p. 24
3.1.2. Safety in pregnancy p. 24-27

3.2. Pharmacokinetics and dosing considerations (general) p. 27
3.2.1. Administration route(s) p. 27
3.2.2. Formulation(s) and practical considerations for dosing p. 27
3.2.3. Recommended dose range p. 27-29
3.2.4. General pharmacokinetic properties p. 29-31
3.2.5. Metabolism and elimination pathways p. 31
3.2.6. Therapeutic range (nonpregnant population) p. 31-33
3.2.7. Interindividual variability p. 33-34
3.2.8. Underdosing or toxicity: clinical or biochemical signs p. 34-35
3.2.9. Underdosing or toxicity: use of laboratory analyses p. 35
3.2.10. Indications for pharmacogenetic testing p. 36
3.2.11. Underdosing or toxicity: margin for intervention p. 36

3.3. Pharmacokinetics, pharmacodynamics & dosing considerations (in pregnancy) p. 37
*Pharmacokinetics*
3.3.1. Pregnancy-induced changes in pharmacokinetics p. 37-38
3.3.2. Data from pharmacokinetic studies p. 38-46

*Pharmacodynamics*
3.3.3. Disease progression in pregnancy p. 46
3.3.4. Pharmacodynamics in pregnancy p. 46-47
3.3.5. Dose-related efficacy in pregnancy p. 47
3.3.6. Dose-related toxicity in pregnancy p. 47
3.3.7. Adverse events and dosing in pregnancy p. 47-48
3.3.8. Drug interactions and dosing in pregnancy p. 48
3.3.9. Pathophysiological covariates and dosing in pregnancy
3.3.10. Postpartum dose recommendation p. 48-49

3.4. Proposed dose p. 49
3.4. Can a preliminary dose recommendation be issued in pregnancy based on available data? p. 49
*****Conclusion part 3** **p. 49**

**Part 4 – Dose selection and implementation in pregnancy p. 50-58**
4.1. Feasibility p. 50
4.1.1. Feasibility of the proposed dose p. 50
4.1.2. Impact on healthcare resources and processes p. 50

4.2. Desirability p. 51
4.2.1. Level of confidence p. 51
4.2.2. Expected benefits p. 51
4.2.3. Expected risks p. 52-53
4.2.4. Risk mitigation p. 53-54
4.2.5. Residual risks after mitigation p. 54
4.2.6. Knowledge gaps p. 54-55
4.2.7. Risk-benefit analysis p. 55-56

4.2.8. Implications of findings for medication indication(s) p. 56
4.2.9. Implications of findings for medication administration p. 57
4.2.10. Added value of the proposed dose p. 57

4.3. Implementation p. 57
4.3.1. Acceptability by healthcare practitioners p. 57
4.3.2. Acceptability by pregnant women and their partners p. 58
4.3.3. Next steps for implementation p. 58

**Part 5 – References p. 59-65**

**Part 6 –Version control p. 66**

#### ABBREVIATIONS

ADME: Absorption, Distribution, Metabolism, Excretion
AUC: Area Under the Curve
CBG: College ter Beoordeling van Geneesmiddelen
Cl: Clearance
Cmax: Peak plasma concentration
Cmin: Lowest concentration of a drug
CPIC: Clinical Pharmacogenetics Implementation Consortium
CYP: Cytochrome P450
EM: Extensive Metabolizer
EMA: European Medicines Agency
F: Bioavailability
FDA: Food and Drug Administration (USA)
FDSP: Framework for Dose Selection in Pregnancy
Fss: Fraction of systemic exposure
FTK: Farmacotherapeutisch Kompas
GA: Gestational age
HCP: healthcare practitioner
IM: Intermediate Metabolizer
MADAM: Model-Adjusted Doses for All Mothers
KNMP: Koninklijke Nederlandse Maatschappij ter bevordering der Pharmacie
MIPF: Model-Informed Pregnancy Formulary
NHG: Nederlands Huisartsen Genootschap
NVOG: Nederlandse Vereniging voor Obstetrie en Gynaecologie
NVvP: Nederlandse Vereniging voor Psychiatrie
OCD: Obsessive-Compulsive Disorder
PBPK: Physiological Based Pharmacokinetic Modelling
PD: Pharmacodynamics
PDRD: Pregnancy-adjusted Dose Rationale Document
PK: Pharmacokinetics
PM: Poor Metabolizer
PNAS: Poor Neonatal Adaptation Syndrome
PP: Postpartum
PPHN: Permanent Pulmonary Hypertension of the Newborn
PTSD: Post-Traumatic Stress Disorder
SmPC: Summary for Product Characteristics
SOP: Standard Operating Procedures
SSRI: Selective Serotonin Reuptake Inhibitor
T: Trimester
T1: First Trimester
TDM: Therapeutic Drug Monitoring
TIS: Teratology Information Service
Tmax: Peak plasma time
T1/2: Half-life
UM: Ultrarapid Metabolizer
Vd: Volume of Distribution

#### PART 1 – DOSE RECOMMENDATION IN PREGNANCY & QUESTIONS TO THE WORKING COMMITTEE

❖ *Context: The following section introduces the preliminary dose recommendation outlined based on the data from parts 2 to 4.*

##### 1.1. Provide the title and hyperlink to the TIS medication page on which the dose will be published.

Newly created page on 'Sertraline dose information during pregnancy', link.

##### 1.2. Outline the proposed dose for publication on the Lareb/TIS website.

❖ *Context: The text below will be published on the aforementioned page of Lareb. NB: if a dose recommendation cannot be issued, briefly outline remaining knowledge gaps and how to address them.*

Based on the previous findings on the pharmacokinetics and pharmacodynamics of sertraline in pregnant women and nonpregnant adults, the following dose recommendation can be outlined for pregnant women:[1]

| | |
|---|---|
| **Sertraline use preceding pregnancy** | Maintain the dose that was used before pregnancy. If symptoms worsen during pregnancy, a dose increase should be considered. Similar steps to those used in non-pregnant adults can be followed in this regard. Doses exceeding 150 mg should only be prescribed following careful consideration and in consultation with the patient. |
| **De novo sertraline use in pregnancy** | Follow guidelines for initial dosing and dose titration of non-pregnant patients. If symptoms worsen, consider increasing the dose. Similar steps to those used in non-pregnant adults can be followed in this regard. Doses exceeding 150 mg should only be prescribed following careful consideration and in consultation with the patient. |

CYP2C19 polymorphisms

Pregnancy is not a standard indication for testing the CYP2C19 profile in sertraline users. If it is known or determined based on indication, the following advice can be followed: Maintain the dose used before pregnancy if the pregnant woman previously used sertraline. Be vigilant for an increase in symptoms, as reduced response may occur, particularly in the 2nd and 3rd trimesters due to increased breakdown during pregnancy.

**CYP2C19 intermediate, normal, or ultrarapid metabolizers**: Consider a dose increase if symptoms worsen. Adjust the dose as necessary based on clinical response and side effects. Follow the guidelines for initial dosage and dose titration in non-pregnant patients when starting treatment de novo.

[1] This dose recommendation fully aligns with the Dutch dose recommendation outlined in 1.2.

CYP2C19 poor metabolizers: Consider a dose increase if symptoms worsen, with a maximum dosage of 100 mg. Follow the guidelines for initial dosage and dose titration as in non-pregnant patients when starting treatment de novo, up to a maximum dosage of 100 mg

Other considerations for dosing
The Body Mass Index of a pregnant woman does not affect the required dose of sertraline.

TDM
TDM is not routinely recommended for sertraline users in the Netherlands. It may be considered on an individual basis if unexplained side effects or inadequate response are observed during pregnancy.

##### 1.3. Outline remaining questions for discussion by the MADAM Working Committee.

| **Questions** | **Feedback MADAM Working Committee** |
|---|---|
| 1 Should the dose recommendation include fewer scenarios and/or subgroups (e.g. CYP phenotypes)? | The initial dose recommendation reviewed by the WC was deemed to comprise an adequate number of scenarios, with a few caveats:<br>a) Given the relatively low incidence of CYP genotyping, the WC had differing views on whether detailed information on CYP genotyping should be included – overall, concise information was deemed useful. Intermediate metabolizers should be included.<br>b) Overall the WC was of the opinion that therapeutic drug monitoring should only be considered as a last resort for sertraline given the lack of a well- established therapeutic range and the highly limited reliance on TDM for nonpregnant sertraline users in the Netherlands. The WC did not favor using the proposed therapeutic range given the lack of consistency of underlying data and as it differed from the available ranges in nonpregnant adults and may therefore be confusing in clinical practice.<br>c) The WC felt it was important to include clear instructions on postpartum dose adjustments.<br>d) Postpartum depression should only be addressed insofar as it may prevented through appropriate treatment exposure antenatally. |
| 2 Should a maximum dose be defined? | Overall the WC held the view that the same maximum dose should be maintained during and outside of pregnancy (i.e. 200 mg). The same maximum dose should be usedfor pre-existing and de novo users of |

| | |
|---|---|
| | sertraline during pregnancy. A lower maximum dose only appeared indicated for CYP2C19 poor metabolizers. |
| 3 Should the dose recommendation include more specific information regarding the clinical follow-up of sertraline users in pregnancy? E.g. regarding the frequency of assessment and/or the nature of symptoms justifying dose adjustments | A few members of the WC suggested that it may be useful to establish clearer guidelines, particularly on the frequency of follow-up. However, it was deemed impractical to offer more precise guidelines in this regard given the diverse profiles of patients receiving sertraline, each with varying (severity of) pathologies, and the ensuing variety of settings of follow-up (general practicitioner, psychiatrist, POP-poli etc). |
| 4 To what extent should TDM be used to guide dosing in pregnancy? What are your thoughts on the proposed reference range?<br>*Given the lack of a clearly defined therapeutic range, we suggest to only rely on TDM in cases where dose adjustments based on clinical follow-up (and potentially genetic screening) do not provide the desired improvements in clinical response. Braten et al.'s proposed reference range of 10-75 ug/l appears the most suited as a reference for TDM, being based on a defined pharmacodynamic endpoint as well as being the most conservative of the available ranges (lowest chance of overdosing).* | See 1b) |
| 5 Should the dose recommendation include more detailed information on the underlying evidence and/or considerations for dosing? | The WC discussed whether to include specific recommendations on patient communication, especially regarding the need for sertraline users to advise their HCP of potential changes in their symptoms and side effects over the course of pregnancy. While the latter was considered a standard part of patient counseling, most members of the WC agreed with the inclusion of one sentence in this regard. |
| 6 Consider the wording of the dose recommendation: should (some parts of the) dose recommendations be articulated in a more stringent/flexible way? | Most members of the WC found the wording of the dose recommendation appropriate. |
| 7 Should postpartum depression be covered? | See 1d). |

#### PART 2 - DRUG SELECTION

❖ *<u>Context</u>: This section outlines the scope of the dose rationale document (which indication(s) for the medication i.e. health conditions being treated) and arguments for investigating the dose of the selected medication in pregnancy. Important arguments that ought to be considered include the extent of use, how the medication ranks compared to alternative interventions to treat the chosen indication(s), pharmacokinetic arguments (i.e.e potential changes in the maternal or fetal plasma concentration of the medication as a result of physiological changes during pregnancy) as well as clinical arguments (e.g. lack of clinical guidance on dosing in pregnancy).*

**2.1. Which drug and indication(s) are under consideration?**

Sertraline, with the following indications:

| | |
|---|---|
| **Antenatal depression** | This document focuses on the treatment of antenatal depression and the prevention of postpartum depression through appropriate pharmacological treatment during pregnancy. |
| **Anxiety disorders** | Including panic disorder, obsessive compulsive disorder, social anxiety disorder and posttraumatic stress disorder. |

The indication for starting an SSRI may be established before (de novo use) or during pregnancy (continued use).

**2.2. How often is the investigated drug used for the chosen indication(s) in pregnancy?**

<u>Prevalence of depression during pregnancy</u>

A literature review by Underwood et al. (2016) found that 17% of pregnant women in Western countries experience symptoms ranging from mood instability to major depressive disorder. In the Netherlands, it was estimated that 5.4% of pregnant women experience depressive symptoms in early pregnancy. This number reached 10.0% in late pregnancy ($p < 0.001$) (van de Loo et al. 2018).

<u>Prevalence of sertraline use in pregnancy</u>

A meta-analysis (40 studies across 15 countries) estimated that 3.0% of pregnant women use specific serotonin reuptake inhibitors (SSRIs) globally. This number was 1.6% in European countries. Sertraline was the most frequently used SSRI in pregnancy internationally (Molenaar et al., 2020). In the Netherlands, it was estimated that in 2013-2014, 2.1% of pregnant women used sertraline (Molenaar et al., 2020(2)). This number may be an **underestimation** of the number of women currently using sertraline during pregnancy according to clinical experts. Sertraline is by far the most widely used antidepressant by pregnant women in the Netherlands among others due to its limited side effects and its relative safety in pregnancy and lactation (clinical expert). Citalopram or sertraline are the preferred selective serotonin reuptake inhibitors (SSRIs) during pregnancy from a fetal safety standpoint (Lareb). Paroxetine or sertraline are the preferred SSRIs during lactation (Lareb).

**2.3. Are there alternative interventions including drugs for the same indication(s) and how does the investigated drug compare to these alternatives for the chosen indication(s)?**

SSRIs constitute a second-line treatment for depression and/or anxiety disorders. The initial treatment consists of non-pharmacological interventions including psychotherapy (Nederlands Huisartsen Genootschap (NHG) 2022, Nederlands Vereniging van Psychiatrie (NVvP) 2010). An SSRI may either be started as a second step in treatment in case of insufficient clinical response to psychotherapy and/or simultaneously with psychotherapy e.g. in the presence of severe symptoms. Non-pharmacological interventions are often preferred for treating anxiety disorders (clinical expert). During pregnancy, the relative preference for non-pharmacological interventions is greater to minimize potential fetal risks associated with SSRI use (Fumeaux et al., 2019). When starting treatment in pregnancy, non-pharmacological interventions will thus typically be initiated prior to starting an SSRI if the severity of the diagnosed symptoms allows . A SSRI treatment will only be initiated if non-pharmacological interventions (e.g. psychotherapy) fail to result in sufficient response (Fumeaux et al. 2019; clinical expert). If an SSRI was started prior to pregnancy, continuation of the SSRI during gestation is advised given the potentially greater vulnerability of pregnant women to depressive and/or anxiety symptoms antenatally and in the postpartum period (NVOG, 2012).

- Citalopram and sertraline are the preferred antidepressants in pregnancy (Lareb). Paroxetine and sertraline are the preferred medications during lactation (Nederlands Vereniging voor Obstetrie en Gynaecologie (NVOG, Lareb).

**2.4. Are there reasons to investigate the current dose in pregnancy based on the pharmacokinetics, mechanism of action and/or clinical experience with the drug?**

❖ Context: *Potential arguments include the pharmacokinetics, mechanism of action and/or clinical experience with the medication in pregnancy.*

Pharmacokinetics

- Both the Farmacotherapeutisch Kompas (FK) and Lareb indicate that due to pharmacokinetic changes in pregnancy, the plasma concentration of sertraline may be reduced in the second and especially the third trimester of pregnancy. According to the FK, this may require a dose increase that is not further specified.
- Sertraline has a complex metabolism involving five polymorphic[2] CYP enzymes whose activity is influenced in different ways by pregnancy (Huddart et al. 2020)
- Sertraline is highly protein bound, meaning that changes in the concentration of plasma proteins in pregnancy may also affect the unbound fraction of sertraline (Soma-Pillay et al. 2016). The overall impact of these pregnancy-induced changes on the pharmacokinetics of sertraline (that is, the maternal plasma concentration of sertraline) is unclear (Pariente et al. 2016; Koren et al. 2018).

Clinical practice

Sertraline is either prescribed by general practitioners (GPs) (first line) or psychiatrists (second line) in the Netherlands. Data from focus groups

[2] *Genetically variable.*

among pregnant women and HCPs in the Netherlands show that SSRI treatment is often interrupted by women before or during pregnancy, either on their own initiative or in some cases, in consultation with their GP, out of concern for fetal safety. In certain cases, the latter decision may not align with clinical guidance that supports the use of SSRIs in pregnancy in clinically relevant cases of depression and/or anxiety disorders as a second-line intervention in combination with non-pharmacological treatment (NVOG, 2012; NVvP 2010). Furthermore, clinical experience reveals that HCPs in both first-line and second-line care tend to favor lower(ing) the dose of sertraline that they prescribe to women during pregnancy, often prescribing doses within a 25-100 mg range (compared to a recommended range of 25-200 mg in nonpregnant adults). An in depth review of the evidence on the pharmacokinetics, efficacy and safety of sertraline during pregnancy can help determine whether this conservative use and dosing of sertraline in pregnant women is justified and still leads to an effective treatment of depression or anxiety symptoms during pregnancy.

##### Conclusion of part 2

Investigating the adequacy of the current dosing strategy of sertraline in pregnancy appears justified based on the following:

- Clinical arguments: frequently used medication in pregnancy with maternal and fetal benefits in clinically relevant cases of depression or anxiety, lack of pregnancy-specific dosing guidance
- Pharmacological arguments: pregnancy-induced changes in sertraline distribution and metabolism with a likely effect on maternal plasma exposure (i.e. the maternal plasma concentration)

#### PART 3 – ANALYSIS of PHARMACOKINETIC, EFFICACY & SAFETY DATA

❖ *Context: In the following section, more detailed information on the administration, pharmacokinetics, efficacy and safety of the medication in pregnancy is analyzed to inform dose selection in pregnant women and their unborn children.*

#### Part 3.1 – Use and safety information

##### 3.1.1. Is the drug registered for use in pregnancy for the selected indication(s)?

Sertraline is not registered for use in pregnancy for the management of depression and/or anxiety disorders in the Netherlands, the European Union or the USA.

##### 3.1.2. What is the safety assessment of the drug in pregnancy?

❖ *Context: safety data was obtained from secondary sources (Lareb teratology information and relevant clinical guidelines), along with ad hoc literature searches. The table includes absolute and relative risks of each outcome occurring for exposure to the drug compared to the baseline risk (which should also be specified). Pregnancy trimesters were included whenever known. Whenever dose-related information could be found regarding the incidence of a given outcome, the latter information was included in the table below. Studies on dose-related toxicity are also discussed in 3.3.6.*

Safety profile of sertraline in pregnancy

*CI 95%: 95% confidence interval; MA: meta-analysis, NVOG: Nederlandse Vereniging voor Obstetrie en Gynaecologie, OR: odds ratio, PNAS: poor neonatal adaptation syndrome; PPHN: permanent pulmonary hypertension of the newborn, RR: relative risk, SSRI: selective serotonin reuptake inhibitor.*

| | Pregnancy outcomes | Perinatal outcomes | Neonatal outcomes | Congenital malformations |
|---|---|---|---|---|
| **NVOG (2012)** | There is no reason to advise against SSRI use based on the risk of pregnancy complications.<br><br>There appears to be no reason to discontinue SSRIs before or during pregnancy due to a potential risk of | There are no indications to advise against SSRI use based on the risk of childbirth complications.<br><br>There are no indications of an association between SSRI use and the occurrence of spontaneous miscarriages, vaginal artificial deliveries, cesarean | SSRI use is associated with permanent pulmonary hypertension of the newborn (**PPHN)** (association is not found in all studies). PPHN leads to persistent high pulmonary vascular resistance after birth, causing abnormal blood flow from the right to the left side of the fetal heart via fetal pathways. This can result in severe hypoxemia unresponsive to standard respiratory support. The estimate prevalence is 1.6 cases per 1000 live births, with a 7-10 % mortality rate in developed countries (Walsh-Sukys et al. 2000). SSRI exposure <20 weeks GA shows an OR of 0.80 (95% CI, 0.51-1.25), | There is no ground for advising against the use of SSRIs in standard doses or to switch to another SSRI during pregnancy based on the risk of **congenital abnormalities.**<br><br>When using SSRIs during pregnancy, the long-term effects in the neonate have not been sufficiently studied.§ |

| | | | |
|---|---|---|---|
| | miscarriage. | section and postpartum hemorrhage**.** (Nakhai-Pour et al., 2010), (Kulin et al., 1998), (Malm et al., 2005), (Salkeld et al., 2008), (Sivojelezova et al., 2005)<br><br>**There were no conclusive findings regarding a potential association between small-for-gestational use and SSRI use.** | suggesting a lower but not significantly different risk of developing PPHN. However, SSRI exposure > 20 weeks GA had an OR of 2.01 (95% CI, 1.32-3.05), indicating a notably higher risk of PPHN.The absolute risk difference for PPHN after SSRI exposure in late pregnancy was 2.6 per 1000 infants compared to 1.3 per 1000 unexposed infants (95% CI, 0.2-2.4) (Munk-Olsen et al. 2021).<br><br>→ The risk of PPHN is an indication for hospital delivery; it is recommended to observe newborns for at least 12 hours under the responsibility of the paediatrician<br><br>SSRI use is associated with **moderate neonatal adaptation** (i.e. a combination of symptoms including breathing difficulties, tremors, hypotonia, gastrointestinal disorders and sleep disorders that result from prenatal SSRI exposure and lead to withdrawal symptoms after birth. Neonatal adaptation affects **25-30% neonates** exposed to SSRIs, especially when exposed in the 3rd trimester. These symptoms largely resolve spontaneously. (Levinson-Castiel, 2006)<br><br>→ Newborns should be observed for the three first days postpartum given the risk of neonatal adaptation (either at hospital or at home) |

| | | | | |
|---|---|---|---|---|
| **Lareb** (last updated 2023) | It is not possible to issue a statement about the association between sertraline and the risk of spontaneous abortion, preterm birth, low birth weight and pre- and postnatal death effects based on available studies. | Several studies have been done on the use of SSRIs and a possible increased risk of **postpartum bleeding**. The results are not unequivocal. Since spontaneous bleeding is a known side effect of SSRIs, an increased risk of postpartum bleeding cannot be excluded. | Permanent pulmonary hypertension of the newborn (**PPHN)** in the newborn child may occur with the use of SSRIs. Observe the child after birth for signs of this, such as blue discoloration and breathing problems.<br>After prolonged use of antidepressants until delivery, the newborn child may develop toxic or withdrawal symptoms **(= poor neonatal adaptation syndrome)**. In the first days after birth, the child may experience irritability, increased muscle tension, trembling, irregular breathing, poor drinking and loud crying. | A slightly increased risk of **specific (heart) abnormalities** with SSRI use in the first trimester cannot be ruled out.[3] |
| **Other sources** (Reprotox [4] & additional ad hoc literature searches) | Desaunay et al. 2023[5] (systematic review of meta-analyses)<br>Sertraline is associated with an increased risk (small RRs) for preterm birth and low birth weight. | No information with regards to the incidence of or a dose relationship for sertraline use and the occurrence of postpartum bleedings or dysmaturity could be found. | Masarwa et al. 2019 (meta-analysis, 156,978 women): risk of **PPHN** significantly increased for SSRIs in any trimester (OR 1.82, CI 95% 1.3-2.5) and especially in the third trimester (OR 2.08, CI 95% 1.4-3.0) compared to baseline risk. Sertraline was ranked most likely to have the lowest risk for PPHN among SSRIs (P = 0.83)<br>Reprotox<br>The FDA has reviewed the additional new study results (Andrade et al. 2009, Wilson et al. 2011) and has concluded that, given the conflicting results from different studies, it is premature to reach any conclusion about a possible link between SSRI use in pregnancy and **PPHN**. | Gao et al. 2018 (systematic review, 9,085,954 births): sertraline was associated with **septal defects[6]** (RR 2.69, 95% CI 1.76-4.10), **atrial septal defects** (RR 2.07, 95% CI 1.26-3.39), and **respiratory system defects** (RR 2.65, 95% CI 1.32-5.32).<br>Myles et al. 2013 (meta-analysis, 2,609,469 women): no increase in cardiac malformations for sertraline. |

[3] Given the occurrence of heart abnormalities as part of impaired organ genesis in the first trimester, the latter risk is unlikely to be associated with dose increases that would most likely be considered in later trimesters based on the proposed dosing strategy in pregnancy.

[4] Reprotox is a US-based secondary information resource providing data on reproductive and developmental effects of chemicals and substances on humans and animals.

[5] Desaunay et al. 2023 is a systematic review of meta-analyses, which includes 51 meta-analyses investigating the risks of SSRI use in pregnancy.

[6] Septal defects occur in approximately 5 per 1000 live births (baseline risk). Atrial septal defects occur in 1-2 per 1000 live births. The outcomes after surgical closure are excellent and comparable with age-matched controls (Wu et al. 2020).

| | | | | |
|---|---|---|---|---|
| | | | Brumbauch et al. 2023 (retrospective cohort study, n = 471 mother-infant pairs)<br>- Incidence of poor **neonatal adaptation syndrome** (PNAS) in sertraline users: 3.2%<br>- RR of **PNAS** for high dose ($\geq$ 100 mg) compared to standard dose (50 mg) sertraline monotherapy in the third trimester: 3.64 (95% CI 1.27-10.47, p = 0.07) | Wang et al. 2015 (meta-analysis, 1,996,519 women): no association between cardiovascular defects and sertraline exposure.<br><br>Pedersen et al. 2009 (registry study, 493,113 children): sertraline is not expected to increase the risk of congenital anomalies.<br><br>Desaunay et al. 2023 Three meta-analyses found statistically sign increased risk of congenital heart defects |

#### Part 3.2. Pharmacokinetics and dosing considerations (general)

❖ *Context: this section focuses on the general pharmacokinetic characteristics of the medication and on how the latter characteristics inform the current dosing strategy for the medication. This section focuses on the characteristics of the medication independently of pregnancy. Pharmacokinetic changes in pregnancy, that is, physiologically-induced changes in the pharmacokinetics of the medication as a result of pregnancy are described in section 3.3.*

**3.2.1. What are administration route(s) for the chosen indication(s)? :** oral

**3.2.2. Which formulation(s) of the drug is(are) used for the chosen indication(s)? What are practical considerations for dosing?**

In the Netherlands, sertraline is available in the following forms:

| | |
|---|---|
| Sertraline hydrochloride tablets | 25 mg, 50 mg and 100 mg |
| Sertraline hydrochloride oral solutions | 20 mg/mL (for patients with difficulty swallowing) |

**3.2.3. What is the recommended dose (range) for the chosen indication(s)?**

Nonpregnant adults

The recommended dose range is 50-200 mg for depression and 25-200 mg for anxiety disorders (as summarized in the **table** below). Given the high interindividual variability in the dose-exposure-response to sertraline, doses should be titrated individually based on clinical response and side effects

(NVvP, 2010). It is recommended to start with a low dose - either 25-50 mg for anxiety disorders or 50 mg for depression and to gradually increase the dose to up to 200 mg if symptom remission is insufficient and if side effects remain tolerable (FTK). The first dose evaluation takes place 4-6 weeks upon treatment initiation. Dose increments of 50 mg can then be made on a weekly basis.

Recommended dosing ranges and strategies for sertraline in nonpregnant adults

*CBG = College ter Beoordeling van Geneesmiddelen, EMA = European Medicines Agency, FDA = (US) Food and Drug Administration, FK = Farmacotherapeutisch Kompas, KNMP = Koninklijke Nederlandse Maatschappij ter bevordering der Pharmacie (Pharmacists association), OCD = Obsessive-Compulsive Disorder, PTSS = Post-Traumatic Stress Disorder, SmPC = Summary of Product Characteristics*

| | |
|---|---|
| FK | Depression: 1 dd 50-200 mg (start with 50 mg and increase in increments +50 mg weekly as needed based on response)<br>General or social anxiety disorder, OCD, PTSS: 1 dd 25-200 mg |
| **SmPCs** | **CBG/EMA**<br>**Depression:** 1 dd 50-200 mg (increments +50 mg weekly as needed)<br>**General or social anxiety disorder, PTSS:** start treatment 1 dd 25 mg, increase to 1 dd 50 mg after 1 week (increments +50 mg weekly as needed), max. 200 mg<br>**FDA**<br>**Depression**: 1 dd 50-200 mg (increments +50 mg weekly as needed)<br>**General or social anxiety disorder, PTSS**: start treatment 1 dd 25 mg (increments +50 mg weekly as needed) |
| **KNMP** | **Depression:** 1 dd 50-200 mg (increments +50 mg weekly as needed)<br>**General or social anxiety disorder, OCD, PTSS:** start treatment 1 dd 25 mg, increase 1 dd 50 mg after 1 week (increments +50 mg weekly as needed, max 1 dd 200 mg) |
| **Lexicomp** | **Depression:** 1 dd 50 mg<br>**General anxiety disorder:** 1 dd 25-200 mg |
| **Uptodate** | **Depression:** 1 dd 50-200 mg; doses of up to 300 mg a day reported (increments +25-50 mg weekly as needed)<br>**Panic/PTSS disorder:** 25-200 mg daily |

Pregnancy

| | |
|---|---|
| Lareb | Due to the changing pharmacokinetics during pregnancy, a dose increase may be necessary. This is because in the second and especially the third trimester, plasma levels of SSRIs can decrease. |
| **FTK** | In the 2nd and particularly the 3rd trimester, plasma levels can decrease, and a dose increase may be necessary. |

##### 3.2.4. What is known about the absorption, distribution, metabolism, and elimination (ADME) of the drug?

General pharmacokinetic properties of sertraline

*AUC = Area under the curve, C/D = Concentration-dose ratio, Cmax = Peak concentration, Cl = Clearance, Cl/F = Apparent oral clearance, CL/Fss = ratio of clearance to the fraction of drug absorbed, P = p-value, PM = Poor metabolizer, T = Trimester, Vd/F = Volume of distribution/bioavailability*

| | |
|---|---|
| **Absorption** | F: 44%<br>Tmax: 4.5-8.4 hours after ingestion<br>Cmax: 20 to 55 µg/L for 1 dd 50-200 mg for 2 weeks) |
| **Distribution** | **Vd:** 20L/kg<br>**Protein binding:** 98% (predominantly albumin) |
| **Metabolism** | Complex hepatic metabolism = main elimination route<br>→ **CYP2B6, CYP2C19, CYP3A4, CYP2D6, CYP2C9**, UGT1A1, P-gp (ABCB1)*<br>Sertraline → N-desmethylsertraline |
| **Elimination** | **Clearance:** 1.09 ± 0.38 L/h/kg - 1.35 ± 0.67 L/h/kg for 200 mg dose<br>**T1/2:** 26 hours (N-desmethylsertraline 62-104 hours) |

The disposition of sertraline is primarily dependent on hepatic metabolism. Sertraline has a complex hepatic metabolism, being transformed into desmethylsertraline by five different CYP enzymes, namely CYP2B6, CYP2C9, CYP2C19, CYP2D6 and CYP3A4, all of which are characterized by the presence of polymorphic (i.e. genetic) variants with various levels of metabolic activity.

Although findings on the respective contribution of each enzyme to the metabolism of sertraline vary across studies (as described in the table below), none of the five enzymes listed above appears to be responsible for more than 30-40% of sertraline metabolism in vitro. The respective contribution of each enzyme to the metabolism of sertraline is likely to be dependent on several factors, including sertraline concentration and the particular combination of genetic variants of each CYP enzyme in a given individual (Greenblatt et al., 2001; Wang et al. 2001). At high in vitro sertraline concentrations, the metabolism of sertraline is primarily driven by CYP2C9, CYP3A4 and CYP2C19 while at lower sertraline concentrations, CYP2D6 and CYP2B6 play a more prominent role in desmethylsertraline formation (Huddart et al. 2020).

Respective contributions of CYP enzymes to the hepatic metabolism of sertraline

| | Polymorphic | Contribution to sertraline metabolism via N-demethylation | | | | |
|---|---|---|---|---|---|---|
| | | **Obach et al., 2005** | **Kobayashi et al., 1999** | **Greenblatt et al., 1999** | **Xu et al., 1999** | **George et al. 2020 (GA = 0 weeks)** |
| | | *In vitro study in human liver microsomes* | *In vitro study in human liver microsomes* | *In vitro study in liver microsomes (1 immunologic & activity measures 2 incubation with specific CYP inhibitors)* | *In vitro study of sertraline kinetics in liver microsomes of CYP2C19 EMs and PMs through specific inhibition* | *In vitro in vivo extrapolation from Obach et al. (for PBPK modelling)* |
| **CYP2B6** | Yes | 40% | 14% | 1% | | 9% |
| **CYP3A(4)** | Yes | 15% | 9% | 17-20% | | 73% |
| **CYP2C19** | Yes (30+ alleles) | 17% | 13% | 15-14% | High affinity N-demethylation of sertraline | 7% |
| **CYP2D6** | Yes | 16% | 35% | 15-5% | | 3% |
| **CYP2C9** | Yes | 14% | 29% | 17-21% | Low affinity N-demethylation of sertraline | 8% |
| **Additional findings** | | | | * Enzyme contribution is dependent on sertraline concentration | | |

While the hepatic metabolism is dependent on multiple CYP enzymes, CYP2C19 has been identified as a major metabolic pathway for the medication due to the influence of genetic variations in CYP2C19 on the pharmacokinetics of sertraline (Hicks et al., 2015). Several studies have shown major differences in the dose-adjusted plasma concentrations of sertraline in individuals with differing CYP2C19 genotypes, suggesting that CYP2C19 polymorphisms are for a large part responsible for interindividual differences in the pharmacokinetics and/or effect of sertraline (Wang et al. 2001; Milosavljevic et al. 2020). The concentration of sertraline in CYP2C19 poor metabolizers (PMs) in particular is estimated to be approximately 50% higher than in normal metabolizers (NMs) among nonpregnant adults (FK). No such findings were made for other enzymes involved in sertraline

metabolism e.g. CYP2D6 (Huddart et al. 2020).
The incidence of various CYP2C19 phenotypes in various populations is described below.

<u>Incidence of different CYP2C19 metabolisms</u>

**Clinical Pharmacogenetics Implementation Consortium (2015)**[7]

| Likely phenotype |
|---|
| Ultrarapid metabolizer (~5–30% of patients)[d] |
| Extensive metabolizer (~35–50% of patients) |
| Intermediate metabolizer (~18–45% of patients) |
| Poor metabolizer (~2–15% of patients) |

**Nederlandse Vereniging van Psychiatrie 2020**

| | CYP2C19 | | | |
|---|---|---|---|---|
| | PM (%) | IM (%) [1] | EM (%) [1] | UM (%) |
| **Kaukasisch** | 2-4 | 15-20 | 69-80 | 3-7 |
| **Aziatisch** | 10-25 | 30-50 | 25-60 | 0–0.2 |
| **Afrikaans** | 1-5 | 10-20 | 72-86 | 3 |

**3.2.5. Does the drug have an active metabolite? If so, describe its pharmacokinetic and pharmacodynamic properties.**

Sertraline has a metabolite, N-desmethylsertraline. N-demethylsetraline has a half-life of 56-120 hours (2 to 4 times longer than sertraline) but only 5-10% of the potency of sertraline. It is considered to have a negligible clinical effect (Huddart et al. 2020; Sprouse et al. 1996).

**3.2.6. Is there a therapeutic range for the chosen indication(s) of the drug and how was it determined? In the absence of such a range, how was the dose determined and what is known about a dose-exposure-response relationship?**

While some degree of association has been found between the dose and response to sertraline in nonpregnant adults, no clear therapeutic range can be defined in this population (Hiemke et al. 2018, Huddart et al. 2020). Concentration ranges described in the literature mostly derive from routine data from therapeutic drug monitoring (TDM) and a limited number of pharmacokinetic studies in nonpregnant adults as opposed to well-substantiated pharmacokinetic-pharmacodynamic endpoints.

Two proposed 'reference' concentration ranges have been described and are frequently referred to in the literature:

▪ **10-150 ug/l:** defined as a reference range by the Arbeitsgemeinschaft für Neuropsychopharmakologie und Pharmakopsychiatrie (**AGNP**) based on a review of pharmacokinetic studies of sertraline in which no toxicity was reported (primary studies in the <u>table below</u>) (Hiemke et al. 2018).

<u>Overview of primary studies cited to support the AGNP reference range for sertraline</u>

[7] *The following numbers were measured in a Caucasian population.*

| Studies | Study design | Study population | Doses used | Concentrations measured | Toxicity |
|---|---|---|---|---|---|
| Gupta et al. 1994 | Retrospective TDM study | 27 patients (not further specified) | 100-300 mg | 20-309 ug/l<br>Dose 100 mg (n = 8), 20-48 ug/L<br>Dose 150 mg (n = 3) 73-103 ug/L<br>Dose 200 mg (n = 10) 40-187 ug/L<br>Dose 250 mg (n = 2) 56-133 ug/L<br>Dose 300 mg (n = 4) 99-309 ug/L | None reported |
| Reis et al. 2009 | Retrospective TDM study | 3869 outpatients (66% women, median 43 (8-98) years) | 25-300 mg | 6-56 ug/l | None reported |

Additional pharmacokinetic studies of sertraline in nonpregnant adults

*CI: confidence interval, IM: intermediate metabolizer, NM: normal metabolizer, PM: poor metabolizer, SD: standard deviation, TDM: therapeutic drug monitoring, UM: ultrarapid metabolizer*

| Studies | Study design | Study population | Doses used | Concentrations measured | Toxicity reported |
|---|---|---|---|---|---|
| **Braten et al. 2020** | Retrospective TDM study | 1202 outpatients (69.3% women, | CYP2C19 genotypes (mean (SD)):<br>UM: 106.4 (52.0)<br>NM: 108.2 (53.4)<br>IM: 97.0 (49.4)<br>PM: 113.4 (49.8) | Sertraline (nM) harmonized to 100 mg/day (95% CI):<br>UM: 92.7 (85.1-101.1)<br>NM: 103.6 (91.8-116.8)<br>IM: 142.9 (130.8-156.2)<br>PM: 277.4 (224.1-343.5) | None |
| **Hong Ng et al. 2006** | Pharmacokinetic study | 15 outpatients (53% women, 43.3 ± 10.0 years) | 50 mg at week 1, mean 89 mg at week 6 | Week 1: 11.19 ± 1.09 ng/mL<br>Week 6: 18.47 ± 1.89 ng/mL | None |
| **Lundmark et al. 2000** | Retrospective TDM study | 319 patients | 25-200 mg<br>(mean 88 mg) | Steady state concentrations (median and range) in nmol/L:<br>25 mg: 23.0 (10.0-175.0)<br>50 mg: 40.0 (10.0-440.0)<br>75 mg: 59.0 (20.0-158.0)<br>100 mg: 66.0 (10.0-355.0)<br>125 mg: 115.0 (63.0-224.0)<br>150 mg: 127.0 (25.0-472.0)<br>200 mg: 93.0 (30.0-267.0) | None |

| | | | | | |
|---|---|---|---|---|---|
| **Ronfeld et al. 1997** | Pharmacokinetic study | 14 healthy female volunteers (mean 34.4 (20-45) years) | 200 mg (titrated with 50 mg over a 9-day interval) | Day 21: 106.6 ± 60.5 ug/L<br>Day 23: 107.9 ± 63.8 ug/L<br>Day 25: 103.6 ± 58.3 ug/L<br>Day 27: 104.7 ± 54.1 ug/L<br>Day 29: 104.2 ± 57.6 ug/L<br>Day 30: 106.9 ± 56.8 ug/L | None |
| **Unterecker et al. 2012** | Retrospective TDM study | 130 outpatients (58.4% women, 45 ± 18 years) | 111 ± 51 (25–300) mg | Dose-corrected serum level: 0.34 ± 0.23 (0.02–1.20) (ng/mL)(mg/day) | None |
| **Unterecker et al. 2013** | | 77 outpatients (58% women) | 25- 100 mg (mean 111+ SD 51 mg) | 1-136 ug/l (mean 34, SD 23) | None |

▪ **10-75 ug/l**: proposed by **Braten et al. (2020)** as an alternative reference range based on a DASB positron emission tomography study by Meyer et al. (2004) that measured 80% occupancy of serotonin transporters at 10 ng/ml sertraline plasma concentration. Based on this pharmacodynamic finding supporting the lower boundary of the AGNP's proposed therapeutic range, Braten et al. hypothesized that maximal sertraline transporter occupancy was likely to be reached at a concentration much lower than 150 ug/l, proposing 75 ug/l as an upper bound for a therapeutic window.

A third reference range is described by the Dutch TDM reference **TDM-monografie.org:**
▪ **50-300 ug/l:** based on habitual concentrations observed for the daily dose range 50-200 mg, 300 ug/l being described as a potentially toxic value, (no reference provided).

Data interpretation

While it remains hard to establish a therapeutic range for sertraline, all proposed ranges appear relatively wide. Pharmacokinetic studies demonstrate that a large majority of patients using standard doses of sertraline (50-200 mg) are well within the reference range proposed by the AGNP (10-150 ug/l) and for a large part within the more conservative range proposed by Braten et al. (10-75 ug/l). No toxicity was reported in the above studies even in individuals with plasma concentrations of sertraline beyond this range.

##### 3.2.7. How much interindividual variability is there in the dose-exposure-response?

Gupta et al. 1994 reported up to 15-fold variation in the steady-state sertraline plasma concentrations of nonpregnant patients receiving 50-150 mg daily doses of sertraline. Lundmark et al. (2000) measured an 88-fold variation in sertraline dose-adjusted concentrations obtained through routine TDM measurements in over 300 nonpregnant outpatients receiving daily doses of 25-250 mg. More information on the design of both studies can be found in the tables of 3.3.3.

It is likely that part of this variation can be explained by genetic variation in the CYP enzymes involved in the metabolism of sertraline, especially CYP2C19 (Braten et al., 2020, Huddart et al. 2020).

##### 3.2.8. Are there clinical or biochemical signs associated with underdosing or toxicity?

Underdosing in nonpregnant adults

In the absence of a clear dose-exposure relationship, sertraline underdosing can primarily be detected as an insufficient or diminished clinical response to treatment of a patient's depression and/or anxiety symptoms. Given the lack of routinely used clinical scales to monitor treatment efficacy for depression or anxiety disorders, the detection of inadequate or diminished response is dependent on anamnesis and/or self-reporting by the patient. Importantly, an insufficient response may not (solely) result from sertraline underdosing and may be determined by a variety of factors including the effect of other non-pharmacological interventions generally used in combination with an SSRI.

Overdosing in nonpregnant adults

While sertraline overdoses may in some cases be detected based on the occurrence of multiple and/or severe side effects, no clear sertraline toxicity threshold (either a dose or a plasma concentration) has been defined for nonpregnant adults (Toxicologie.org). Sertraline overdoses, defined as a daily dose >200 mg, are generally 'well tolerated' (Singh et al. 2023; clinical expert). One particular complication that may theoretically result from sertraline overdoses is serotonin syndrome, defined as a combination of neurological symptoms including cognitive, autonomic and somatic effects due to the overactivation of serotonin receptors. The main risk factor for serotonin syndrome is the use of several serotonergic medications. Clinical experience shows that the latter syndrome occurs very rarely, that causality is often hard to establish, and that the associated symptoms are generally self-limiting. In a prospective study across five US regional poison centres, Lau et al. (1996) describe 17 cases of isolated sertraline overdoses (doses up to 8000 mg, mean ingested dose 1579 mg) of which >50% were asymptomatic. Symptomatic overdoses mainly involved neurological symptoms (nausea, tremor, lethargy and rarely agitation, confusion or vomiting). In a review of sertraline overdosing admissions (> 200 mg daily dose) in a tertiary Australian referral centre, Isbister et al. (2004) reported that 20% of cases resulted in serotonin syndrome following the Sternbach criteria and/or as diagnosed by a clinical toxicologist. Other reported effects were rare and included seizures (2%), coma (3%) and transient cardiovascular effects including prolonged QTc (see reported incidence of symptoms summarized in the two tables below).

Characteristics of studies on sertraline overdosing:

| **Studies** | **Study design** | **Study population** | **Doses used** | **Toxicity reported** |
|---|---|---|---|---|

| | | | | |
|---|---|---|---|---|
| Isbister et al. 2004 | TDM database study | 156 cases (for sertraline) | 24 (16-30) daily dose | Sertraline overdose is not significantly associated with QTc-prolongation. |
| Lau et al. 1996 | Prospective observational study | 17 sertraline-only overdoses and 23 combined overdoses | The mean amount of sertraline stated to have been ingested was 1600 mg; the median ingestion was 1400 mg (range 50 to 8,000 mg) | Of the 40 presumed ingestions, 17 were isolated sertraline ingestions. Of this subgroup of isolated ingestions, ten patients (59%) were asymptomatic. Of the seven patients who had symptoms, only tremor was commonly reported (four of seven subjects). |
| Singh et al. 2022 | Literature review | - | - | Sertraline can prolong the QT interval; however, the prolongation is dose-dependent and is very modest. Sertraline may rarely produce symptoms of serotonin syndrome, though this generally happens when combining it with another serotonergic medication. |

Incidence of a) neurotoxicity and b) cardiovascular effects in sertraline overdoses (from Isbister et al., 2004)

a)

| | |
|---|---|
| No. of cases (N) | 156 |
| Seizures | 3 (2%) |
| Coma | 4 (3%) |
| Serotonin syndrome | 31 (20%) |

b)

| | |
|---|---|
| No. of cases (N) | 103 |
| Arrhythmia/conduction block* | 0 |
| Bradycardia | 8 (8%) |
| Tachycardia | 20 (19%) |
| Hypotension | 0 |
| QTc >440 msec | 41 (40%) |
| QTc >500 msec | 6 (6%) |

*Excluding bradycardia and tachycardia.

##### 3.2.9. Are laboratory analyses (e.g., therapeutic drug monitoring) routinely used to detect underdosing or toxicity?

❖ *Context: this paragraph describes whether laboratory analyses are routinely used for detecting underdosing or toxicity in the Netherlands.*

In the absence of a clearly defined therapeutic range, clinical guidance on the utility of TDM of sertraline diverges. The German Society of Neuropsychopharmacology (AGNP) recommends therapeutic monitoring of sertraline based on data from steady-state pharmacokinetic studies (Hiemke et al., 2018). The Dutch website TDM monografie does not advise TDM for sertraline apart from a number of selected indications including pregnancy ( reference concentration range: 50-300 ug/l). In practice, TDM of sertraline is rarely performed in the Netherlands (clinical expert).

##### 3.2.10. Is pharmacogenetic testing used for dosing the drug?

❖ *Context: this paragraph describes whether pharmacogenetic is routinely used for detecting underdosing or toxicity in the Netherlands.*

In the Netherlands, pharmacogenetic testing for CYP2C19 variants is advised for sertraline users that experience unexplained and/or severe side effects, especially in the presence of a history of side effects with CYP2C19-metabolized medications (NVvP, 2020). Dosing recommendations have been issued for the various CYP2C19 phenotypes, with several international clinical guidelines advising to lower the starting dose for CYP2C19 poor metabolizers (PMs) as summarized in the table below.

Dose recommendations for CYP2C19 phenotypes (nonpregnant adults)

| | Poor metabolizer | Intermediate, normal metabolizer | Ultrarapid metabolizer |
|---|---|---|---|
| **KNMP (2020)** | **Max. 75** mg daily | Start with standard dose | Start with standard dose |
| **TDM monografie** | **Max 50 mg** daily + monitoring | Max 100 mg daily + monitoring | Plasma concentration can be decreased, not clinically relevant |
| **FTK** | Adjust dose as needed in consultation with pharmacist in case of CYP2C19 polymorphism. | | |
| **CPIC** | Consider a **50% dose reduction** or switch to an alternative SSRI | Start with standard dose | Start with standard dose |

##### 3.2.11. What is the margin for intervention in the event of underdosing and toxicity?

❖ *Context: this paragraph describes the extent to which underdosing and toxicity can be prevented by acting upon the information that was collected using the strategies described in 3.2.5 to 3.2.7.*

The risk of **underdosing** can primarily be minimized by monitoring the clinical response to sertraline through a regular follow-up. If underdosing is suspected, the dose of sertraline can be increased on a weekly basis using +25 mg or 50 mg increments.

The risk of **overdosing** in nonpregnant adults can be minimized by not exceeding the daily recommended dose of sertraline and by monitoring the occurrence of side effects through regular follow-up. In the presence of (excessive) side effects, the dose of sertraline can be decreased on a weekly basis using steps of up to 50 mg weekly (KNMP 2018).

#### 3.3. Pharmacokinetics, pharmacodynamics and dosing considerations (in pregnancy)

❖ *Context: in this section we examine the available data on pregnancy-induced changes in the **pharmacokinetics** (i.e. ADME processes and resulting plasma concentration) of a medication and hence potential dosing needs during pregnancy. This section also summarizes the available data on how the **pharmacodynamics** of the medication (i.e. the medication's effect as determined by its ability to interact with its target receptors in the body) are influenced by pregnancy.*

#### Pharmacokinetics

##### 3.3.1. What are the expected effects of pregnancy-induced changes in maternal physiology, placental transfer or metabolism, and fetal metabolism on the pharmacokinetics of the drug?

The main physiological changes with a potential influence on the pharmacokinetics of sertraline during pregnancy are:

- **Changes in hepatic metabolism**: induction/inhibition of the various CYP enzymes involved in sertraline metabolism resulting in changes in the clearance and hence in the plasma levels of sertraline during pregnancy. This includes a potential influence of pregnancy on different CYP enzyme phenotypes and potential phenoconversion (= a change in phenotype, e.g. from normal metabolizer to poor metabolizer) as a result of pregnancy. The latter changes are of particular importance for CYP2C19 as a major pathway for sertraline metabolism (Hicks et al., 2015; Poweleit et al., 2022).
- **Decrease in albumin**: given the high protein binding of sertraline, a decrease in the concentration of albumin as a result of haemodilution and increased glomerular capillary leak during pregnancy may result in a higher unbound fraction of sertraline, hence increasing the volume of distribution and reducing sertraline plasma levels (Soma-Pillay, 2016). No sertraline-specific data are available in this regard.

Placental transfer of sertraline measured after delivery

*CB/MP: cord blood/maternal plasma ratio, IP/MP ratio: infant plasma/maternal plasma ratio, N = number, SD = standard deviation*

| Author | Study design | N participants | Maternal dose range (mg/day) | Maternal serum concentration (ng/mL) | Cord blood concentration (ng/mL) | Penetration ratio (cord blood/maternal plasma) |
|---|---|---|---|---|---|---|
| **Heinonen et al. 2021** | Randomized controlled trial | 9 women | 50-100 | 14.38 (3.64–24.17) | 4.28 (median) | CB/MP: 0.33 (median), range 0.14-1.17<br>IP/MP: 0.25 (median), range 0.23-0.26<br>*(no statistical significance)* |
| **Hendrick et al. 2003** | Prospective observational study | 11 women | 25-150 | 20.5 (3-50) | 14 (median), range <1-72 (7-35 hours after oral dose of 25-150 mg) | **0.23** (median), range 10-0.66<br>Maternal dose correlated significantly with cord serum concentration (r=0.71, df=9, p= 0.01) |
| **Hostetter et al. 2000** | Prospective observational study | 1 woman | 175 | 45 | 21 (21 hours after oral dose) | No penetration ratios given |

| | | | | | | |
|---|---|---|---|---|---|---|
| **Paulzen et al. 2017** | Prospective observational study | 6 women | 25-100 | 15.4 (9.7-30.1) | 5.85 (mean), range 3.1-9.3 (at steady-state concentrations) | 0.36 (median)<br>(Not statistically significant correlation with cord blood concentrations) |
| **Pogliani et al. 2016** | Prospective observational study | 10 women | 25-100 | 22.91 (7.4-**89.0)** | 7.39 (median), range <5-35.5 | 56.44% (mean), range 23.9-102.4%<br>*(stat. significance not specified)* |
| **Rampono et al. 2009** | Prospective observational study | 6 women | 50 | 15 (11-45) | 6 (median), range 4-11 | **0.33** (median), range 0.29-0.36<br>*(cord blood level increase not stat. significant)* |
| **Rampono et al. 2004** | Prospective observational study | 4 women | 50 | 23.5 (9-64) | 12.25 (mean), range 5-24 | **0.58** (mean), range 0.38-1.2<br>*(stat. significance not specified)* |
| **Sit et al. 2011** | Prospective observational study | 9 women | 50-200 | 17.2 (5.8-32.4) | 6 (mean), range 2.7-11.0 | **0.43** (mean), range 0.19-0.99<br>*(No significant association in SSRI cord-to-maternal blood ratios and perinatal events (95% CI, 0.2-32.8%)* |
| **Zheng et al. 2022** | Systematic review | 8 studies (41 women) | 25-200 | - | No concentrations given | **0.42** ± SD0.24<br>*(no linear correlation between the daily administered dose nor the time of blood sampling relative to last dose with the measured F:M concentration ratio (P = .482 and P = .955, respectively).* |

Data interpretation

Despite the large interindividual variation observed between and within studies (in part determined by the large interindividual variation in maternal sertraline plasma concentrations and otherwise derived from heterogeneity in study design), the placental transfer of sertraline, as measured at the end of pregnancy,[8] appears moderat**e.** A systematic review performed by Zheng et al. (2022) found a pooled penetration ratio[9] of 0.42 (standard deviation 0.24). This measure was based on a total of 8 studies including 41 women and their infants.

##### 3.3.2. What can be learnt from pharmacokinetic studies of the drug in pregnancy?

❖ *Instructions: describe data obtained from traditional as well as simulation-based (PBPK and pop-PK) studies.*

Ten in vivo pharmacokinetic studies in pregnancy were identified alongside two case reports and one meta-analysis that included three individual pharmacokinetic studies also featured below (Schoretsanitis et al., 2020).

[8] *It is generally assumed that the placental transfer of medications increases with placental blood flow throughout pregnancy and it can thus be posited that placental transfer at the end of pregnancy is maximal compared to earlier trimesters.*

[9] *Ratio of sertraline concentrations in umbilical cord blood and maternal plasma measured shortly after delivery.*

<u>Key information from in vivo pharmacokinetic studies of sertraline in human pregnancy</u>[10]

*Blue: decreasing mean/median maternal plasma concentration of sertraline during pregnancy; red: increasing mean/median maternal plasma concentration of sertraline during pregnancy; Abbreviations: AUC = Area Under the Curve, BMI: Body Mass Index, Cl = Clearance, C/D = Concentration-dose ratio, F = Fetal, M = Maternal, PP = Postpartum, T = Trimester*

| | **Study population** | **Measured concentrations (F,M)** | **Level of evidence** [11] | **Maternal dose range (mg/day)** | **Maternal & fetal concentration range (ng/mL)** | **AUC** | **Cl** | **Overall trend in maternal concentration during pregnancy** |
|---|---|---|---|---|---|---|---|---|
| **1 Bodnar et al. 2006** | * 1 woman<br>* 1 neonate | *Maternal Blood<br>***Cord blood**<br>***Neonatal blood 4 weeks PP** | C | 50 | *At birth: 11.2<br>*4 weeks PP: 26.9<br>*30 weeks PP: 26.3 | No | No | |
| **2 Colombo et al. 2021** | * 24 women<br>* 21 neonates | *Maternal blood<br>***Cord blood** | C | 25-150 | *T3: <5-42.7 (median 15.5)<br>*At birth: <5-89 (16.7) | No | No | |
| **3 Freeman et al. 2008** | * 8 women | *Maternal blood | B (pharmacokinetic curve) | 25-200 | Mean C/D<br>*T2**:** 0.37-1.33 (ng/mL/mg)<br>*T3: 0.37-1.25 *12-52 weeks PP: 0.37-1.25 | **Yes** | **Yes** | Not statistically significant **decrease in concentratio**n throughout pregnancy with lowest concentration in 3rd trimester and **large heterogeneity** across participants (changes in concentration may be clinically significant for some women). |

10 *The search and screening strategy for obtaining the latter pharmacokinetic studies is described in Supplement 1. The complete table of information obtained from the sampled pharmacokinetic studies can be requested to the MADAM team.*

11 *The level of evidence of pharmacokinetic studies was assessed based on a classification established by Gastine et al. 2019. Level B qualifies pharmacokinetic studies in which sufficient data was collected to construct a <u>pharmacokinetic curve</u> (continuous function of the plasma concentration of a medication over time). The availability of such a curve allows to derive pharmacokinetic parameters (e.g. the clearance of medication) with more accuracy than isolated plasma concentration measurements of a medication. Pharmacokinetic studies that used the latter sampling strategy are qualified as level C and are considered to be lower level evidence than B-level studies. Level B and C pharmacokinetic studies are both considered lower quality evidence than <u>population pharmacokinetic studies</u> in which data is obtained from larger groups of patients.*

| | | | | | | | | |
|---|---|---|---|---|---|---|---|---|
| **4 Heinonen et al. 2021** | * 9 women<br>* 7 infants | *Maternal blood<br>***Cord blood**<br>***Neonatal blood 48h PP** | C | 50-100 | Mean C/D<br>*T2: 0.15 (0.12-0.24 (ng/mL)(mg/day))<br>*T3: 0.19 (0.12-0.23)<br>*At birth: 0.19 (0.15-0.25)<br>*1 month PP: 0.25 (0.17-0.29) | No | No | Median **dose-adjusted concentrations** were **67%** lower in the 2nd trimester compared to postpartum, but the individual **variation was large** ( not statistically significant). |
| **5 Hendrick et al. 2003** | * 11 women<br>* 11 infants | *Maternal blood<br>***Cord blood** | C | 25-150 | *At birth: 3-50 | No | No | |
| **6 Hostetter et al. 2000** | * 1 woman<br>* 1 infant | *Maternal blood<br>***Cord blood**<br>***Amniotic fluid** | C | 150 | *At birth: 53 | No | No | |
| **7 Leutritz et al. 2023** | *11 women<br>*11 infants | *Maternal blood<br>***Cord blood**<br>***Neonatal blood >2 weeks pp** | C | Not specified | Mean C/D:<br>*T1: 0.56<br>*T2: 0.29<br>*T3: 0.39 | No | No | |
| **8 Paulzen et al. 2017** | * 6 women<br>* 6 infants | *Maternal blood<br>***Cord blood**<br>***Amniotic fluid** | C | 25-100 | *T3: 0.7-30.1 | No | no | |
| **9 Rampono et al. 2009** | *6 women<br>*6 infants | *Maternal blood<br>***Cord blood**<br>*** Neonatal blood 3 days PP** | C | 50 | *At birth: 15 (11-45) | No | No | |
| **10 Rampono et al. 2004** | *4 women<br>*4 infants | *Maternal blood<br>***Cord blood**<br>***Neonatal blood 5 days PP** | C | 50 | *At birth: 9-64 | No | No | |
| **11 Schoretsanitis et al. 2020 (meta-analysis) **** | **Freeman + Sit +Westin**<br>(n = 18) | *Maternal blood | **C** | 10-200 | | No | No | Mean alterations ratio (dose-adjusted concentration 3rd trim/baseline): 1.38 (range 0.77-1.53 per study; individual range 0.34-3.04) = **higher dose-adjusted concentration** |

| | | | | | | | | |
|---|---|---|---|---|---|---|---|---|
| **12 Sit et al. 2008** | *6 women | *Maternal blood | C | 50-200 | *20 weeks: 4-93<br>*30 weeks: 10-45<br>*36 weeks: 66-**147** (one woman 147, all other women < 75)<br>*At birth: 7-32<br>*2 weeks PP: 41-149<br>*4-6 weeks PP: 15-83<br>*12 weeks PP: 21-99 | No | No | **Decreasing dose-adjusted concentrations** were observed throughout pregnancy in five out of six women |
| **13 Stika et al. 2022** | *47 women | *Maternal blood<br><br>+ maternal genotype of CYP2C19 polymorphisms | B (pharmacokinetic curve) | 25-225 | C/D ratio mean (SD)<br>*4-8 weeks: 0.19 (0.05)<br>*8-12 weeks: 0.24 (0.11)<br>*12-16 weeks: 0.25 (0.16)<br>*16-20 weeks: 0.26 (0.15)<br>*20-24 weeks: 0.26 (0.17)<br>*24-28 weeks: 0.32 (0.2)<br>*28-32 weeks: 0.27 (0.17)<br>*32-36 weeks: 0.29 (0.19)<br>*36+ weeks: 0.24 (0.13)<br>*<8 weeks PP: 0.39 (0.23)<br>*>8 weeks PP: 0.32 (0.2) | No | Yes | * **Reduced mean C/D ratio** at GA **18 weeks** and relatively constant throughout pregnancy compared with postpartum<br>* 3rd trimester mean C/D ratio **22% less than after delivery** (not statistically significant)<br>* **Marked interindividual variability** in both absolute C/D ratios & intra-individual trajectories<br>* **BMI** not significantly associated with C/D ratios<br>* No meaningful differences among the individual trajectories of the **CYP2C9** and **CYP2D6** phenotypes<br>* Mean C/D ratios in participants with **functional CYP2C19** activity did not change significantly during pregnancy, whereas ratios in participants with **poor or intermediate CYP2C19 activity decreased by 51%** (no information on statistical significance)<br>* **In CYP2C19 PM/IMs,** the decrease in plasma sertraline concentrations during pregnancy may reflect increased **clearance by other CYP enzymes, most likely CYP3A4 and/or CYP2B6** |
| **14 Westin et al. 2017** | *34 women | *Maternal blood | C | 10-100 | *T1: 9.8<br>*T2: 12.2<br>*T3: 15.1 | No | No | **Mean increased serum concentration (+68%; P < 0.001)** during pregnancy (attributed to CYP2C19 inhibition) |

<u>Data interpretation</u>

*Study quality*

The sampled studies were characterized by a heterogeneous design and a relatively low level of evidence. Sample size was small (ranging from one mother-infant pair to 47 pregnant women; two studies of over 20 participants). Pregnant participants received various doses of sertraline within the standard range (25-200 mg) and dose-adjusted concentrations were not always calculated. While all studies described maternal measurements, only five studies performed measurements in the second trimester and one in the first trimester (Leutritz et al., 2023). Nine studies reported fetal and/or neonatal measurements. The latter measurements were performed in different mediums (cord blood, neonatal blood, amniotic fluid), rendering comparison more difficult. No study reported maternal or fetal plasma levels of unbound sertraline.[12] The timing of concentration measurement after ingestion and/or birth was almost never indicated. Only two studies reported a full pharmacokinetic curve (Freeman et al. 2008 and Stika et al. 2022) with one study describing an area under the curve and a sertraline clearance (Freeman et al., 2008).

*Key findings*

- All but one pregnant participant (in Sit et al., 2008) had sertraline plasma concentrations within the reference concentration range of 10-75 ng/mL proposed by Braten et al. throughout pregnancy. The latter pregnant woman had concentrations of 141 ng/ml at 30 weeks and of 147 ng/ml at 36 weeks of pregnancy. Both concentrations, it should be noted, are within the reference range (10-150 ng/mL) proposed by the AGNP. The dose of sertraline that she received was not reported.
- Five pharmacokinetic studies (n = 81 pregnant women) reported decreases in the mean or median maternal plasma concentrations of sertraline during pregnancy (Freeman et al., 2008; Heinonen et al., 2011; Leutritz et al., 2023; Sit et al., 2008 and Stika et al., 2022). The measured decreases were especially pronounced in the second and third trimesters . Alongside significant heterogeneity in study design (e.g. different baseline concentrations used for comparison), a large intra-individual variability was reported and the measured decreases in plasma concentrations were never statistically significant. One study by Westin et al. (2017) (n = 34 women) reported an increase in sertraline plasma concentrations (+68%; $P < 0.001$) in the third trimester compared to baseline.
- One study by Stika et al. (n = 47 pregnant women) examined associations between maternal sertraline plasma concentrations in pregnancy and various maternal CYP enzyme phenotypes and found that while there were no differences in the trajectories of sertraline plasma concentrations of pregnant individuals with differing CYP2C9 and CYP2D6 phenotypes, different CYP2C19 phenotypes were associated with distinct trends in sertraline plasma concentrations during pregnancy. Whereas the mean concentration dose ratios (concentrations adjusted to the ingested dose) in participants with functional CYP2C19 activity (EMs and UMs; n = 39) did not change significantly during pregnancy, they were 51% lower in participants with poor or intermediate CYP2C19 activity (PMs and IMs; n = 7) at 36 weeks compared to 8 weeks postpartum. According to the authors, the observed decrease in plasma sertraline concentrations in CYP2C19 PM and IMs during pregnancy may reflect increased clearance by other CYP enzymes, most likely CYP3A4 and/or CYP2B6 as a result of induction in pregnancy.

In addition, two pregnancy PBPK models were identified for sertraline, both of which simulated the maternal exposure to sertraline during pregnancy.

[12] *The unbound fraction of a medication is generally the therapeutically active portion of this medication.*

Key characteristics and findings from pregnancy PBPK models on sertraline

*EM = extensive metabolizer[13], F = fetal, M = maternal, NM = normal metabolizer PM = poor metabolizer, UM = ultrarapid metabolizer*

| | Study objective | Full/ partial model | Elimi- nation pathways | Predicted exposure (F, M) | Reference therapeuti c range | Verification | Pharmacokinetic observation | Dose recommendation |
|---|---|---|---|---|---|---|---|---|
| **George et al. 2020 (FDA)** | To develop a PBPK model that allows gestational-age dependent prediction of sertraline dosing in pregnancy. | Partial | Hepatic clearance (CYP3A4, CYP2B6, CYP2C9, CYP2C19, CYP2D6)<br><br>Contributio n based on **Obach et al.** | Maternal exposure during pregnancy | None | Nonpregnant validation Ronfeld et al.<br><br>Pregnancy validation Freeman et al. | **Decreased concentration** (not further specified) | Gestational-dependent changes in physiology and metabolism account for **increased clearance** of sertraline, potentially leading to **underdosing** of pregnant women when nonpregnancy doses are used. |

[13] The Dutch Pharmacogenetics Working Group now refers to extensive metabolizers as 'normal metabolizers'. We thus use the terminology EM/NMs in the tables.

| | | | | | | | | |
|---|---|---|---|---|---|---|---|---|
| **Almurjan et al. 2021** | To assess gestational changes in sertraline trough plasma concentrations for **CYP2C19 phenotypes, a**nd identify appropriate dose titration strategies to stabilize sertraline levels within a defined therapeutic range throughout gestation | Full | Hepatic clearance (CYP3A4, CYP2B6, CYP2C9, CYP2C19, CYP2D6)*<br><br>** Respective contribution of CYP enzymes based on Obach et al.* | Maternal exposure during pregnancy | 10–75 ng/<br><br>ml (Bråten et al., 2020) | Nonpregnant validation<br><br>Niyomnaitham et al., Chen et al, Kim et al., Saletu et al. and Ronfeld et al.<br><br>Pregnancy validation<br><br>Westin et al. | **In all tested phenotypes, intrinsic clearance decreased** throughout pregnancy, mirroring **decreases in CYP2C19 activity,** the largest significant difference in clearance being noticed in trimester 3. The decrease in clearance was expected to increase sertraline trough plasma concentrations, on the contrary, **trough plasma concentrations for EMs and UMs decreased during gestation,** the greatest significant decrease occurring in trimester 3. | * **Regardless of the phenotype,** the daily sertraline dose required to maintain trough concentrations within the reference range was **above the usual 50 mg/day** throughout pregnancy<br><br>* **All CYP2C19 phenotypes required a dose increase throughout gestation**. For EMs/NMs and UMs, doses of 100–150 mg daily are required throughout gestation. For PMs, 50 mg daily during trimester 1 followed by a dose of 100 mg daily in trimesters 2 and 3 are required. |

Summary and interpretation of data

**George et al. (2020)** developed a partial PBPK model[14] and simulated decreasing maternal concentrations of sertraline during pregnancy for daily doses of 200 mg. No additional dosing scenarios were explored.

**Almurjan et al. (2021)** developed a full PBPK model to assess gestational changes in the maternal plasma concentration of sertraline and to explore dosing scenarios for different maternal CYP2C19 phenotypes. Braten et al. (2020)'s proposed concentration range for non-pregnant adults (10-75 ng/ml) (study described in 3.2.2.) was used as a reference for adequate maternal exposure during pregnancy as part of dose finding. The assessment of the credibility and applicability of Almurjan et al.'s model for dose finding in pregnancy is described in **Supplement 2.**[15] In short,

- A key limitation of the model was that it only predicted the maternal and not the fetal exposure to sertraline during pregnancy

[14] *As opposed to 'full' PBPK models, partial models only offer a coarse representation of the anatomy and physiology of organs in pregnant women's bodies (limited numbers of few organs considered). Because they incorporate less detailed data, the predictions of partial PBPK models are considered less accurate. While partial models can be used to evaluate the safety of a medication in a given population, full models tend to be preferred to explore different dosing scenarios and how they affect the plasma exposure to the medication.*

[15] *George et al.'s model was not evaluated using the Decision Tree for Model-Informed Dosing because it did not explore dosing scenarios.*

- The credibility of the model was deemed satisfactory overall: while only one observational study in pregnant women (Westin et al., 2017, described in 3.3.2) was used for validation even though other pharmacokinetic studies were available, the simulated maternal concentrations were within the range of Westin et al.'s observed concentrations. Importantly Westin's study did not provide differentiated data for various CYP2C19 polymorphic variants, meaning that this aspect of Almurjan et al.'s model was not compared with observational data. Of note, Almurjan et al. predicted a decrease in maternal plasma concentrations of sertraline during pregnancy as opposed to the increase in observed sertraline plasma concentrations reported by Westin et al. The predicted decrease in maternal plasma concentrations of sertraline, however, aligned with trends in the observed maternal concentrations of sertraline described by the majority of available pharmacokinetic studies (Freeman et al., 2008; Sit et al., 2008; Stika et al. 2022 - all described in 3.3.2). Although the predicted increase in maternal sertraline concentrations during pregnancy appeared to contradict the inhibition of CYP2C19 and decreased hepatic clearance in pregnancy that were also simulated by the model, the physiological explanations put forward by Almurjan et al. to justify this discrepancy were credible (see below). Lastly, Almurjan et al. only reported simulations of trough maternal concentrations and no maximum concentrations of sertraline. However, given the use of Braten et al.'s conservative reference concentration range to guide dose finding and the cautious approach that was adopted with regards to exposure matching - more tolerance for underdosing than overdosing in the virtual pregnant population- the risk of maternal overdosing for Almurjan et al.'s proposed dose recommendations is considered low.

Almurjan et al.'s findings were as follows:

- In all tested CYP2C19 phenotypes, hepatic clearance decreased throughout pregnancy, mirroring decreases in CYP2C19 activity, the largest significant difference in clearance occurring in the third trimester.
- The predicted decrease in hepatic clearance was expected to increase the predicted maternal plasma concentrations of sertraline during pregnancy. On the contrary, sertraline plasma concentrations for extensive and ultrarapid CYP2C19 metabolizers (and to a lesser extent for CYP2C19 poor metabolizers) decreased during gestation, as illustrated in Figure 1. The most significant decrease occurred in the third trimester. To explain these seemingly contradicting findings Almurjan et al. referred to a) the induction of the four other CYP enzymes involved in sertraline metabolism as a result of hormonal changes in pregnancy that would likely counterbalance CYP2C19 inhibition (but did not prevent an overall decrease in hepatic clearance) and to b) **other**

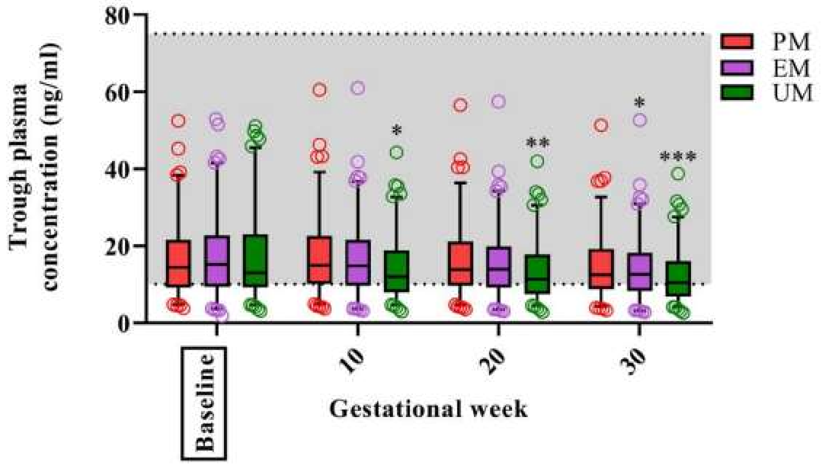

*Figure 1 – Simulated sertraline trough plasma concentrations for CYP2C19 polymorphs from Almurjan et al. 2021*

**pregnancy-induced physiological changes of potentially greater magnitude including a decrease in albumin concentration** leading to a higher volume of distribution and hence to lower plasma concentrations during pregnancy. The latter variable was demonstrated to be a covariate for predicted sertraline concentrations through a global sensitivity analysis conducted by Almurjan et al.

- Regardless of the CYP2C19 phenotype, the daily sertraline dose required to maintain sertraline trough concentrations within the reference range was above 50 mg per day throughout pregnancy.
- All CYP2C19 phenotypes required a dose increase throughout gestation. For normal metabolizers and ultrarapid metabolizers, doses of 100–150 mg daily are required throughout gestation. For poor metabolizers, 50 mg daily during the first trimester followed by 100 mg daily in the second and trimesters are required.

#### Pharmacodynamics

##### 3.3.3. Is disease progression similar in pregnancy than in nonpregnant adults?

❖ <u>Context</u>: *The following paragraph describes whether disease progression in pregnancy differs from disease progression in nonpregnant adults looking at the pathophysiology, natural history etc. We discuss how this may affect dosing needs.*

As previously outlined, epidemiological data show a higher incidence of depressive symptoms in pregnant women compared to nonpregnant adults. Furthermore, the incidence of depressive symptoms increases as pregnancy progresses, with the largest incidence of symptoms observed in the third trimester (Van de Loo et al. 2018).[16] The pathophysiology of antenatal depression remains poorly understood however. In addition to the likely influence of psychosocial factors on the occurrence of depressive symptoms during pregnancy (Oh et al. 2022), it is probable that physiological and especially hormonal changes during pregnancy contribute to a higher risk of depression during that period (Brummelte et al., 2016). The latter hormonal changes may also play a role in the observed decrease in anxiety symptoms during pregnancy. In view of our limited understanding of these mechanisms however, it remains hard to take them into account for dosing sertraline in pregnancy.

##### 3.3.4. How does pregnancy influence the pharmacodynamics of the drug?

❖ <u>Context</u>: *The following paragraph describes whether physiological changes in pregnancy are likely to affect the pharmacodynamics (i.e. drug action) of the medication during pregnancy and the extent to which this may affect dosing needs*

Apart from its likely effect on the plasma concentration of sertraline, the previously described decrease in albumin concentration during pregnancy may also affect the unbound fraction of sertraline. This may potentially lead to changes in the effectiveness of the medication at a given plasma concentration. The latter effect has not been investigated however. In addition, it is likely that hormonal changes as part of pregnancy affect the pharmacodynamics of sertraline by influencing the availability of and response to serotonin. In particular, increased estrogen in pregnancy is thought

[16] *And especially in the post-partum period.*

to affect serotonin levels and response through different pathways, for example by influencing the sensitivity of serotonin receptors and the breakdown of serotonin (Lokuge et al. 2011). Again, our understanding of these pregnancy-induced changes in the pharmacodynamics of sertraline appears insufficient to adjust dosing needs accordingly in pregnancy.

**3.3.5. Summarize the available evidence on dose-related efficacy of the drug in pregnancy.**

No literature was found on dose-related efficacy of sertraline in pregnancy.

**3.3.6. Summarize the available evidence on dose-related toxicity of the drug in pregnancy.**

A combination of literature searches (see **supplement 1**) were conducted to investigate a potential dose relationship between maternal use of sertraline during pregnancy and the various perinatal and fetal outcomes listed in the safety assessment made by Lareb and the NVOG (paragraph 3.1.3). Searches were conducted for the following outcomes: dysmaturity, postpartum bleedings, poor neonatal adaptation syndrome (PNAS), persistent pulmonary hypertension of the neonate (PPHN) and fetal cardiac anomalies.

- No studies were found that explored a dose relationship between sertraline use and dysmaturity, postpartum bleedings, PPHN or fetal cardiac anomalies.
- One study was found that identified a dose relationship for maternal sertraline use in the third trimester of pregnancy and the incidence of PNAS. Brumbaugh et al. 2023 (retrospective cohort study, n= 471 mother-infant pairs with sertraline use as monotherapy in pregnancy, 15 (3.2%) infants that developed PNAS) described a relative risk of PNAS of 3.64 (95% CI 1.27-10.47, $p = 0.07$) for high daily doses of ($\geq$ 100 mg) sertraline in the third trimester compared to standard daily doses (50 mg).

**3.3.7. Are there adverse events associated with use of the drug that may influence the recommended dose in pregnancy?**

❖ *Context: The following paragraph describes adverse events associated with use of the medication that may be of importance for dosing in pregnancy.*

A risk of spontaneous and/or prolonged bleeding has been signaled for sertraline (FTK). While very rarely observed in nonpregnant adults (clinical expert), caution may be advised in pregnant women with a history of spontaneous bleedings concomitant with sertraline use. This is also true in regard to a possible association between sertraline use during gestation and postpartum bleedings (see 3.1.3.). Similar caution should be used in the event of concomitant use of anticoagulant medications in pregnancy.

**3.3.8. Are there drug interactions associated with use of the drug that may influence the recommended dose in pregnancy?**

❖ *Context: The following paragraph describes drug interactions undergone by the medication that may be of importance for dosing in pregnancy.*

See paragraph 3.1.8 on anticoagulants. Caution should be used when using sertraline in combination with other medications that are metabolized by or that may impact the activity of CYP enzymes involved in the metabolism of sertraline, especially CYP2C19 (major pathway, inhibited activity during pregnancy - see more information in paragraph 3.2.2.).

##### 3.3.9. Are there (patho)physiological covariates of influence for the pharmacokinetics of the drug?

❖ *<u>Context</u>: The following paragraph discusses the influence of covariates including but not limited to obesity, renal impairment and liver insufficiency.*

Body Mass Index was not significantly associated with dose-adjusted concentrations in pregnant women. This finding aligned with a previous pharmacokinetic study of sertraline in nonpregnant adults (Reis et al., 2004).

##### 3.3.10. To what extent can postpartum changes in pharmacokinetics and pharmacodynamics inform the postpartum dose?

❖ *<u>Context</u>: The following paragraph discusses the influence of postpartum changes on the pharmacokinetics and pharmacodynamics of the medication for the chosen indication(s) and potential dosing requirements in that period of time.*

As is the case for most medications, little information can be found on changes in the pharmacokinetics of sertraline shortly after birth. While the maternal physiology is assumed to largely return to a pre-pregnancy state within 4-6 weeks postpartum, it is also likely that gestation induces long-term changes in maternal physiology (Quinney et al., 2023). Postpartum changes that may be of importance for sertraline are listed below:

<u>Postpartum changes in maternal physiology that may be of importance for sertraline pharmacokinetics and dosing</u>

*GFR: glomerular filtration rate*

| Physiological parameter | Factor of influence during pregnancy | Trends in physiological parameter in the immediate postpartum period |
|---|---|---|
| CYP metabolism | Hormonal changes | - Inhibition of CYP enzymes, especially when partial, generally wanes off immediately after pregnancy due to an abrupt decline maternal hormones postpartum (Brummelte et al. 2016)) (pharmacological expert)<br>- Induction of CYP enzymes takes 2 to 3 weeks postpartum to recede (pharmacological expert)* |
| Albumin concentration | Increase in total body water (haemodilution) and GFR, hormonal changes, comorbidity (e.g. gestational diabetes, preeclampsia) | Albumin concentration rapidly increases in the first 2 to 3 weeks postpartum until stabilizing 15 weeks after delivery (Zhao et al. 2021). |

All in all, it is likely that changes in maternal physiology that affect the pharmacokinetics of sertraline during pregnancy largely normalize by 2 or 3 weeks postpartum. From a pharmacokinetic perspective, this means that in case of a pregnancy-adjusted dose of sertraline, the dose prescribed in the third trimester could be re-adjusted to pre-pregnancy levels around that time. Considering the impact of postpartum hormonal changes on the occurrence postpartum depression however (Brummelte et al., 2016), it is common practice to maintain (i.e. not decrease) the dose of sertraline that was used by pregnant women at the end of their pregnancy for the next six months to one year in line with clinical guidance on SSRI dose titration for nonpregnant adults (NVvP, 2010).

#### 3.4. Proposed dose

**3.4.1. If a therapeutic range has been defined for non-pregnant patients (3.2.6.), can this range be used in pregnancy or do pregnancy-related changes in disease progression, drug efficacy and safety (as outlined in 3.3.3-3.3.6) warrant a pregnancy-specific therapeutic range? In both cases, discuss whether a dosing strategy can be defined leading to appropriate maternal and fetal exposures.**

Given the lack of information on the dose-related efficacy and dose-related toxicity of sertraline in pregnancy, a pragmatic as well as cautious approach consists in following the most conservative reference range that is available to guide sertraline exposure in nonpregnant adults. We thus propose to use Braten et al.'s range of 10-75 ng/ml as a reference for dosing in pregnancy. Please refer to question 1.2 for the dose recommendation that will be published on the TIS website.

**3.4.2. If no therapeutic range has been defined outside of pregnancy (3.2.6), can a dosing strategy be identified that matches observed exposures in non-pregnant patients and that appears warranted based on available information outlined in 3.3.3-3.3.**6? See 3.4.1.

##### Conclusion of part 3:

- Non-pregnant population: given the large interindividual variation in the pharmacokinetics of sertraline (most likely related to genetic variation in CYP2C19) and the absence of a well-defined therapeutic range, sertraline dosing needs to be individually titrated based on a careful follow-up of side effects and clinical response.
- It is likely that pregnancy-induced changes in maternal physiology result in an overall decrease in the maternal plasma concentration of sertraline, especially in the second and third trimesters. The latter decrease is likely to be for a large part caused by the combined effect of CYP enzyme induction (apart from CYP2C19) and a decrease in albumin concentration during pregnancy.
- Maternal exposure to sertraline during pregnancy is marked by the presence of large interindividual variability which can most likely be attributed to genetic variation in CYP2C19.
- All in all, these pharmacokinetic findings suggest that sertraline doses should not be reduced antenatally and may even have to be increased for certain pregnant women in the second or third trimesters depending on the observed clinical response to treatment.
- Our knowledge of the effect of pregnancy on the pharmacodynamics of sertraline is insufficient to inform a pregnancy-adjusted dose.
- While the data on dose-related fetal toxicity of sertraline appears very scarce (none available for dysmaturity, PPHN or cardiac anomalies), evidence of a dose-relationship between the occurrence of poor neonatal adaptation syndrome and the use of doses of sertraline > 100 mg in the third trimester of pregnancy was found. The latter dose-effect relationship may also exist for other fetal or neonatal outcomes and may constitute a trade-off for increasing the dose in the third trimester of pregnancy.

#### PART 4 – DOSE IMPLEMENTATION

- *Context: the following section summarizes the information previously collected as part of the dose rationale document and outlines other relevant clinical and practical considerations to inform a balanced assessment of the maternal and fetal risks and benefits and remaining uncertainties associated with various possible dosing strategies for the medication at hand. The aim is to issue a dose recommendation in pregnancy that is acceptable, clinically feasible and evidence-based. Input from the clinical expert(s) consulted as part of the dose rationale document writing process should be obtained on the following questions. Feedback from the MADAM Working Committee on this section is also critical.*

##### 4.1. Feasibility

###### 4.1.1. How feasible is the proposed dose in the relevant setting(s) of care?

- *Context: assess the feasibility of the proposed dose recommendation considering aspects such as available drug formulations, medication preparation and costs in the chosen setting(s) of care (consider the various relevant levels of healthcare delivery and the targeted country(ies)). If the proposed dose recommendation does not appear feasible, could it be altered in such a way that it is attainable while still being evidence-based? The acceptability of the proposed dose and the availability of monitoring strategies are addressed in later questions.*

Because it does not fundamentally deviate from the dose recommendation for nonpregnant adults, no practical hurdles are expected in the implementation of the proposed dose in primary and secondary healthcare in the Netherlands.

###### 4.1.2. How is the proposed dose recommendation expected to affect current healthcare resources and processes?

*Context: consider any implication for healthcare demand, work routines, medication costs and procurement (e.g. if a different formulation is recommended).*

The proposed dose may indirectly increase the prevalence of antenatal sertraline use  by offering new evidence underlying the benefit-risk ratio of sertraline use in pregnancy and thereby potentially limiting the practice of ad hoc sertraline discontinuation by HCPs or pregnant women. While an adequate follow-up of the clinical response to sertraline is recommended during pregnancy, the latter follow-up is already common practice, at least in secondary healthcare (clinical expert). Implementation of the proposed dosing strategy however increase the burden on Psychiatry, Obstetrics and Pediatrics outpatient clinics (POP poli's), assuming that more pregnant women using sertraline will be referred to the latter clinics by primary care HCPs that feel less confident about making decisions on antenatal sertraline dosing. Lastly, the proposed dose recommendation may lead to an increased demand for pharmacogenetic testing, and thus additional costs.

#### 4.2. Desirability of the proposed dose

##### 4.2.1. What is the level of confidence in the proposed dose?

❖ *<u>Context</u>: assess the overall level of confidence in the proposed dose recommendation looking at the availability and quality of the evidence summarized in part 3. Consider both the fetal and maternal adequacy of the recommended dose(s) and the level of certainty of the dose-exposure-response relationship and any other remaining areas of uncertainty for dosing. If relevant for the indication(s), take into consideration all trimesters of pregnancy.*

The level of confidence in the proposed dose is sufficient. While no therapeutic range can be defined for sertraline due to a high interindividual variability in the dose-exposure-response, Braten et al. 's reference range of 10-75 ng/ml can be used as a conservative standard for adequate maternal plasma exposure. Pharmacokinetic data from ten studies (> 180 pregnant women) show that for daily dose ranges of 50-200 mg used in pregnancy, observed maternal plasma concentrations almost never exceed the proposed reference concentration range. Furthermore, most available studies suggest that on average, without dose adjustments, the maternal plasma concentrations of sertraline tend to decrease in pregnancy, especially in the second and third trimesters. Additional data obtained from pharmacokinetic modelling show a similar trend across pregnant individuals with various CYP2C19 phenotypes, with a greater decrease in the predicted plasma concentration of sertraline of CYP2C19 EMs and UMs than PMs (of note: this finding does not align with the pharmacokinetic data collected by Stika et al. who report a non-significant decrease in the plasma concentration of sertraline in CYP2C19 intermediate metabolizers and PMs as opposed to participants with functional CYP2C19 activity in pregnancy). Physiological explanations put forward to support the observed and predicted decrease in plasma concentration of sertraline during pregnancy - i.e. the induction of four out of five hepatic enzymes involved in the metabolism of sertraline, and a decrease in the concentration of albumin leading to a higher distribution volume, and in turn, lower sertraline plasma concentrations during pregnancy- appear sound. A remaining area of uncertainty concerns the fetal safety of comparatively higher doses of sertraline as information on a potential dose relationship between fetal and neonatal outcomes and sertraline exposure is mostly lacking apart from PNAS. Existing teratologic data on sertraline use in the standard dose range (25-200 mg) however supports the current benefit-risk ratio for the proposed antenatal dosing strategy for sertraline in clinically relevant cases of depression and/or anxiety disorders (see **Figure** 1 p. 38).

##### 4.2.2. What are the expected benefits of (not) implementing the dose?

❖ *<u>Context</u>: consider the maternal and fetal benefits associated with the proposed dose recommendation. If the latter recommendation diverges from the dose recommendation currently applied in pregnancy, compare the benefits of using the adjusted dose with the benefits of using the standard dose.*

The primary benefit of the recommended sertraline dosing strategy consists in an enhanced probability of achieving an effective treatment for maternal depression and/or anxiety symptoms during pregnancy. This outcome has proven to be beneficial for both mothers and their fetuses in cases where the indication for pharmacological treatment has been established (Sit et al. 2008).

##### 4.2.3. What are the expected risks associated with (not) implementing the dose?

❖ *<u>Context</u>: consider the likelihood of underdosing and overdosing considering the width and certainty of the proposed therapeutic or reference concentration range. Review the acceptability of underdosing and overdosing in light of the severity of the investigated indication(s) of the medication and the efficacy of the medication for treating this indication.*

<u>Likelihood of underdosing and overdosing</u>

Given the width of the available reference concentration ranges of sertraline in nonpregnant adults and the use of the most conservative range as a reference for establishing the recommended dosing strategy in pregnancy, the probability of maternal underdosing or overdosing occurring for the proposed doses appears limited despite the presence of high interindividual variability in sertraline exposure and response. Given the cautious approach adopted by Almurjan et al. for dose finding across CYP2C19 phenotypes - use of a conservative reference concentration range, low tolerance for overdosing as opposed to underdosing in the virtual pregnant population and recommended doses at the lower end of the standard dose range for nonpregnant adults- maternal underdosing may occur in some women at the lower end of the proposed dosing range. Maternal overdosing, on the other hand, is unlikely to occur even at the higher end of the range. As discussed in paragraph 3.2.5, maternal toxicity appears highly unlikely for these exposure levels. Despite the scarcity of fetal pharmacokinetic and safety data, the moderate placental transfer of sertraline and the availability of teratogenicity data for habitual dose ranges (50-200 mg) suggest that risks of fetal toxicity are also limited.

<u>Consequences and acceptability of underdosing and overdosing</u>

Both underdosing and overdosing have poor acceptability given the potentially wide-ranging consequences in the context of pregnancy:

- **Underdosing** (= suboptimal maternal efficacy of treatment for depression and/or anxiety symptoms) may have adverse consequences for a pregnant woman and her fetus. E.g. suboptimal treatment of antenatal depression is associated with multiple risk factors for complications during pregnancy including alcohol consumption, smoking and poor antenatal care (level of evidence 3[17]) (Fumeaux et al. 2019). Furthermore, mood disorders have been associated with several obstetric complications including preterm delivery and low birth weight (Wu et al. 2003). While underdosing may not be the (only) cause for suboptimal treatment given the fact that sertraline is a second-line treatment for depression or anxiety disorders in pregnancy and given the lack of clear dose-exposure-response relationship, adequate (pharmacological) treatment appears critical especially in cases of severe depression or anxiety disorders.
- **Overdosing**, while seldom associated with toxicity in nonpregnant adults, remains highly undesirable in pregnancy, both for the mother and fetus. Even in the absence of data on a dose threshold for **fetal or neonatal toxicity** for outcomes other than PNAS, a relationship between the maternal dose of sertraline and fetal complications is plausible for other fetal or neonatal outcomes. Of note, PNAS, despite a high incidence in

---

[17] *Studies were classified in 5 levels according to their level of evidence: Level 1 = Systematic review of randomized controlled trials (RCT), Level 2 = RCT or observational study of high methodological quality with dramatic effect, Level 3 = Non-randomized controlled trial or prospective cohort study, Level 4 = Case-series, case-control study or retrospective cohort study, Level 5 = Mechanism-based reasoning.*

sertraline exposed neonates, is a transient and self-limiting outcome of limited severity, making it more acceptable than severe neonatal outcomes such as PPHN. While a dose-response is also likely for PPHN, the increased likelihood of PPHN for higher sertraline doses (that are still within the normal range for non-pregnant adults) is deemed acceptable given the low absolute incidence of PPHN, even among SSRI-exposed neonates. Cardiac defects, which result from impaired organ genesis in the first trimester of pregnancy, are unlikely to be associated with dose adjustments in the second and third trimesters. This outcome, therefore, is not taken into consideration in the risk-benefit analysis for adjusted sertraline dosing in pregnancy.

- Efforts should be made to minimize the risk of **maternal adverse reactions** in regard to maternal well-being and therapy adherence.

**4.2.4. Are risk mitigation strategies available, feasible and warranted within the relevant setting(s) of implementation?**

❖ *Context: assess the applicability of the previously described monitoring strategies and associated interventions (paragraphs 3.2.6. to 3.2.8) to increase the level of confidence in the maternal and fetal efficacy and safety of the recommended dose during pregnancy. Consider the feasibility and acceptability of such strategies in the various relevant settings of implementation.*

Monitoring of clinical response and dose adjustments

Monitoring the clinical response to sertraline and the occurrence of side effects appears key and the primary measure to reduce the risks of **maternal underdosing or overdosing**. Regular follow-up should therefore be organized for both pre-existing users of sertraline (especially in the second and the third trimester of pregnancy) as well as new users (throughout pregnancy, in line with standard follow-up at SSRI treatment initiation). The follow-up should focus on identifying potential signs of suboptimal clinical response and/or unacceptable side effects. If a suboptimal response or side effects are detected, it is then possible to increase or decrease the dose of sertraline on a weekly basis in line with clinical guidance for nonpregnant adults (NVK, 2022; NVvP 2010). Depending on the severity of symptoms and patient preference, follow-up can be performed in a primary care or secondary care setting.

With regards to the risk of **fetal toxicity:** apart from cardiac anomalies and dysmaturity for which associations with sertraline use in pregnancy have been shown, no signs of fetal toxicity can be detected antenatally. Detection of the latter signs as part of regular obstetric controls should however lead to an assessment of a patient's sertraline dosing, among other potential causal factors. Given the demonstrated association between moderately high doses (≥100 mg) of sertraline in the third trimester and PNAS, a precautionary approach might then be to not increase the dose in the third trimester. Important considerations in this view however, are the risk of postpartum depression, which may increase with suboptimal treatment at the end of pregnancy, and the fact that PNAS is transient in nature and moderate in severity for sertraline (NVOG, 2012).

Genetic testing for CYP2C19 polymorphisms

Genetic testing should be considered if unexplained side effects or an insufficient response to sertraline treatment are reported during pregnancy, especially for women with a history of side effects or poor response to CYP2C19 metabolized drugs (NVvP, 2020). A dose reduction may be advised for CYP2C19 PMs in particular but should be considered for all pregnant women reporting unacceptable side effects, independently of CYP2C19 phenotype. Genetic testing can be ordered by HCPs at all levels of care in the Netherlands. One should take into account the time needed to obtain the test results (4 to 6 weeks) which should not delay potential dose adjustments. CYP2C19 testing costs 120-180 EUR and is reimbursed if medically indicated (Erasmus MC, Pharmacogenetics).

TDM

The discrepancies in clinical guidance on sertraline TDM in nonpregnant adults have been discussed in paragraph 3.2.6. Looking at the relevance of TDM as a risk mitigation strategy in pregnancy, it is worth noting that TDM-monografie lists pregnancy as one of the indications for TDM of sertraline. The FTK furthermore advises that HCPs consider regularly determining pregnant women's plasma concentration of sertraline in view of pregnancy-induced changes in the pharmacokinetics of sertraline. Given the lack of clear therapeutic range for sertraline, we advise to consider TDM only if (unexplained) side effects or a diminished response to sertraline with no other plausible explanation are observed during pregnancy. TDM can then be used to identify potential outliers in plasma concentrations, to monitor clinically relevant changes in concentration over the course of pregnancy and/or as a precautionary step prior to dose increase especially in the third trimester. Given the lack of a well-defined dose-exposure-response relationship for sertraline, one may choose to follow the most conservative available reference range, namely 10-75 ug/l (Braten et al., 2020). TDM can be ordered at all levels of care in the Netherlands.

**4.2.5. What are the residual risks of implementing the dose and how do they compare to the residual risks of the current dose?**

Compared to standard dosing practice in pregnancy (which as discussed often involves lowering the dose or interrupting sertraline use preemptively out of concern for fetal safety), the proposed dose recommendation can help reduce the risk of maternal underdosing and the associated maternal and fetal complications. Regular clinical follow-up and when indicated, genetic testing can be used to inform potential ad hoc dose adjustments in pregnancy, thereby lowering the risk of maternal underdosing and overdosing. As previously outlined, the risk of fetal toxicity, while it may exist for certain neonatal outcomes (especially PNAS for which a dose-relationship was shown for daily doses of sertraline $\geq$ 100 mg in the third trimester of pregnancy), appears moderate overall given the conservative nature of the proposed dosing strategy and its alignment with standard dosing ranges of sertraline (25-200 mg) for which observational data in pregnancy are available and reassuring.

**4.2.6. Are there remaining knowledge gaps regarding the adequacy of the proposed dose?**

Some remaining knowledge gaps include:

- The pathophysiological mechanisms through which pregnancy influences depression and anxiety symptoms
- The influence of pregnancy on the pharmacodynamics of sertraline (dose-response)
- The factors that determine the contributions of different CYP enzymes to the metabolism of sertraline during and outside of pregnancy
- The influence of pregnancy on CYP phenotypes (phenoconversion?)
- The nature and speed of postpartum changes in the pharmacokinetics of sertraline, as well as the influence of lactation on these processes
- Potential associations between fetal and neonatal toxicity (e.g. congenital heart defects, PPHN) and sertraline dose or exposure

**4.2.7. Do the expected benefits, residual risks and the level of confidence in the proposed dose warrant its clinical use?**

❖ <u>Context</u>: *complete the following table and outline the conclusions drawn by the Working Committee after evaluating the risks and benefits presented in this table. Discuss the risks and benefits associated with the proposed dose in comparison to the benefits and risks associated with alternative interventions. NB: if a dose recommendation in pregnancy cannot be issued due to excessive uncertainty, outline any remaining knowledge gaps that should first be addressed. Determine which of these knowledge gaps should be included in the research agenda of project MADAM.*

Based on answers to 4.2.2 to 4.2.5 and weights attributed to the maternal risks and benefits below (see **supplement 3** on the principles used for weight attribution), the following risk-benefit balance can be established:

**Maternal & fetal risks and benefits associated with proposed antenatal sertraline dosing strategy in pregnancy**

| **Maternal risks** | **Maternal benefits** |
|---|---|
| - Limited: no toxicity observed for standard dose range in nonpregnant adults ⇔ | **- Better controlled antenatal depression** ⥣<br>*(Caveat: dose-effect relationship for sertraline is unclear at a population level, but likely exists at the individual level)*<br>**- Reduced likelihood of postpartum depression** ⥥<br>*(further decreased)* |
| **Fetal risks** | **Fetal benefits** |
| - Possible risk of dysmaturity ⥣<br>- Low risk of persistent pulmonary hypertension of the newborn (PPHN) if sertraline is used in third trimester ⥣<br>**- Risk of moderate poor neonatal adaptation syndrome (PNAS)** ↑ | **- Indirect benefits through better controlled maternal antenatal depression** associated with a reduced risk of harmful maternal behaviors, prematurity, intrauterine growth restriction etc ⥣<br>**- Indirect through reduced likelihood of postpartum depression (caveat: dose-effect relationship is unclear)** ⥣ |

↑ = likelihood demonstrably increases with dose adjustment (evidence on dose-effect relationship)
⥣ = likelihood theoretically increases with dose adjustment (no evidence on dose-effect relationship)
↓ = likelihood demonstrably decreases with dose adjustment (evidence on dose-effect relationship)
⥥ = likelihood theoretically decreases with dose adjustment (no evidence on dose-effect relationship)
↔ = likelihood unchanged with dose adjustment

**Bold: risks that carry relatively more weight**
*Green arrow: beneficial effects of dose adjustment*
*Blue arrow: neutral effects of dose adjustment*
*Red arrow: detrimental effects*

After examining the overall balance between the maternal and fetal risks and benefits related to the proposed sertraline dose in pregnancy, the Working Committee arrived at the following conclusions:

- Even in the absence of a proven dose-response relationship between sertraline and **fetal adverse events** apart from PNAS, a higher sertraline dose may theoretically increase the probability of certain fetal outcomes, mainly persistent pulmonary hypertension of the newborn. However, given the low absolute incidence of this risk, the self-limiting nature of PNAS, and the alignment of the proposed dosing strategy with the standard dose range for non-pregnant adults, potential excess fetal harm from the suggested dosing strategy in pregnancy is expected to remain limited, and thus acceptable.

- On the other hand, an individually titrated sertraline dose, tailored to achieve better control of maternal depression and/or anxiety symptoms is likely to be associated with considerable benefits for both mother and child. **Maternal benefits** include enhanced well-being and reduced stress, and are likely to extend beyond pregnancy as better antenatal control of maternal depression has been shown to reduce the chances of postpartum depression. In turn the maternal benefits derived from the proposed sertraline dose are demonstrably associated with **indirect fetal benefits** including reduced maternal stress, diminished risks of harmful maternal behaviors (e.g. substance abuse), as well as a reduced incidence of adverse perinatal outcomes such as intrauterine growth restriction and prematurity. Reduced postpartum depression is also likely to benefit neonates e.g. through better bonding between mother and child.

<u>Comparison with risks and benefits from alternative interventions</u>

The potential risks and benefits of the proposed sertraline dosing strategy compare favorably to the risks and benefits of **conservatively lower doses**, which may lead to inadequate control of maternal depression and/or anxiety disorders and indirect fetal harm.

As previously outlined (paragraph **2.3**), an alternative to increasing maternal sertraline doses is to introduce and/or intensify **non-pharmacological interventions**, mainly psychotherapy. The latter option, either as a stand-alone, or in combination with a low sertraline dose must be considered on a case-by-case basis, depending on the severity of patient symptoms.

**4.2.8. Do the previous findings impact the ranking of the drug in the proposed dose compared to alternative interventions for the chose indication(s)?**

The previous findings do not alter the selected indications for sertraline, namely antenatal depression and anxiety disorders. The indication for starting or continuing sertraline treatment during pregnancy should be considered on an individual basis taking into account the maternal and fetal benefits and risks of sertraline in line with the relevant clinical guidance (NVOG, 2012; NvVP, 2010). The previously collected data on a) the wide reference concentration range and limited toxicity of sertraline in nonpregnant adults and b) the decreasing trend in observed and simulated maternal plasma concentrations of sertraline during pregnancy add to the evidence supporting sertraline use in pregnancy.

**4.2.9. Do the previous findings impact the selected route(s) of administration for the drug?**

No, sertraline can only be administered orally.

###### 4.2.10. What is the added value of the proposed dose for clinical practice?

While it does not deviate from the dose recommendation in nonpregnant adults nor from the available dosing information in pregnancy on the Farmacotherapeutisch Kompas and Lareb, the proposed dose recommendation is based on an extensive review of the underlying evidence for antenatal dosing of sertraline. In addition, it builds on an explicit discussion of the maternal and fetal risks and benefits associated with various available dosing strategies in pregnancy with relevant stakeholders. This may help enhance certainty among HCPs regarding the dosing approach to follow in pregnancy, potentially reducing the misinformed practice of reducing sertraline doses of pregnancy out of caution. Ultimately it may help ensure more homogeneous antenatal dosing practices. It may also be helpful for HCPs to refer to the collected evidence and other considerations outlined as part of the benefit-risk analysis as part of shared decision-making on dosing with pregnant women.

#### 4.3. Implementation

###### 4.3.1. Is the proposed dose likely to be accepted by prescribers of the drug?

❖ *Context: discuss the acceptability of the proposed dose recommendation in regard to HCPs' current prescribing practices and preferences. Describe any expected hurdle and how these could be addressed as part of the implementation strategy for the proposed dose recommendation.*

Working Committee of project MADAM

Based on the risk-benefit analysis conducted in 4.2.7, the Working Committee deems the proposed dose recommendation acceptable for use in standard clinical practice.

Healthcare practitioners in the Netherlands

The proposed dosing strategy does not significantly differ from the standard sertraline dosing for nonpregnant adults and provides more evidence to support sertraline dosing decisions in pregnancy. Because it suggests to maintain and/or increase the dose that would otherwise be used outside of pregnancy, the proposed dosing strategy may however deviate from current practice which in some cases consists in reducing the dose or even halting sertraline use out of concern for fetal safety. Implementation of the proposed dose recommendation should therefore involve awareness raising and careful communication towards HCPs, especially GPs whose levels of experience and confidence with medication dosing in pregnancy may be more limited. This communication should focus on a) the availability of an evidence-based dose recommendation in pregnancy on the Lareb/TIS website and b) information on the key underlying dosing considerations and evidence.

###### 4.3.2. Is the proposed dose recommendation likely to be accepted by pregnant women and their partners?

- *<u>Context</u>: discuss the acceptability of the proposed dose recommendation and any remaining knowledge gaps and concerns according to pregnant women and their partners. Describe how these concerns could be addressed as part of the implementation strategy for the proposed dose including aspects such as patient information and clinician-patient communication.*

<u>Working Committee</u>
The proposed dose is deemed acceptable for use by representatives of pregnant women and their partner in the Working Committee.

<u>Pregnant women and partners in the Netherlands</u>
Pregnant women and/or their partners may have concerns about the fetal safety of the proposed dose recommendation in pregnancy. It is therefore important to ensure that HCPs in first and secondary care feel equipped to explain the key considerations behind the chosen dose - including the main physiological changes affecting the behavior and effect of medications in pregnancy- to them. This entails education of HCPs on the concepts of physiologically-based dosing in pregnancy, potential aids to support clinician-patient communication on this topic and online patient information that can be consulted by pregnant women and their partners. All the previous elements will be made available online as part of the pilot deployment of online dose recommendations in pregnancy as part of project MADAM.

##### 4.3.3. What are the next steps for implementation?

The next steps for implementation the proposed dose involve publishing the dose on the Dutch TIS website and including the dose in clinical guidelines for various medical specialists, including the guidelines for obstetrician-gynecologists and GPs. The implementation of the dose on the TIS website and in guidelines will further ensure its adoption and adherence in clinical care.

#### PART 5 – REFERENCES

1. Alhadab AA, Brundage RC. Population Pharmacokinetics of Sertraline in Healthy Subjects: a Model-Based Meta-analysis. AAPS J. 2020;22(4):73.
2. Almurjan A, Macfarlane H, Badhan RKS. The application of precision dosing in the use of sertraline throughout pregnancy for poor and ultrarapid metabolizer CYP 2C19 subjects: A virtual clinical trial pharmacokinetics study. Biopharm Drug Dispos. 2021;42(6):252-62.
3. Baumann P, Ulrich S, Eckermann G, Gerlach M, Kuss HJ, Laux G, et al. The AGNP-TDM Expert Group Consensus Guidelines: focus on therapeutic monitoring of antidepressants. Dialogues Clin Neurosci. 2005;7(3):231-47.
4. Bodnar LM, Sunder KR, Wisner KL. Treatment with selective serotonin reuptake inhibitors during pregnancy: deceleration of weight gain because of depression or drug? Am J Psychiatry. 2006;163(6):986-91.
5. Braten LS, Haslemo T, Jukic MM, Ingelman-Sundberg M, Molden E, Kringen MK. Impact of CYP2C19 genotype on sertraline exposure in 1200 Scandinavian patients. Neuropsychopharmacology. 2020;45(3):570-6.
6. Brumbaugh JE, Ball CT, Crook JE, Stoppel CJ, Carey WA, Bobo WV. Poor Neonatal Adaptation After Antidepressant Exposure During the Third Trimester in a Geographically Defined Cohort. Mayo Clin Proc Innov Qual Outcomes. 2023;7(2):127-39.
7. Brummelte S, Galea LA. Postpartum depression: Etiology, treatment and consequences for maternal care. Horm Behav. 2016;77:153-66.
8. Colombo A, Giordano F, Giorgetti F, Di Bernardo I, Bosi MF, Varinelli A, et al. Correlation between pharmacokinetics and pharmacogenetics of Selective Serotonin Reuptake Inhibitors and Selective Serotonin and Noradrenaline Reuptake Inhibitors and maternal and neonatal outcomes: Results from a naturalistic study in patients with affective disorders. Hum Psychopharmacol. 2021;36(3):e2772.
9. Desaunay P, Eude LG, Dreyfus M, Alexandre C, Fedrizzi S, Alexandre J, et al. Benefits and Risks of Antidepressant Drugs During Pregnancy: A Systematic Review of Meta-analyses. Paediatr Drugs. 2023;25(3):247-65.
10. DeVane CL, Liston HL, Markowitz JS. Clinical pharmacokinetics of sertraline. Clin Pharmacokinet. 2002;41(15):1247-66.
11. Dickmann LJ, Isoherranen N. Quantitative prediction of CYP2B6 induction by estradiol during pregnancy: potential explanation for increased methadone clearance during pregnancy. Drug Metab Dispos. 2013;41(2):270-4.
12. Drugbank. Sertraline 2023 [Available from: https://go.drugbank.com/drugs/DB01104.
13. EMA. <EMA SmPC sertraline.pdf>. 2009.
14. Fabre LF, Abuzzahab FS, Amin M, Claghorn JL, Mendels J, Petrie WM, et al. Sertraline safety and efficacy in major depression: a double-blind fixed-dose comparison with placebo. Biol Psychiatry. 1995;38(9):592-602.
15. FDA. FDA SmPC sertraline. 2016.
16. Fischer Fumeaux CJ, Morisod Harari M, Weisskopf E, Eap CB, Epiney M, Vial Y, et al. Risk-benefit balance assessment of SSRI antidepressant use during pregnancy and lactation based on best available evidence - an update. Expert Opin Drug Saf. 2019;18(10):949-63.
17. Freeman MP, Nolan PE, Jr., Davis MF, Anthony M, Fried K, Fankhauser M, et al. Pharmacokinetics of sertraline across pregnancy and postpartum. J Clin Psychopharmacol. 2008;28(6):646-53.

33. Isbister GK, Bowe SJ, Dawson A, Whyte IM. Relative toxicity of selective serotonin reuptake inhibitors (SSRIs) in overdose. J Toxicol Clin Toxicol. 2004;42(3):277-85.
34. Ke AB, Nallani SC, Zhao P, Rostami-Hodjegan A, Unadkat JD. Expansion of a PBPK model to predict disposition in pregnant women of drugs cleared via multiple CYP enzymes, including CYP2B6, CYP2C9 and CYP2C19. Br J Clin Pharmacol. 2014;77(3):554-70.
35. KNMP Kennisbank Sertraline [Internet].
36. KNMP, NVVP, NHG. Multidisciplinair document 'Afbouwen met SSRI & SNRI's'. 2018.
37. Kobayashi K, Ishizuka T, Shimada N, Yoshimura Y, Kamijima K, Chiba K. Sertraline N-demethylation is catalyzed by multiple isoforms of human cytochrome P-450 in vitro. Drug Metab Dispos. 1999;27(7):763-6.
38. Koh KH, Jurkovic S, Yang K, Choi SY, Jung JW, Kim KP, et al. Estradiol induces cytochrome P450 2B6 expression at high concentrations: implication in estrogen-mediated gene regulation in pregnancy. Biochem Pharmacol. 2012;84(1):93-103.
39. Kompas F. Sertraline [Available from: https://www.farmacotherapeutischkompas.nl/bladeren/preparaatteksten/s/sertraline.
40. Lareb B. Databank Sertraline 2020 [
]. Available from: https://www.lareb.nl/nl/databank/result?formGroup=&atc=N06AB06&ref=fk.
41. Lareb B. SSRI's tijdens de borstvoedingsperiode 2021 [cited 2023. Available from: https://www.lareb.nl/mvm-kennis-pagina?id=72&naam=SSRI's+tijdens+de+borstvoedingsperiode.
42. Lau GT, Horowitz BZ. Sertraline overdose. Acad Emerg Med. 1996;3(2):132-6.
43. Leutritz AL, van Braam L, Preis K, Gehrmann A, Scherf-Clavel M, Fiedler K, et al. Psychotropic medication in pregnancy and lactation and early development of exposed children. Br J Clin Pharmacol. 2023;89(2):737-50.
44. Levinson-Castiel R, Merlob P, Linder N, Sirota L, Klinger G. Neonatal abstinence syndrome after in utero exposure to selective serotonin reuptake inhibitors in term infants. Arch Pediatr Adolesc Med. 2006;160(2):173-6.
45. Lokuge S, Frey BN, Foster JA, Soares CN, Steiner M. Depression in women: windows of vulnerability and new insights into the link between estrogen and serotonin. J Clin Psychiatry. 2011;72(11):e1563-9.
46. Lundmark J, Reis M, Bengtsson F. Therapeutic drug monitoring of sertraline: variability factors as displayed in a clinical setting. Ther Drug Monit. 2000;22(4):446-54.
47. Malm H, Klaukka T, Neuvonen PJ. Risks associated with selective serotonin reuptake inhibitors in pregnancy. Obstet Gynecol. 2005;106(6):1289-96.
48. Masarwa R, Bar-Oz B, Gorelik E, Reif S, Perlman A, Matok I. Prenatal exposure to selective serotonin reuptake inhibitors and serotonin norepinephrine reuptake inhibitors and risk for persistent pulmonary hypertension of the newborn: a systematic review, meta-analysis, and network meta-analysis. Am J Obstet Gynecol. 2019;220(1):57 e1- e13.
49. Mauri MC, Laini V, Cerveri G, Scalvini ME, Volonteri LS, Regispani F, et al. Clinical outcome and tolerability of sertraline in major depression: a study with plasma levels. Prog Neuropsychopharmacol Biol Psychiatry. 2002;26(3):597-601.

50. MC E. Farmacogenetica: Klinische chemie 2021 [Available from: https://www.erasmusmc.nl/nl-nl/patientenzorg/laboratoriumspecialismen/farmacogenetica#f6cc3cd3-2afb-4ede-a18a-34a5fbf7cddf.
51. McGready R, Stepniewska K, Edstein MD, Cho T, Gilveray G, Looareesuwan S, et al. The pharmacokinetics of atovaquone and proguanil in pregnant women with acute falciparum malaria. Eur J Clin Pharmacol. 2003;59(7):545-52.
52. McGready R, Stepniewska K, Seaton E, Cho T, Cho D, Ginsberg A, et al. Pregnancy and use of oral contraceptives reduces the biotransformation of proguanil to cycloguanil. Eur J Clin Pharmacol. 2003;59(7):553-7.
53. Meyer JH, Wilson AA, Sagrati S, Hussey D, Carella A, Potter WZ, et al. Serotonin transporter occupancy of five selective serotonin reuptake inhibitors at different doses: an [11C]DASB positron emission tomography study. Am J Psychiatry. 2004;161(5):826-35.
54. Lexicomp Midromedex Sertraline [Internet]. 2023.
55. Milosavljevic F, Bukvic N, Pavlovic Z, Miljevic C, Pesic V, Molden E, et al. Association of CYP2C19 and CYP2D6 Poor and Intermediate Metabolizer Status With Antidepressant and Antipsychotic Exposure: A Systematic Review and Meta-analysis. JAMA Psychiatry. 2021;78(3):270-80.
56. Molenaar NM, Bais B, Lambregtse-van den Berg MP, Mulder CL, Howell EA, Fox NS, et al. The international prevalence of antidepressant use before, during, and after pregnancy: A systematic review and meta-analysis of timing, type of prescriptions and geographical variability. J Affect Disord. 2020;264:82-9.
57. Molenaar NM, Lambregtse-van den Berg MP, Bonsel GJ. Dispensing patterns of selective serotonin reuptake inhibitors before, during and after pregnancy: a 16-year population-based cohort study from the Netherlands. Arch Womens Ment Health. 2020;23(1):71-9.
58. TDM Monografie SSRI [Internet]. 2018.
59. Morishita S, Kinoshita T. Predictors of response to sertraline in patients with major depression. Hum Psychopharmacol. 2008;23(8):647-51.
60. Murdoch D, McTavish D. Sertraline. A review of its pharmacodynamic and pharmacokinetic properties, and therapeutic potential in depression and obsessive-compulsive disorder. Drugs. 1992;44(4):604-24.
61. Obach RS, Cox LM, Tremaine LM. Sertraline is metabolized by multiple cytochrome P450 enzymes, monoamine oxidases, and glucuronyl transferases in human: an in vitro study. Drug Metab Dispos. 2005;33(2):262-70.
62. Oh J, Ahn S. Predictors of Antenatal Depression in Pregnant Couples. Clin Nurs Res. 2022;31(5):881-90.
63. Paulzen M, Goecke TW, Stickeler E, Grunder G, Schoretsanitis G. Sertraline in pregnancy - Therapeutic drug monitoring in maternal blood, amniotic fluid and cord blood. J Affect Disord. 2017;212:1-6.
64. Pogliani L, Falvella FS, Cattaneo D, Pileri P, Moscatiello AF, Cheli S, et al. Pharmacokinetics and Pharmacogenetics of Selective Serotonin Reuptake Inhibitors During Pregnancy: An Observational Study. Ther Drug Monit. 2017;39(2):197-201.
65. Poweleit EA, Cinibulk MA, Novotny SA, Wagner-Schuman M, Ramsey LB, Strawn JR. Selective Serotonin Reuptake Inhibitor Pharmacokinetics During Pregnancy: Clinical and Research Implications. Front Pharmacol. 2022;13:833217.

66. Preskorn SH, Lane RM. Sertraline 50 mg daily: the optimal dose in the treatment of depression. Int Clin Psychopharmacol. 1995;10(3):129-41.
67. Prevost RR, Akl SA, Whybrew WD, Sibai BM. Oral nifedipine pharmacokinetics in pregnancy-induced hypertension. Pharmacotherapy. 1992;12(3):174-7.
68. Psychiatrie NVvd. <Leidraad farmacogenetica voor de dagelijkse psychiatrische praktijk.pdf>. 2020.
69. Quinney SK, Bies RR, Grannis SJ, Bartlett CW, Mendonca E, Rogerson CM, et al. The MPRINT Hub Data, Model, Knowledge and Research Coordination Center: Bridging the gap in maternal-pediatric therapeutics research through data integration and pharmacometrics. Pharmacotherapy. 2023.
70. Rampono J, Proud S, Hackett LP, Kristensen JH, Ilett KF. A pilot study of newer antidepressant concentrations in cord and maternal serum and possible effects in the neonate. Int J Neuropsychopharmacol. 2004;7(3):329-34.
71. Rampono J, Simmer K, Ilett KF, Hackett LP, Doherty DA, Elliot R, et al. Placental transfer of SSRI and SNRI antidepressants and effects on the neonate. Pharmacopsychiatry. 2009;42(3):95-100.
72. Reis M, Aamo T, Spigset O, Ahlner J. Serum concentrations of antidepressant drugs in a naturalistic setting: compilation based on a large therapeutic drug monitoring database. Ther Drug Monit. 2009;31(1):42-56.
73. Rowland Yeo K, Walsky RL, Jamei M, Rostami-Hodjegan A, Tucker GT. Prediction of time-dependent CYP3A4 drug-drug interactions by physiologically based pharmacokinetic modelling: impact of inactivation parameters and enzyme turnover. Eur J Pharm Sci. 2011;43(3):160-73.
74. Ryu RJ, Eyal S, Easterling TR, Caritis SN, Venkataraman R, Hankins G, et al. Pharmacokinetics of metoprolol during pregnancy and lactation. J Clin Pharmacol. 2016;56(5):581-9.
75. Schoretsanitis G, Spigset O, Stingl JC, Deligiannidis KM, Paulzen M, Westin AA. The impact of pregnancy on the pharmacokinetics of antidepressants: a systematic critical review and meta-analysis. Expert Opin Drug Metab Toxicol. 2020;16(5):431-40.
76. Shah M, Xu M, Shah P, Wang X, Clark SM, Costantine M, et al. Effect of CYP2C9 Polymorphisms on the Pharmacokinetics of Indomethacin During Pregnancy. Eur J Drug Metab Pharmacokinet. 2019;44(1):83-9.
77. Shelton RC. Steps Following Attainment of Remission: Discontinuation of Antidepressant Therapy. Prim Care Companion J Clin Psychiatry. 2001;3(4):168-74.
78. Singh HK, Saadabadi A. Sertraline. StatPearls. Treasure Island (FL)2023.
79. Sit DK, Perel JM, Helsel JC, Wisner KL. Changes in antidepressant metabolism and dosing across pregnancy and early postpartum. J Clin Psychiatry. 2008;69(4):652-8.
80. Soma-Pillay P, Nelson-Piercy C, Tolppanen H, Mebazaa A. Physiological changes in pregnancy. Cardiovasc J Afr. 2016;27(2):89-94.
81. Sprouse J, Clarke T, Reynolds L, Heym J, Rollema H. Comparison of the effects of sertraline and its metabolite desmethylsertraline on blockade of central 5-HT reuptake in vivo. Neuropsychopharmacology. 1996;14(4):225-31.

82. Stika CS, Wisner KL, George AL, Jr., Avram MJ, Zumpf K, Rasmussen-Torvik LJ, et al. Changes in Sertraline Plasma Concentrations Across Pregnancy and Postpartum. Clin Pharmacol Ther. 2022;112(6):1280-90.
83. Tomson T, Lindbom U, Ekqvist B, Sundqvist A. Disposition of carbamazepine and phenytoin in pregnancy. Epilepsia. 1994;35(1):131-5.
84. Tracy TS, Venkataramanan R, Glover DD, Caritis SN, National Institute for Child H, Human Development Network of Maternal-Fetal-Medicine U. Temporal changes in drug metabolism (CYP1A2, CYP2D6 and CYP3A Activity) during pregnancy. Am J Obstet Gynecol. 2005;192(2):633-9.
85. Underwood L, Waldie K, D'Souza S, Peterson ER, Morton S. A review of longitudinal studies on antenatal and postnatal depression. Arch Womens Ment Health. 2016;19(5):711-20.
86. Unterecker S, Riederer P, Proft F, Maloney J, Deckert J, Pfuhlmann B. Effects of gender and age on serum concentrations of antidepressants under naturalistic conditions. J Neural Transm (Vienna). 2013;120(8):1237-46.
87. UpToDate. Selective serotonin reuptake inhibitors: Pharmacology, administration, and side-effects 2023 [
88. van de Loo KFE, Vlenterie R, Nikkels SJ, Merkus P, Roukema J, Verhaak CM, et al. Depression and anxiety during pregnancy: The influence of maternal characteristics. Birth. 2018;45(4):478-89.
89. Ververs FF, Voorbij HA, Zwarts P, Belitser SV, Egberts TC, Visser GH, et al. Effect of cytochrome P450 2D6 genotype on maternal paroxetine plasma concentrations during pregnancy. Clin Pharmacokinet. 2009;48(10):677-83.
90. Wadelius M, Darj E, Frenne G, Rane A. Induction of CYP2D6 in pregnancy. Clin Pharmacol Ther. 1997;62(4):400-7.
91. Walsh-Sukys MC, Tyson JE, Wright LL, Bauer CR, Korones SB, Stevenson DK, et al. Persistent pulmonary hypertension of the newborn in the era before nitric oxide: practice variation and outcomes. Pediatrics. 2000;105(1 Pt 1):14-20.
92. Wang JH, Liu ZQ, Wang W, Chen XP, Shu Y, He N, et al. Pharmacokinetics of sertraline in relation to genetic polymorphism of CYP2C19. Clin Pharmacol Ther. 2001;70(1):42-7.
93. Wangboonskul J, White NJ, Nosten F, ter Kuile F, Moody RR, Taylor RB. Single dose pharmacokinetics of proguanil and its metabolites in pregnancy. Eur J Clin Pharmacol. 1993;44(3):247-51.
94. Westin AA, Brekke M, Molden E, Skogvoll E, Spigset O. Selective serotonin reuptake inhibitors and venlafaxine in pregnancy: Changes in drug disposition. PLoS One. 2017;12(7):e0181082.
95. Wu J, Viguera A, Riley L, Cohen L, Ecker J. Mood disturbance in pregnancy and the mode of delivery. Am J Obstet Gynecol. 2002;187(4):864-7.
96. Xu ZH, Wang W, Zhao XJ, Huang SL, Zhu B, He N, et al. Evidence for involvement of polymorphic CYP2C19 and 2C9 in the N-demethylation of sertraline in human liver microsomes. Br J Clin Pharmacol. 1999;48(3):416-23.
97. Yin X, Sun N, Jiang N, Xu X, Gan Y, Zhang J, et al. Prevalence and associated factors of antenatal depression: Systematic reviews and meta-analyses. Clin Psychol Rev. 2021;83:101932.

98. Zhao S, Gockenbach M, Grimstein M, Sachs HC, Mirochnick M, Struble K, et al. Characterization of Plasma Protein Alterations in Pregnant and Postpartum Individuals Living With HIV to Support Physiologically-Based Pharmacokinetic Model Development. Front Pediatr. 2021;9:721059.
99. Zheng L, Yang H, Dallmann A. Antidepressants and Antipsychotics in Human Pregnancy: Transfer Across the Placenta and Opportunities for Modeling Studies. J Clin Pharmacol. 2022;62 Suppl 1:S115-S28

### PART 6 – VERSION CONTROL

| | |
|---|---|
| **Version** | **3** |

| | |
|---|---|
| **Date** | 18 November 2023 |
| **Authors** | |
| **Clinical experts consulted** | |
| **Pharmacological experts consulted** | |
| **Modelling experts consulted** | |
| **Reviewers of the dose rationale document** | |
| **Authorization date MADAM Working Committee** | **20 December 2023** |

#### 2.b – Supplements FDSP sertraline

**Supplement 1: Literature searches**

**General search on the pharmacokinetics and pharmacodynamics of sertraline (06-02-2023)**

| | |
|---|---|
| **Medication** | ((Sertraline[MeSH Terms] OR Sertraline[Title/Abstract] OR Zoloft[Title/Abstract] OR Lustral[Title/Abstract] OR Serlain[Title/Abstract] OR Besitran[Title/Abstract]) |
| **Pregnancy** | AND ((Pregnancy[MeSH] OR 'pregnant women'[MeSH] OR Pregnan*[tiab] OR childbearing*[tiab] OR child-bearer*[tiab] OR 'With child'[tiab] OR expecting[tiab] OR mother*[tiab] OR embryo*[tiab] OR fetus[MeSH] OR fetus*[tiab] OR foetus*[tiab] OR fetal*[tiab] OR foetal*[tiab] OR baby*[tiab] OR "Infant, Newborn"[MeSH] OR newborn*[tiab] OR neonate*[tiab] OR infant*[tiab] OR unborn*[tiab] OR prenatal[tiab] OR antenatal[tiab] OR labor[tiab] OR labour[tiab] OR delivery[tiab] OR birth[tiab] OR perinatal[tiab] OR postpartum[tiab] OR "Fetal Blood"[Mesh] OR umbilical*[tiab] OR placenta*[tiab] OR "Amniotic Fluid"[Mesh] OR amniot*[tiab])) |
| **Pharmacokinetics/ Pharmacodynamics** | AND ((Sertraline/administration and dosage[Majr:NoExp] OR Sertraline/pharmacokinetics[Majr:NoExp] OR Sertraline/blood[Majr:NoExp] OR Sertraline/therapeutic use[Majr:NoExp] OR Pharmacodynam*[tiab] OR Dose*[tiab] OR Dosi*[tiab] OR Dosage*[tiab] OR efficac*[tiab] OR effectiv*[tiab] OR kinetic*[Tiab] OR concentration[Tiab] OR level[Tiab] OR expos*[tiab]<br><br>OR "Sertraline/adverse effects"[Majr:NoExp] OR safe*[tiab] OR teratogen*[tiab] OR toxic*[tiab] OR outcome*[tiab])) |
| **Human** | NOT (animals[mesh] NOT humans[mesh]) |

**Safety Studies (14-03-2023)**

(Sertraline OR Sertraline[Pharmacological Action] OR Sertraline[Title/Abstract] OR Zoloft[Title/Abstract] OR Lustral[Title/Abstract] OR Besitran[Title/Abstract] OR Lustral[Title/Abstract]) AND (Pregnancy[MeSH] OR pregnant women[MeSH] OR maternal health[ti] OR maternal safety[ti] OR maternal complication*[tiab] OR birth*[tiab] OR child-bear*[tiab] OR childbear*[tiab] OR gestation[tiab] OR mother*[tiab] OR postpartum[tiab] OR Pregnan*[ti]) AND (Sertraline/pharmacokinetics[Mesh:NoExp] OR Sertraline/blood[Mesh:NoExp] OR Sertraline/therapeutic use[Mesh:NoExp] OR Sertraline/adverse effects[Mesh:NoExp] OR "administration"[tiab] OR "pharmacokinetic*"[tiab] OR Pharmacodynam*[tiab] OR efficac*[tiab] OR effectiv*[tiab] OR kinetic*[Tiab] OR expos*[tiab] OR safe*[tiab] OR toxic*[tiab] OR outcome*[tiab]) AND (Sertraline/administration and dosage[Mesh:NoExp] OR "Drug Monitoring"[Mesh] OR "Drug Dosage Calculations"[Mesh] OR Dose*[tiab] OR Dosi*[tiab] OR Dosage*[tiab] OR concentration*[Tiab] OR level[ti] or levels[ti]) NOT (Animals[Mesh] NOT Humans[Mesh])

**Efficacy studies (11-03-2023)**

(Sertraline[Title/Abstract] OR Zoloft[Title/Abstract] OR Lustral[Title/Abstract] OR Besitran[Title/Abstract] OR Lustral[Title/Abstract]) AND (efficac*[tiab] OR effectiv*[tiab] OR kinetic*[Tiab] OR expos*[tiab] OR safe*[tiab] OR toxic*[tiab] OR outcome*[tiab]) AND (Sertraline/administration and dosage[Mesh:NoExp] OR "Drug Monitoring"[Mesh]) NOT (Animals[Mesh] NOT Humans[Mesh])

#### Supplement 2: Quality & credibility assessment of PBPK simulations of sertraline

| |
|---|
| **Step 1. Assess PBPK model credibility** |

<table>
<tr><td colspan="5">Model parameterization</td></tr>
<tr><td rowspan="2">1</td><td>Is the drug file adequately parameterized (e.g. contributing eliminating pathways)?</td><td>No</td><td>Q</td><td>Yes</td></tr>
<tr><td colspan="4">The compound file from Templeton et al. 2016, which was used to build a PBPK model to assess CYP3A4- and CYP2D6 mediated drug-drug interactions with sertraline, was used. Data on hepatic clearance was obtained from Templeton et al. 2016 (CYP2C9, CYP2C19 and CYP2B6) and Obach et al. 2005 (CYP3A4 and CYP2D6). Modifications were performed in the compound file to allow for use in gestation.</td></tr>
<tr><td rowspan="2">2</td><td>Is the physiology file adequately parameterized (e.g. incorporation of biological variability and relevant ontogeny profiles)?</td><td>No</td><td>Q</td><td>Yes</td></tr>
<tr><td colspan="4">Almurjan et al. used the Simcyp pregnancy population. The extent to which pregnancy-induced changes in ADME processes likely to be of clinical relevance to sertraline are captured in the latter file is discussed below.<br><br>Maternal metabolism<br><u>Absorption</u><br>Simcyp does not incorporate pregnancy-induced changes in absorption other than changes in first pass effect as a result of altered CYP metabolism. The latter changes represent the main variables of importance for the absorption of sertraline, which is thus well captured.<br><u>Distribution</u><br>Simcyp incorporates changes in the plasma volume and fat composition of the body as well as changes in plasma albumin as part of pregnancy. These constitute all the main likely variables of change for the distribution of sertraline (a highly protein bound, lipophilic medication) in pregnancy.<br><u>Metabolism</u><br>The metabolism of sertraline depends on five polymorphic enzymes, CYP2B6, CYP2C9, CYP2C19, CYP2D6 and CYP3A4, none of which is responsible for more than 30% of the hepatic metabolism of sertraline (Obach et al. 2005; Kobayashi et al. 1999; Greenblatt et al. 1999).<br><br>Maternal CYP ontogeny<br>Pregnancy-induced changes in maternal CYP expression are incorporated in Simcyp for CYP3A4 and CYP2D6 (Abduljalil et al., 2012). Almurjan et al. added pregnancy-related changes in the expression of CYP2B6 and CYP2C19 based on studies by Ke, Greupink, & Abduljalil, 2018; McGready, Stepniewska, Edstein, et al., 2003 and McGready, Stepniewska, Seaton, et al., 2003). Changes in CYP2C9 expression during pregnancy are not accounted for. CYP2C9 is estimated to contribute to 14-29% of sertraline N-demethylation according to in vitro studies in human liver microsomes (Obach et al. 2005; Kobayashi et al. 1999; Greenblatt et al. 1999). CYP2C9 activity was estimated to increase by 1.6 fold by the third trimester of pregnancy according to in vitro in vivo extrapolations (Ke et al. 2014).<br><br>Maternal CYP polymorphisms<br>The CYP2C19 phenotype frequency described in Simcyp was used (ultrarapid metabolizer (UM) 31.8%, extensive metabolizer (EM) 59% and poor metabolizer (PM) 9.2%) which roughly aligns with the phenotype frequencies described by the CPIC 2015 (UM 5-30%, EM 35-50% and PM 2-15%)</td></tr>
</table>

| | | | | |
|---|---|---|---|---|
| | (Hicks et al. 2015). Enzyme abundance for various CYP2C19 phenotypes was incorporated based on Djebli et al. 2015 (a PBPK study of the effect of CYP2C19 genetic polymorphisms on clopidogrel; no references cited in this study on how enzyme abundance was determined). **It is unclear whether polymorphisms in other CYP enzymes involved in sertraline metabolism was incorporated in the model**. While CYP2C19 polymorphisms are considered to be the major pharmacogenetic driver of variability in sertraline pharmacokinetics in nonpregnant adults (Greenblatt et al. 1999; CPIC 2015; Huddart et al. 2020), the combined influence of polymorphisms in the four other enzymes involved in sertraline metabolism may affect the pharmacokinetics of this drug.<br><br>Elimination<br>Sertraline elimination is mainly dependent on hepatic metabolism, described above.<br><br>*Placental and fetal metabolism*<br>**Simcyp does not incorporate the placental and fetal metabolism of medications in its pregnancy population.** So far little is known on how the latter processes may affect sertraline disposition in pregnancy. That said, given the low fetal metabolic capacity resulting from the absent to low expression of most CYP enzymes (Shea et al., 2012), fetal metabolism is unlikely to play an important role in the disposition of sertraline. Almurjan et al.'s model thus appears to be in line with the current state of knowledge on the placental and fetal metabolism of sertraline.<br><br>Blood-brain-barrier<br>As an SSRI, sertraline exerts its effect by inhibiting serotonin reuptake in the cerebral synapses. Pregnancy-induced changes in the blood-brain-barrier may therefore alter the effect of sertraline. While the latter changes are not captured in the Simcyp pregnancy population, no additional information can be found in the literature suggesting clinically relevant alterations in the blood-brain-barrier as a result of pregnancy (this is also true for other psychoactive medications). | | | |
| 3 | **Are there any assumptions made regarding model input parameters?**<br>**If yes, comment on the influence of change in model input parameters on modeling output.** | Yes | **Q** | No |
| | See previous answer. | | | |
| **Model verification** | | | | |
| 4 | **Is model performance verified using *in vivo* PK data from a population using the similar drug, for a similar indication and of comparable age? If no, comment on the data used for model verification (which age, indication and/or other drug with similar ADME properties) and whether PK may be different in *in vivo* population compared to healthy virtual individuals.** | No | *NA* | **Yes** |

| | | | | |
|---|---|---|---|---|
| | <u>Verification in nonpregnant adults</u><br>The model was verified in nonpregnant adults using in vivo PK data from five studies (four single dose studies and one multiple dose study).<br><br><u>Verification in pregnancy</u><br>**The model was verified in pregnancy using in vivo data from pregnant subjects in one study (Westin et al., 2017).**<br>Although multiple other PK studies of sertraline in pregnancy published prior to Almurjan et al.'s study can be found (e.g. Sit et al., 2008, Freeman et al. 2008), Westin et al.'s study was chosen for validation purposes as it included a relatively large study population of pregnant women (N = 34, more so than other PK studies of sertraline in pregnancy) including individualized sample data.<br>Westin et al.'s observational study reported on 56 sertraline serum samples obtained from routine TDM performed in 34 psychiatry inpatients and outpatients in pregnancy (37 pregnancies) by the two largest TDM services in Norway. The demographic characteristics of the sampled pregnant women (including ethnicity, age) were not reported. Importantly the CYP2C19 profiles of the sampled population were not investigated by Westin et al. meaning that **Almurjan et al. did not verify the model for different CYP2C19 phenotypes.** The indications and doses of sertraline used were not described in the study. | | | |
| 5 | **Is PBPK model performance considered adequate (based on visual predictive check of predicted vs. observed PK curve and predicted-to-observed PK parameter ratios)?If no, comment on whether this be explained by differences populations (i.e., *in vivo* population vs. virtual 'healthy' population, see 5) or not.** | No | **Q** | Yes |
| | The predicted data were verified using a visual predictive check and a two-fold predicted to observed ratio (the most commonly used ratio for PBPK modelling (van der Heijden et al., 2022). Given the broad therapeutic window and limited toxicity of sertraline (Hiemke et al., 2018), the use of a two-fold ratio appears sufficient for ensuring adequate model performance.<br><br><u>Verification in nonpregnant adults</u><br>Predicted concentrations and PK parameters were all within 2-fold of the observed data from all five studies in nonpregnant adults (Figure 2).<br><br><u>Verification in pregnancy</u><br>While the distribution of simulated sertraline plasma concentrations was visually comparable to the range of in vivo concentrations reported by Westin et al. in pregnancy (Figure 3), the predicted to observed data ratio was not reported for pregnant women and could not be used to assess model performance. In addition, the predicted decrease in concentrations across pregnancy diverged from the increase in mean plasma concentrations of sertraline reported by Westin et al. | | | |
| **OVERALL MODEL CREDIBILITY** | | | | |

| | How credible are the outcomes of the PBPK model considering model parameterization, robustness and verification? | Low | Q | High |
|---|---|---|---|---|
| | **Model credibility is considered sufficient.** While **only one study was used for validation of Almurjan et al.'s model in a pregnant population** and it would have been preferable to validate the model using pooled observational data from other available PK studies, and while the concentrations simulated by Almurjan et al. followed a trend opposite to that reported in the study Westin et al. 201), **the overall credibility of the model remains satisfactory in view of the alignment of the predicted concentrations with data from other observational studies** (e.g. Sit et al. 2008, Freeman et al., 2008, Stika et al. 2022, which all reported a decrease in maternal sertraline concentrations across pregnancy) **and of the plausible pharmacokinetic reasons for a decrease in sertraline concentration throughout pregnancy**. These arguments include a) the induction of the four other CYP enzymes involved in sertraline metabolism as a result of hormonal changes in pregnancy as reported by multiple in vivo and vitro studies which may partially counterbalance the inhibition of CYP2C19 (of note the overall hepatic clearance of sertraline was still predicted to diminish in pregnancy) and b) other pregnancy-associated physiological changes of potentially greater magnitude (e.g. a decrease in the concentration of albumin which was demonstrated to be a covariate for the predicted sertraline concentrations through a global sensitivity analysis performed by Almurjan et al., especially in the third trimester of pregnancy). | | | |

| **Step 2. Establish a model-informed dose** | | | | | | | |
|---|---|---|---|---|---|---|---|
| **Dose-finding method** | | | | | | | |
| 1 | **Which dose-finding method was applied (see Figure 1 below)?**<br>**If model-informed dose is based on exposure matching, comment on the population for which the plasma exposure target (AUC or $C_{max}$) is established. Also, comment on whether the plasma target is a surrogate exposure measurement for another compartment (e.g., lungs) and hence a similar plasma:target compartment is assumed between the populations.** | 1 | 2 | 3 | 4 | 5 | **Other** |

| | | | | |
|---|---|---|---|---|
| | The plasma concentration of sertraline was used as a surrogate marker for intracerebral concentrations, as is typically done outside of pregnancy. The target therapeutic range defined in Braten et al. 2020 was used for dose-finding**. A therapeutic range has been hard to define given the lack of a clear dose-exposure-response relationship for sertraline** (Huddart et al. 2020). Proposed ranges in the literature are mostly based on in vivo pharmacokinetic data associated with routine sertraline use in nonpregnant adults as opposed to well-substantiated pharmacokinetic-pharmacodynamic endpoints. The Arbeitsgemeinschaft für Neuropsychopharmakologie und Pharmakopsychiatrie (AGNP) recommends a 10-150 ng/ml range based on multiple pharmacokinetic studies of sertraline without outlining specific pharmacodynamic arguments for the recommended range (Hiemke et al. 2018). Reflecting on the proposed AGNP therapeutic range, **Braten al. suggested a 10-75 ng/ml range as an alternative based on the 80% occupancy of serotonin transporters measured at 10 ng/ml** concentration by positron-emission tomography imaging (Meyer et al., 2004). The latter ranges are plasma concentration ranges and are assumed to be proxies for therapeutically effective concentrations of sertraline at its site of action, namely the brain.<br><br>Almurjan et al.'s optimal doses were defined in such a way that less than 20% subjects had **predicted trough concentrations** falling outside the therapeutic range of 10-75 ng/ml. For CYP2C19 poor metabolizers (PMs) however, the recommended dose of 50 mg daily for women in their first trimester of pregnancy led to a subtherapeutic concentration (< 10 ng/mL) in 24% women (Figure 6). In addition, although Almurjan et al. recommend a dose of 100 mg daily in the second and third trimesters of pregnancy for CYP2C19 PMs, a dose of 150 mg daily would also lead to < 20% pregnant women having trough concentrations above the upper limit of the therapeutic range (>75 ng/ml). This may not apply to the Cmax which was not reported by Almurjan et al. Similarly a daily dose of 150 mg in the first trimester of pregnancy for extensive and ultrarapid metabolizers (EMs or UMs) or a daily dose of 200 mg in the second and third trimesters of pregnancy for UMs also led to predicted concentrations below the recommended maximum of 75 ng/ml.<br><br>While the mean Cmax of sertraline in pregnancy was not reported by Almurjan et al. and overdosing in reference to the chosen therapeutic range may in theory occur, pregnancy is expected to reduce the swing (ratio between Cmax and Cmin for a given medication) of medications as a result of an increased Vd, Given the known Cmax of sertraline (20 to 55 ng/ml) and the decreasing trend in sertraline trough concentrations in pregnancy, Cmax appears unlikely to exceed the upper bound of the therapeutic range. | | | |
| **Pharmacokinetics** | | | | |
| 2 | **Does the drug have a narrow therapeutic window?** | Yes | Q | **No** |
| | No. The upper limit of the therapeutic range used by Almurjan et al. is twice lower than the upper bound of the therapeutic range recommended by the AGNP. A study by Unterecker et al. 2013 demonstrated that among 100 women receiving a standard daily dose of sertraline (50-200 mg), 12 were below the AGNP-defined range with none above this range, illustrating a general tendency for concentrations to occur at the lower end of the proposed therapeutic ranges. Additionally the toxicity of sertraline appears limited overall (>50% of overdoses asymptomatic, transient symptoms in the event of symptomatic overdoses (Lau et al. 1996, Isbister et al. 2004)). | | | |

| | | | | |
|---|---|---|---|---|
| 3 | **Is there a wide interindividual variability in PK? If yes, comment on whether you expect that some individuals will reach the efficacy or safety limit of the therapeutic window?** | **Yes** | Q | No |
| | Yes – De Vane et al. 2002 reported up to 15-fold variation in the steady-state sertraline plasma concentrations of nonpregnant patients receiving 50-150 mg/day while Lundmark et al. 2000 measured an 88-fold variation in sertraline concentration-dose in 319 patients receiving 25-250 mg/day. It is likely that part of this variation can be explained by genetic polymorphisms in the CYP enzymes involved in the metabolism of sertraline, especially CYP2C19 (Braten et al., 2020, Huddart et al. 2020). It is likely that the physiological changes occurring in pregnancy constitute an additional source of variability in the PK of sertraline. Given the breadth of the therapeutic window of sertraline however, the risk of crossing the lower or upper bound of this range appears limited in practice as testified by the available pharmacokinetic data for sertraline in pregnancy (Freeman et al., 2008; Sit et al., 2008; Westin et al. 2017, Stika et al. 2022 etc). | | | |

| 1 | 2 | 3 | 4 | 5 |
|---|---|---|---|---|
| An existing dose recommendation (backed by a low level of evidence) or a dose suggested by clinician | Plasma level suggested by clinician | Exposure* suggested to be effective for a **different disease** | Exposure* suggested to be effective for the **disease of interest** but **different age group** | Established PD-based plasma level target |
| e.g., "this dose always works well", does exposure seem reasonable? | e.g., "based on experience, we aim to reach this level" | e.g., AUC matching with data from another viral infection | e.g., AUC matching with adult data | e.g., time above MIC |

* Exposure target is dependent on the drug and can be AUC, $C_{max}$ or $C_{trough}$ (drug-dependent)

**Figure 1. Dose-finding method**

| | | | | |
|---|---|---|---|---|
| **Step 3. Implement the model-informed dose** | | | | |
| **Model extrapolation** | | | | |
| 1 | **Are there any differences expected in drug <u>PK</u> between the virtual population and the real-world population*? If yes, comment on the disease, co-morbidity, treatment, etc., that is likely to affect PK.** | Yes | **Minimally** | No |

<table>
<tr><td></td><td colspan="4">It is likely that Almurjan's et al.'s model does not capture all the genetic variability in the CYP enzymes involved in the metabolism of sertraline. Other sources of variability for sertraline PK in a real-world pregnant population (e.g. maternal BMI) may also be underestimated.</td></tr>
<tr><td rowspan="2">2</td><td>Are there any differences expected in drug <u>PD</u> between the virtual population and the real-world population*?<br>If yes, comment on the disease, co-morbidity, treatment, etc., that is likely to affect PD.</td><td>Yes</td><td>Minimally</td><td>No</td></tr>
<tr><td colspan="4">The PD of the virtual population is not captured by this PBPK model. It is important to note that pregnancy itself is known to affect the pharmacodynamics of anxiety and depression (high prevalence of de novo depression and anxiety symptoms in pregnancy; high prevalence of postpartum depression (Yin et al. 2021)).</td></tr>
<tr><td colspan="5">Model influence</td></tr>
<tr><td rowspan="2">3</td><td>Does the model-informed dose deviate from the current dosing advice or from dosing strategies applied in clinical care (incl. expert opinion, scientific clinical studies and dosing handbooks)?</td><td>Yes</td><td>Somewhat</td><td>No</td></tr>
<tr><td colspan="4">The proposed doses (50 mg in T1 followed by 100 mg in T2-T3 of pregnancy for poor metabolizers (PMs) and 100-150 mg throughout gestation for extensive or ultrapid metabolizers (EMs or UMs) are within the dose range that is currently recommended for nonpregnant adults (50 mg-200 mg) internationally. The doses recommended by Almurjan et al. based on modelling & simulations somewhat diverge from current practice in two regards:<br><br>a)Almurjan et al. issue distinct dose recommendations for various CYP2C19 genetic profiles while CYP2C19 genotyping is only conducted in a minority of sertraline users in the Netherlands (primarily, in the event of recurring adverse reactions to consecutive medications metabolized by CYP2C19). Both the CPIC and the KNMP outline distinct recommendations for CYP2C19 EMs/UM and PMs of nonpregnant adults when pharmacogenetic testing has been conducted however.<br><br>b) Almurjan et al. recommend not to decrease the dose of sertraline below 50 mg and later 100 mg daily for PMs and advise daily doses of 100-150 mg for EMs/UMs throughout pregnancy. This is lower than the maximum dose of 200 mg that is recommended outside of pregnancy. While Almurjan et al. does not comment on this discrepancy and while their predicted plasma concentrations for UMs would allow a 200 mg daily dose in the 2nd and 3rd trimesters of pregnancy, it is likely that Almurjan et al. were cautious in their dose recommendations given their focus on predicting sertraline trough concentrations as opposed to Cmax in pregnancy, making it more difficult to assess the potential for overdosing for the proposed dosing regimens. In fact, anecdotal evidence reveals that clinicians tend to reduce sertraline doses in pregnancy out of concern for fetal safety. Almurjan's et al. recommended doses may thus be perceived to be relatively high compared to the status quo for dosing sertraline in pregnancy.</td></tr>
<tr><td colspan="5">Decision consequence (pharmacodynamics)</td></tr>
</table>

| | | | | |
|---|---|---|---|---|
| 4 | **Are the consequences of over-or underdosing deemed acceptable?** | No | Q | Yes |
| | See paragraph 4.2.3. in main document. | | | |
| 5 | **Is it possible to monitor exposure (e.g., with TDM or a clinical measurable effect) and to act if needed?** | No | **Somewhat** | Yes |
| | Given the large interindividual variability in clinical response to sertraline, it is recommended to monitor the clinical effectiveness and occurrence of side effects in nonpregnant patients starting a course of sertraline (Dutch Society of Psychiatry, 2013). In view of the likely influence of pregnancy on the pharmacodynamics of sertraline (Amiel Castro et al. 2017), monitoring the efficacy of a newly started or pre-existing treatment with sertraline appears highly advisable in pregnancy. There are no routinely used clinical scales of anxiety or depression to this effect (this is true outside of or during pregnancy). If suboptimal efficacy or side effects are detected in an individual patient, it is possible to increase or decrease the dose of sertraline on a weekly base in line with clinical guidance for nonpregnant adults (Dutch Society of General Practitioners, 2022; Dutch Society of Psychiatry, 2013).<br><br>In the absence of a clearly defined therapeutic range for sertraline, clinical guidance on the utility of TDM of sertraline diverges. While the German Society of Neuropsychopharmacology (AGNP) recommends therapeutic monitoring of sertraline based on data from steady-state pharmacokinetic studies (Hiemke et al., 2018), the Dutch Toxicology Centre does not recommend the use of TDM for sertraline (Toxicologie.org). | | | |
| 6 | **Do the benefits outweigh the potential risks?** | No | Q | **Yes** |
| | Overall, given the wide therapeutic range of sertraline and the conservative range adopted as a reference by Almurjan et al. in combination with the predicted decrease in mean maternal concentrations of sertraline in pregnancy across all CYP2C19 phenotypes, the benefits of administering sufficiently high doses of sertraline in pregnancy following Almurjan et al.'s recommendations (with regards to the clinical effectiveness of the medication) appear to outweigh the potential risks of sertraline use in pregnancy if treatment with a SSRI is indicated in that period. A caveat refers to a potential dose-relationship found between higher daily doses (≥ 100 mg) of sertraline and the incidence of poor neonatal adaptation syndrome (PNAS), a transient condition (Brumbaugh et al. 2023), which should also be considered when increasing the dose of sertraline, especially in the third trimester of pregnancy.<br><br>Given the large interindividual variability in the severity of anxiety and depression symptoms across pregnancy (Yin et al. 2021) as well as the large interindividual variation in sertraline PK, the latter decision ought to be made on an individual basis by women planning pregnancy or pregnant women and their clinician.<br><br>Risk mitigation measures<br>Considering the potential added contribution of pregnancy to variability in response to sertraline, it appears advisable to organize additional clinical controls of sertraline users throughout pregnancy, especially at the start of treatment with sertraline (if started during or shortly before pregnancy, similar to the controls | | | |

| | | | | |
|---|---|---|---|---|
| | indicated for newly started sertraline courses in nonpregnant adults). Similar to nonpregnant adults, sertraline doses can be titrated based on a clinical response. Genetic screening for CYP2C19 polymorphisms and/or TDM can be considered in the event of unexplained adverse reactions or if a suboptimal response persists following dose increase. | | | |
| **Practical considerations** | | | | |
| 7 | **Is it possible to administer the model-informed dose, e.g. based on drug formulation, dose calculation and excipient safety?** | No | *NA* | **Yes** |
| | Given that the model-informed doses recommended by Almurjan et al. in pregnancy are within the recommended dose range of sertraline for nonpregnant adults and considering that titration can be performed following similar steps than in nonpregnant adults in the event of symptomatic underdosing or overdosing, administering sertraline according to the model-informed dose in pregnancy should not pose additional practical challenges (other than the additional burden of added clinical follow-up for pregnant women in a vulnerable time). | | | |
| 8 | **Do clinicians and patients and parents agree with the model-informed dose?** | No | **Tbc** | Yes |
| | This will be determined when the model-informed dose recommendation is reviewed by the Editorial Board of project MADAM. | | | |

* The real-world population is defined as the patient population who is intended to receive the model-informed dose.

**Supplement 3: Considerations for weight attribution as part of risk-benefit analysis of antenatal doses**

The following considerations should drive the attribution of weights to individual risks and benefits of proposed antenatal dose adjustments.

| **Magnitude of the risk (hazard)/benefit** | **Likelihood of the risk (hazard)/benefit** |
|---|---|
| Transient or permanent<br>Severe or non-severe<br>Treatable or not | What is the absolute incidence of the risk?<br>How established is the dose-effect relationship for the risk/benefit (PD mechanism)?<br>How strong is the causal link between the chosen dose and the risk/benefit (relative risk)? |

Comparing the risks & benefits, the following value ranking applies:

- Permanent > transient
- Not treatable > treatable
- High incidence of the risk/benefit > low incidence
- Experimentally established causality link between dose & risk/benefit > plausible theoretical causality link between dose & risk/benefit
- High relative risk > low relative risk
